# Conversational Speech for Respiratory Triage in Primary Care: A Pilot Study

**DOI:** 10.64898/2026.06.09.26355284

**Authors:** Vijay Ravi, Camille Noufi

**Affiliations:** Amplifier Health, Inc., San Francisco, California, USA

**Keywords:** vocal biomarkers, differential diagnosis, hierarchical classification, ambient audio, acoustic features, ICD-10 cohorts, confounder analysis, real-world clinical audio

## Abstract

**Background:** Respiratory complaints account for a substantial share of adult ambulatory visits, and triaging them accurately has direct consequences for antibiotic stewardship and pathogen-specific therapy. Prior work has investigated voice as a triage signal, but that literature is dominated by single-condition detection from scripted speech in crowdsourced or controlled clinical settings and has not been evaluated at primary care scale on conversational ambient audio.

**Methods:** A dataset of 514,377 ambient-recorded primary care visits from 379,225 adult patients at a US clinic network was used, with per-visit clinically assigned ICD-10 diagnosis codes and de-identified demographic and geographic metadata. Patient audio was extracted from each doctor-patient conversation, and spectral, voice quality, and prosodic features were computed. Eleven binary classification tasks were defined, aligned with a respiratory triage cascade (e.g., acute respiratory versus acute non-respiratory illness, and lower versus upper respiratory tract infection). An acoustic model was trained independently for each task using patient-stratified five-fold cross-validation and evaluated on a held-out test set. Each model was also compared against six non-acoustic baselines using a single demographic, geographic, or temporal variable. The 11 trained classifiers were composed into a hierarchical cascade and illustrated as case studies.

**Results:** Test-set AUC across the 11 tasks ranged from 0.602 (95% CI: 0.588–0.614) to 0.745 (95% CI: 0.742–0.748), with a mean expected calibration error of 0.018. Six of eleven binaries outperformed all six confounder baselines after multiple-testing correction. Four binaries showed median within-stratum AUC of 0.61–0.70 when the confounder was held fixed, indicating acoustic discrimination beyond what the confounder alone explains. Five binaries failed at least one axis; the only one outperformed by a confounder baseline was the pneumonia versus non-pneumonia lower respiratory tract infection binary, which failed against the patient-city confounder baseline, plausibly reflecting a clinic-level difference in ICD-10 coding.

**Conclusion:** Conversational primary care audio carries acoustic signal that discriminates clinically meaningful respiratory contrasts. Absolute performance is moderate, but the conditions are stricter than prior work: conversational speech and differential-diagnosis contrasts among sick patients. This pilot study is a baseline for voice-based clinical AI moving beyond sick-versus-healthy detection toward differential-diagnosis panels and a proof-of-concept for hierarchical composition.

## 1 INTRODUCTION

In recent national primary care data, a lung- and breathing-related (respiratory) diagnosis was recorded at approximately 10.6 to 13.5% of visits to US health centers across biannual periods from January 2022 to December 2025, making respiratory illness a common condition in primary care (National Center for Health Statistics, 2025). Most of these visits are viral upper respiratory infections that resolve with supportive care alone. A meaningful fraction, however, are conditions where the choice of clinical action affects outcome: bacterial pneumonia warrants chest imaging and timely antibiotics, influenza and severe acute respiratory syndrome coronavirus 2 (SARS-CoV-2) warrant antiviral therapy and isolation guidance, Group A streptococcal pharyngitis warrants targeted antibiotic prescribing, and chronic respiratory disease flares warrant medication review and longitudinal management rather than acute treatment. Distinguishing among these presentations relies on clinical judgement, rapid diagnostic tests where available, and selective laboratory and imaging studies.

The consequences of triaging these visits incorrectly are well documented. Acute respiratory infections account for approximately 44% of outpatient antibiotic prescriptions, of which roughly half are unnecessary (Fleming-Dutra et al., 2016), and inappropriate ambulatory prescribing is a leading driver of community-level antimicrobial resistance (Centers for Disease Control and Prevention, 2019). Missed or delayed identification of pneumonia in primary care contributes to avoidable hospitalization (Singh et al., 2013). Influenza and coronavirus disease 2019 (COVID-19) cases that go unrecognized within the antiviral treatment window lose the opportunity for early therapy and household-exposure mitigation; the Infectious Diseases Society of America influenza guideline (Uyeki et al., 2019) and the EPIC-HR trial of nirmatrelvir-ritonavir (Hammond et al., 2022) each define a narrow window from symptom onset.

Voice is a candidate signal for respiratory triage because the speech production apparatus is mechanically coupled to the respiratory tract. Phonation requires airflow generated by the lungs, modulated by the larynx, and shaped by the resonant cavities of the pharynx, nasal cavity, and oral cavity. Every component of this chain is directly affected by the conditions a respiratory triage system must distinguish. Mucosal swelling in the upper airway shifts vocal tract resonances and articulation (Behrman, 2023). Nasal congestion alters the spectral balance of voiced segments through changed nasal coupling (de Boer and Bressmann, 2016). Inflammation of the pharynx and larynx produces dysphonia and altered phonation (Pimenta et al., 2022). Lower respiratory tract involvement, whether airway-only as in acute bronchitis or parenchymal as in pneumonia, reduces effective lung volume and changes breath support, with measurable effects on prosodic timing and phonation stability (Weglarz et al., 2025). Systemic features common to influenza, COVID-19, and other acute respiratory infections may produce additional voice changes through fatigue and reduced respiratory drive (Behrman, 2023). Each of these mechanisms acts on a distinct component of the speech production chain, which is why an acoustic system that examines a broad set of voice features simultaneously is, in principle, well suited to a panel of respiratory contrasts rather than to any single condition. Voice also has an operational advantage over many other candidate biomarkers in primary care: it is already present in every clinical encounter and, in the increasingly common setting of ambient-recorded visits, is already captured passively without changing the patient experience or adding to clinician workload (Olson et al., 2025; Shah et al., 2025).

A substantial body of work has investigated voice and cough as acoustic markers of respiratory disease, dominated since 2020 by efforts to detect COVID-19 from voluntary cough, sustained vowel, or read-speech recordings collected through smartphone apps and web platforms (Xia et al., 2021; Muguli et al., 2021). Adjacent work has applied similar acoustic methods to tuberculosis screening using clinic-recorded solicited cough (Huddart et al., 2024; Pahar et al., 2021a) and to single-condition vocal biomarker development for asthma, chronic obstructive pulmonary disease, and interstitial lung disease using brief sustained-vowel tasks (Kaur et al., 2023). These efforts have established that voice and cough carry detectable signal for the specific contrasts they study, typically against healthy control populations and typically using elicited rather than spontaneous speech. Three structural gaps remain. First, the contrasts that primary care triage actually requires are not single-condition detections against healthy controls but differentiations among sick patients: pneumonia versus bronchitis, infectious versus non-infectious, viral versus bacterial, influenza versus other acute infection. No published acoustic system, to our knowledge, has been evaluated across the panel of such differential-diagnosis contrasts that a primary care clinician faces in a single visit; the closest prior work (Porter et al., 2019a) evaluates solicited cough across five common pediatric respiratory disorders (N=585) in a clinical research setting. Second, almost all prior datasets in this space rely on prompted speech tasks performed under controlled or semi-controlled conditions, while the deployment scenario for any practical primary care triage tool is conversational ambient audio with the variability of real clinical encounters. Third, the scale at which prior conversational respiratory work has been conducted is small relative to the primary care population it would need to serve; the largest crowdsourced conversational and read-speech collections include tens of thousands of unique subjects (Xia et al., 2021; Bhattacharya et al., 2023), while the largest published clinical solicited-cough datasets include a few thousand (Huddart et al., 2024). Whether voice carries discriminating signal at the contrasts and conditions that primary care triage demands, at the scale at which primary care actually operates, has not been tested.

This paper investigates whether conversational primary care audio carries acoustic signal at clinically meaningful respiratory contrasts. The work is structured as a panel of 11 binary classification tasks chosen as a proof-of-concept set rather than as a complete triage system. Three criteria guided the selection. First, the contrast had to correspond to a clinically meaningful differential decision in primary care: respiratory versus non-respiratory illness at the gate, then progressively finer contrasts including acute versus chronic, infectious versus non-infectious, lower versus upper tract, pneumonia versus bronchitis, acute sinusitis versus other upper respiratory infection, Group A streptococcal versus other pharyngitis, influenza versus other acute infection, COVID-19 versus other acute infection, and bacterial versus viral overall. Second, the contrast had to be supported by sufficient labeled data from the source extract to yield train, development, and test partitions large enough for meaningful evaluation at the patient level. Third, each side of the contrast had to be biologically plausible as a voice-affecting condition, in the sense that documented mechanisms exist for how the underlying disease alters voice production. The 11 tasks correspond to nodes along a clinical decision cascade for respiratory triage.

The contributions of this paper are as follows:

1. The first evaluation of voice-based respiratory triage at population scale on conversational primary care audio. Approximately 514,000 visits from 379,000 unique adult patients drawn from a US primary care clinic network spanning 50 states and over two years, with no scripting, no prompted speech tasks, and no change to the patient or clinician workflow. Each visit carries the clinically assigned ICD-10 diagnosis codes from the encounter as the labeling source. The dataset is not publicly released, but the cohort definitions, modeling pipeline, and evaluation protocol are described in full to support replication on comparable data.
2. A panel of 11 binary classification tasks operationalizing a respiratory triage cascade. Each task is defined by ICD-10-based (International Classification of Diseases, 10th Revision) inclusion and exclusion rules and selected by criteria of clinical meaningfulness, data sufficiency, and biological plausibility, spanning the gate (respiratory vs non-respiratory), acute vs chronic, infectious vs non-infectious mimic, anatomical (lower vs upper), pneumonia and bronchitis contrasts, upper-tract antibiotic decisions, and pathogen-specific differentiations for influenza, SARS-CoV-2, Group A streptococcus, and bacterial vs viral etiology.
3. Per-task evaluation on patient-stratified held-out test sets. Each of the 11 classifiers is trained, hyperparameter-selected, and evaluated independently using five-fold cross-validation and a held-out 30% of unique patients, with discrimination, calibration, and threshold-dependent metrics reported.
4. Panel-level confounder analysis. For each binary, six baselines along demographic, geographic, and temporal axes are compared against the acoustic model on the same held-out test set; comparisons are corrected for multiple testing across the 11-binary panel, and we report which binaries’ discrimination is genuinely attributable to voice and which is partly or fully explained by an axis-specific prior.
5. Illustrative cascade case studies. Selected test-set patients are routed through the 11 trained classifiers in the order of the cascade to demonstrate end-to-end traversal on real clinical encounters; we report a small set of successful traversals as case studies, not as a validated end-to-end triage system.

The remainder of this paper is organized as follows: Section 2.1 describes the primary care audio dataset, its demographic, geographic, and clinical distribution, and its comparison with prior datasets in the field. Section 2.2 formalizes the panel of 11 binary classification tasks, including the ICD-10-based cohort definitions, clinical motivation, and biological grounding for each task. Section 2.3 describes the audio preprocessing, feature extraction, model architecture, training protocol, evaluation metrics, and confounder analysis. Section 3 reports per-task discrimination, calibration, and threshold-dependent results, together with the panel-level confounder analysis. Section 4 discusses the implications, limitations, and open questions raised by these results, and Section 5 concludes.

## 2 MATERIALS AND METHODS

The end-to-end methods pipeline is shown in Figure 1. This section describes the dataset (Section 2.1), the panel of 11 binary classification tasks (Section 2.2), and the modeling and evaluation protocol (Section 2.3).

**Figure 1.**
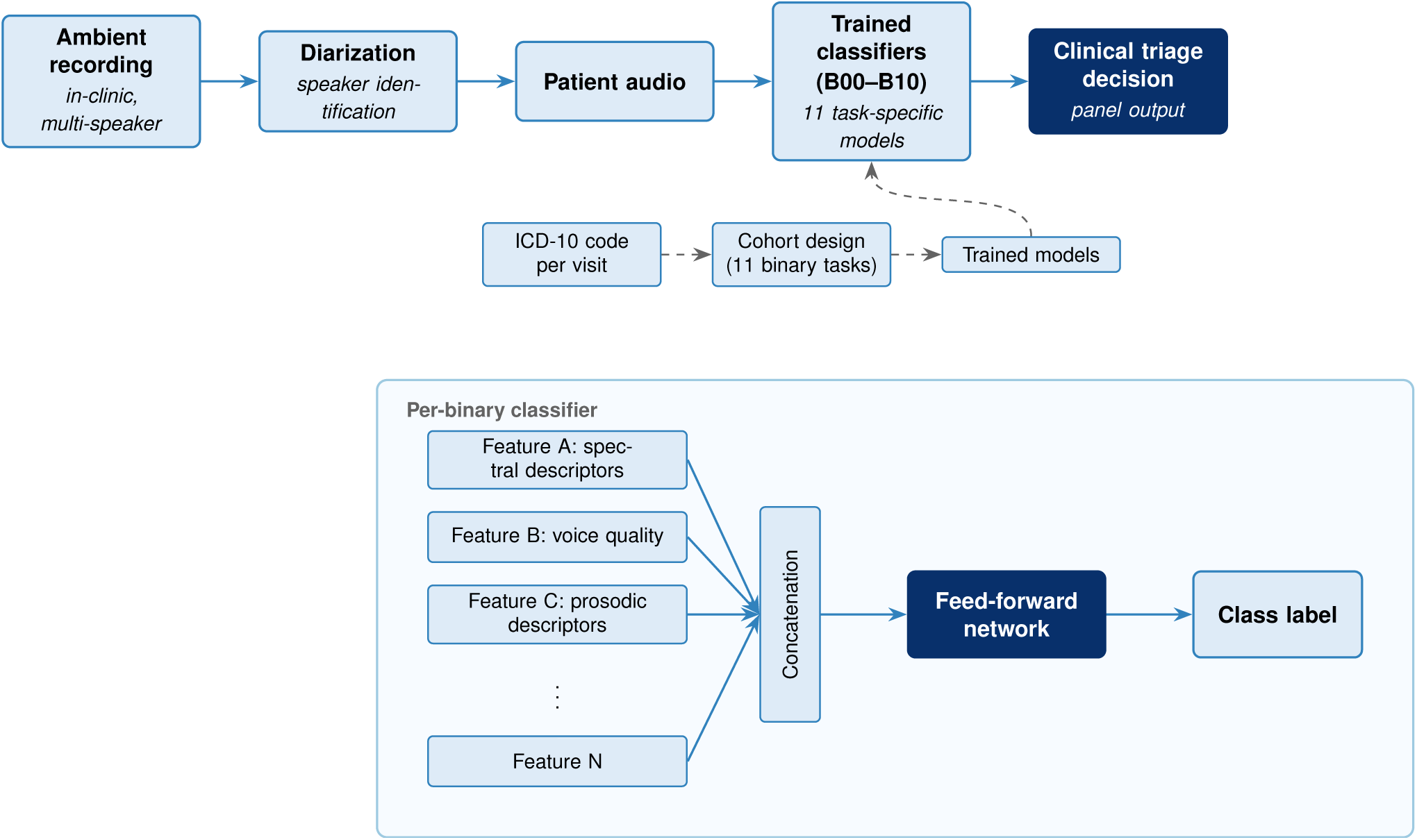
End-to-end methods pipeline. The full-visit audio is diarized to extract a patient-only channel, which is processed through nine acoustic feature streams to produce a 412-dimensional feature vector per visit. The feature vector is the input to a per-binary feed-forward classifier, trained independently for each of the 11 classification tasks under patient-stratified five-fold cross-validation. Each trained classifier is evaluated on a held-out test set, compared against six non-acoustic confounder baselines, and composed into a hierarchical screening cascade.

### 2.1 Dataset

#### 2.1.1 Overview

The dataset comprises ambient-scribed audio of routine adult primary care visits, that is, in-room recordings of natural conversations between patients and clinicians captured by an ambient clinical-documentation system, with no headworn microphone, no scripted protocol, and no prompted speech task. The recordings come from a US primary care clinic network operating across multiple sites. Audio was collected during the ordinary course of clinical care, and the resulting acoustic data reflect the variability any deployed triage system would encounter at the point of care: background noise, overlapping speech, interruptions, accents, conversational disfluencies, and the full range of patient affect and presentation severity seen in primary care. Secondary research use of the recordings was disclosed to patients through the originating network’s Notice of Privacy Practices, under which patients could opt out of research participation; patients who opted out were excluded, and identifying information was removed from the visit metadata before the data were made available for analysis. This work is a secondary analysis of those recordings, conducted under the data-use terms of the originating network and under an IRB exempt determination with an approved waiver of HIPAA authorization (see Ethics Statement). The source extract for this work covers 1,734,936 visits from 1,008,836 unique adult patients, with recording dates spanning 2024-01-01 to 2026-02-18, a window of 779 days. Each visit corresponds to a single audio file of the full patient-clinician encounter, and the analyses in this paper use only the patient-speech portion of each recording, isolated through the diarization pipeline described in Section 2.3.1.

Patients under 18 years of age at the time of the encounter were excluded prior to source-extract construction; the visit and patient counts reported above are after this pediatric exclusion. A two-stage audio-quality filter was then applied to the source extract. The first stage operated on the raw recording before diarization. A visit was retained if its audio duration fell between 40 seconds and one hour, and was excluded otherwise. Recordings shorter than 40 seconds were treated as misfires, dropped sessions, or test files, and recordings longer than one hour were treated as cases where the recording device was left running past the end of the visit, captured an unrelated procedure, or reflected a system error. Recordings that could not be diarized at all (for example, due to file corruption, unsupported format, or pipeline failure) were excluded at this stage as well, and were not carried forward to feature extraction. The second stage operated on the diarized output. After the diarizer produced speaker-segmented audio, the total duration of speech assigned to the patient channel was required to meet a nominal one-second floor. Visits failing this second stage included those where only the clinician spoke and no patient channel was identifiable, those where the diarized output was empty, and those where total patient speech fell below the one-second floor; the threshold was set on the assumption that any shorter segment is dominated by background noise, cross-talk artifact, or transient utterance and carries no analyzable acoustic content. A small fraction of visits fall just below this nominal floor owing to a minor leak in the filter (3,542 visits, 0.7% of the analytic sample, reported in the supplementary material), too small to affect the downstream analysis. After both stages, 1,418,206 visits (81.7% of the source extract) were retained and 316,730 visits (18.3%) were excluded. Per-stage and per-reason counts within the excluded group were not retained during processing and are not reported here; we acknowledge this as a limitation, since we cannot stratify the exclusion to test for selective loss by site, demographic, or condition. No other audio-quality filters were applied: sample rate, codec, signal-to-noise ratio, and silence ratio were not used as exclusion criteria, and visits surviving both stages entered the downstream analysis at their native recording quality.

After the audio-quality filter, the analyzable set consists of 1,418,206 visits from 843,069 unique patients. Each visit in this set is represented as a single row carrying the visit-level metadata used throughout this work. This metadata comprises the primary ICD-10 diagnosis code assigned by the treating clinician for the encounter, the full array of ICD-10 codes recorded during the encounter, patient age, patient sex, the patient’s city and state of residence, the date and time of service, and a reference to the patient-speech audio extracted from the recording. Patient identifiers were retained in pseudonymized form so that visits could be grouped by patient for longitudinal and patient-level analyses, but no name, address, contact information, or other direct identifier was carried through to the analytic table. The work reported in this paper does not use the full 1,418,206-visit set. Downstream analyses operate on a subset of 514,377 visits from 379,225 unique patients, defined by the inclusion and exclusion rules of the classification tasks described in Section 2.2. All distributions reported in the following subsections (demographic, geographic, clinical, and audio duration) describe this 514,377-visit subset, referred to throughout as the analytic sample. The full visit, patient, and class composition of each individual classification task is reported in Section 2.2.2.

The analytic sample is an adult population (all patients at least 18 years of age at the visit): median age is 38 years (interquartile range 28 to 55, range 18 to 106) and 63.5% of visits are female. It spans all 50 US states, the District of Columbia, and four US territories, with California the largest single state (58.4% of visits) and the top five states accounting for 86.9%. Patient-channel audio has a median duration of 27.8 seconds (interquartile range 15.4 to 47.2) and totals 5,258 hours. The full demographic (age, sex, longitudinal visit structure), geographic (state, city), clinical (ICD-10 chapter and primary-code distributions, documented comorbidities), and audio-duration distributions of the analytic sample are reported in the supplementary material.

#### 2.1.2 Comparison with prior voice and cough datasets in respiratory research

Voice and cough datasets in published respiratory research span three regimes: crowdsourced web or app collections that ask subjects to perform scripted tasks remotely, single-site clinical collections that record voluntary cough or scripted speech under controlled conditions, and a small number of mixed-mode collections that combine the two. Table 1 compares the present dataset against the principal datasets used in this literature. Two patterns are immediately visible. Almost every prior dataset targets a single condition, with COVID-19 dominating the crowdsourced regime (Coswara (Bhattacharya et al., 2023), COUGHVID (Orlandic et al., 2021), COVID-19 Sounds (Xia et al., 2021), DiCOVA-1 (Muguli et al., 2021), DiCOVA-2 (Sharma et al., 2022), Virufy (Chaudhari et al., 2020), NoCoCoDa (Cohen-McFarlane et al., 2020)) and tuberculosis dominating the clinical-cough regime (Botha (Botha et al., 2018), Pahar (Pahar et al., 2021a), CODA TB DREAM (Huddart et al., 2024), TBscreen (Sharma et al., 2024)). The Sonde Health respiratory-responsive vocal biomarker work is the closest prior multi-condition effort and covers asthma, chronic obstructive pulmonary disease, interstitial lung disease, and COVID-19 across roughly 5,000 subjects in its respiratory-impairment validation cohorts, with COVID-19 evaluated in a separate case-control study (Kaur et al., 2023), but uses a single scripted task (a six-second sustained vowel). The second pattern is on speech naturalness: every dataset in Table 1 except the present work uses scripted elicitations (cough on demand, sustained vowels, counting, or read sentences), or in the case of NoCoCoDa, cough events extracted from media interviews. No prior public or institutionally documented dataset in this space, to our knowledge, consists of conversational primary care visits.

**Table 1.** Voice and cough datasets used for respiratory disease research in the published literature, compared with the present work. n/r = not reported in primary source.

| Dataset | Year | Subjects | Recordings | Recording task | Setting | Conditions | Public |
| --- | --- | --- | --- | --- | --- | --- | --- |
| Botha TB cough (Botha et al., 2018) | 2018 | 38 | n/r | Voluntary cough | Clinic | TB | No |
| Coswara (Bhattacharya et al., 2023) | 2020 | 2,635 | 23,715 | Vowel, cough, breath, count | Crowdsourced web/app | COVID-19 | Yes |
| COUGHVID (Orlandic et al., 2021) | 2021 | n/r | >25,000 | Voluntary cough | Crowdsourced web/app | COVID-19 | Yes |
| COVID-19 Sounds, Cambridge (Xia et al., 2021) | 2021 | 36,116 | 53,449 | Read sentence, cough, breath | Crowdsourced web/app | COVID-19, asthma | On request |
| NoCoCoDa (Cohen-McFarlane et al., 2020) | 2020 | 10 | 73 | Cough from media interviews | Media (no participation) | COVID-19 | On request |
| Virufy open cough (Chaudhari et al., 2020) | 2020 | 16 | 121 | Voluntary cough | Clinic + smartphone | COVID-19 | Yes |
| DiCOVA-1 (Muguli et al., 2021) | 2021 | 1,040 | n/r | Cough, breath, vowel, count | Crowdsourced web/app | COVID-19 | Yes |
| Pahar TB cough (Pahar et al., 2021a) | 2021 | 51 | 1,358 | Voluntary cough, count, breath | Clinic | TB | No |
| DiCOVA-2 (Sharma et al., 2022) | 2022 | 1,436 | n/r | Cough, count, breath | Crowdsourced web/app | COVID-19 | Yes |
| Sonde RRVB (Kaur et al., 2023) | 2023 | ~5,000 | n/r | Sustained vowel | Mixed (smartphone) | Asthma, COPD, ILD, COVID-19 | No |
| CODA TB DREAM (Huddart et al., 2024) | 2024 | 2,143 | 733,756 coughs | Solicited cough x 5 | Clinic | TB | On request |
| TBscreen (Sharma et al., 2024) | 2024 | 195 | 34,600 coughs | Passive + forced cough | Clinic | TB, other respiratory | Yes |
| **This work** | 2026 | 379,225 | 514,377 visits | Conversational primary care visit | Clinic (ambient) | 11 respiratory contrasts (panel) | No |

*Notes.* Recordings counts mix visit-level recordings and per-event extractions across datasets and are not directly comparable in isolation; they should be read alongside recording task and setting. AICovidVN 115M, ComParE CCS, and ComParE CSS are excluded because they are derived from or are subsets of datasets already listed.

On scale, the comparison is also informative. The largest crowdsourced collection by unique subjects is COVID-19 Sounds at 36,116 participants (Xia et al., 2021); the present work covers 379,225 unique patients, an order of magnitude larger. The largest clinical-cough collection by total recording count is CODA TB DREAM at 733,756 cough events (Huddart et al., 2024), but the cough events are short solicited bursts (five coughs per subject in the training set, with a 565-subject sub-cohort contributing two weeks of longitudinal community-setting coughs), drawn from 2,143 subjects. The present work yields 514,377 patient-clinician conversational recordings averaging 36.8 seconds of patient speech per visit (median 27.8 seconds), totaling 5,258 hours of conversational acoustic content. The dataset is therefore positioned at the unoccupied intersection of three properties: large scale (order of magnitude above prior unique-subject counts), conversational and unscripted speech (not present in any prior public dataset), and clinical breadth across the 11 respiratory contrasts of the triage panel rather than a single condition. This positioning is the empirical basis on which the analyses in the rest of this paper rest.

### 2.2 Classification task design

The acoustic respiratory triage system is decomposed into 11 binary classification tasks, indexed B00 through B10, each defined by a positive and a negative cohort constructed by ICD-10-based inclusion and exclusion rules applied to the post-audio set described in Section 2.1. The 11 tasks were chosen to populate clinically meaningful decision points along the triage cascade shown in Figure 2: respiratory versus non-respiratory at the gate, then progressively finer contrasts including acute versus chronic, infectious versus non-infectious, lower versus upper tract, pneumonia versus bronchitis, and specific pathogen contrasts for influenza, SARS-CoV-2, and Group A streptococcus. Several of these contrasts have not been characterized acoustically in conversational primary care speech to our knowledge, and the panel is designed as a test of whether voice carries discriminating signal at these contrasts, not as a confirmatory analysis of established acoustic phenotypes. Each binary is evaluated independently in this work. The 11 tasks are summarized below; each entry names the positive cohort, the negative cohort, and the biological motivation supporting the contrast. Full ICD-10 inclusion and exclusion logic, comorbidity-based exclusions, date-aware washouts, the seven patient-level covariate flags carried through every cohort, and the supporting literature for the biological motivation are provided in the supplementary material.

**Figure 2.**
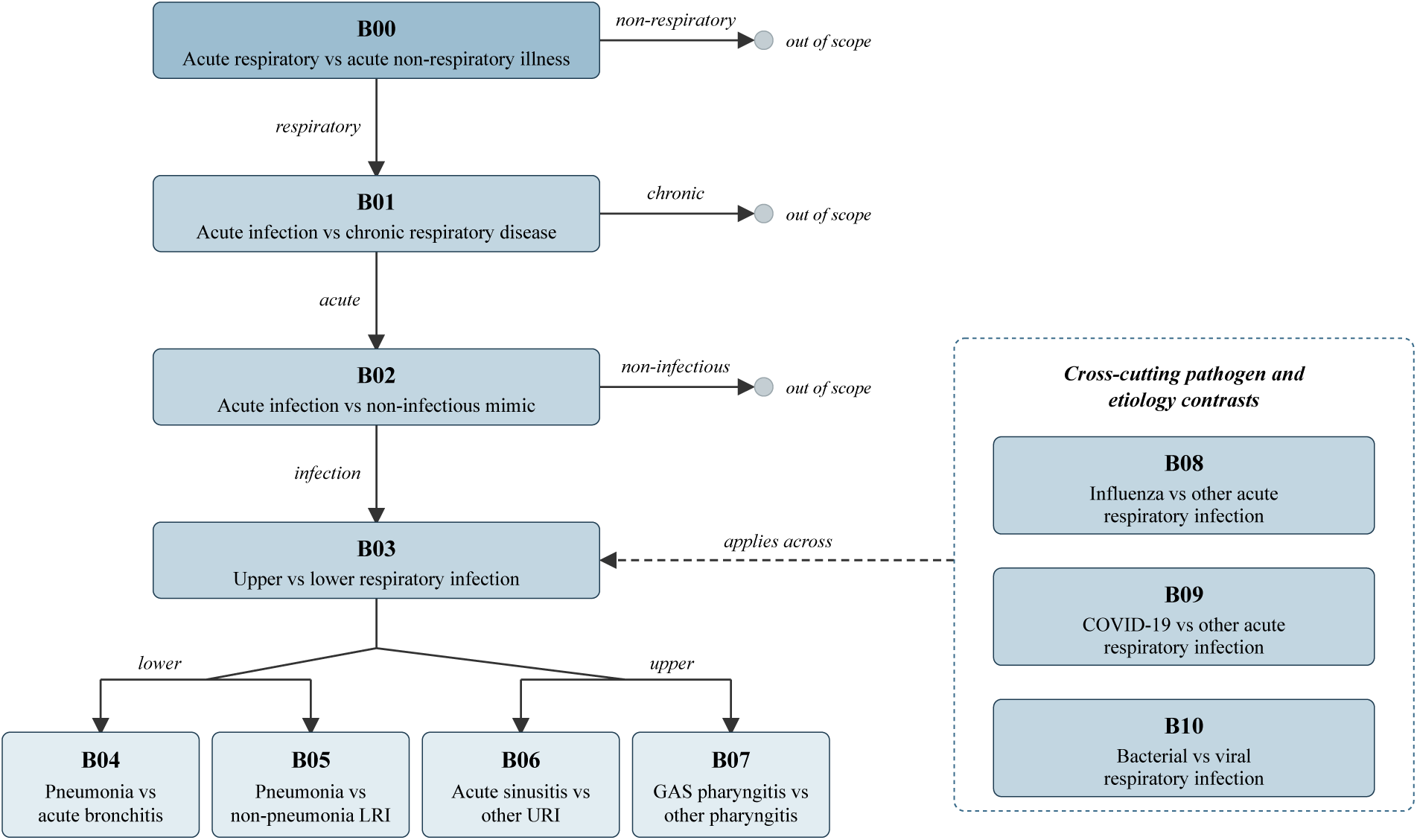
Respiratory triage cascade. The 11 binary classification tasks (B00 through B10) populate decision nodes along the cascade, beginning at the respiratory-versus-non-respiratory gate (B00) and progressively narrowing through acute-versus-chronic, infectious-versus-non-infectious, anatomical (lower versus upper tract), pneumonia and bronchitis contrasts, upper-tract antibiotic decisions, and pathogen-specific differentiations for influenza, SARS-CoV-2, Group A streptococcus, and bacterial versus viral etiology.

#### 2.2.1 Task specifications

B00: Acute respiratory versus acute non-respiratory illness. The positive cohort is visits with a primary diagnosis of acute respiratory infection. The negative cohort is visits with an acute non-respiratory primary diagnosis (gastrointestinal, genitourinary, musculoskeletal, headache, skin, mental health, or non-respiratory symptom). Acute respiratory illness alters voice production through upper-airway mucosal swelling, changed nasal coupling, and altered breath support and prosodic timing (Sara et al., 2023; de Boer and Bressmann, 2016; Pimenta et al., 2022; Weglarz et al., 2025).

B01: Acute respiratory infection versus chronic respiratory disease management. The positive cohort is visits coded for acute respiratory infection. The negative cohort is visits coded for chronic respiratory disease (COPD, asthma, bronchiectasis, interstitial pulmonary disease, cystic fibrosis, and related entities). Acute infection produces transient upper-airway changes while chronic disease produces persistent changes tied to lower-airway acoustics, breath support, and subglottal pressure (Lyu et al., 2025; Yan et al., 2025; Weglarz et al., 2025).

B02: Acute respiratory infection versus symptomatic non-infectious respiratory presentation. The positive cohort is visits coded for acute respiratory infection. The negative cohort is visits coded for symptomatic non-infectious respiratory presentations (allergic rhinitis, chronic rhinosinusitis, cough, dyspnea, wheeze, and throat pain). The two states converge on similar surface symptoms but may diverge in acoustic signature: infection produces transient mucosal edema and secretion-driven changes, while non-infectious presentations such as allergic rhinitis themselves measurably alter voice (Koç et al., 2014).

B03: Lower versus upper respiratory infection. The positive cohort is visits coded for lower respiratory infection (pneumonia, acute bronchitis, bronchiolitis, unspecified acute LRI, and pneumonia sequelae). The negative cohort is visits coded for upper respiratory infection (common cold, sinusitis, pharyngitis, tonsillitis, and unspecified URI). The contrast is between inflammation that primarily affects vocal tract resonance and articulation (upper tract) and inflammation that primarily affects breath support, cough character, and gas exchange (lower tract) (de Boer and Bressmann, 2016; Weglarz et al., 2025).

B04: Pneumonia versus acute bronchitis. The positive cohort is visits coded for pneumonia (J12–J18). The negative cohort is visits coded for acute bronchitis (J20). Pneumonia involves alveolar consolidation that alters acoustic transmission through the chest cavity and changes breath support during phonation, while bronchitis involves airway-level inflammation with a more localized acoustic signature; cough-acoustic analysis has discriminated pneumonia from bronchitis in pediatric cohorts (Liao et al., 2022; Porter et al., 2019b; Sharan et al., 2024), though to our knowledge not from conversational adult speech.

B05: Pneumonia versus non-pneumonia lower respiratory infection. The positive cohort is visits coded for pneumonia. The negative cohort is visits coded for acute bronchitis, bronchiolitis, or unspecified acute LRI (J22). The contrast is the same alveolar-versus-airway distinction as B04, but the J22 unspecified code in the negative arm introduces label noise: J22 is assigned in practice when a lower respiratory infection is documented without committing to pneumonia or bronchitis.

B06: Acute sinusitis versus other upper respiratory infection. The positive cohort is visits coded for acute sinusitis (J01). The negative cohort is visits coded for other upper respiratory infection (common cold, pharyngitis, tonsillitis, acute laryngitis and tracheitis, croup and epiglottitis, and unspecified URI). Sinusitis involves paranasal inflammation that changes the resonant properties of the nasal and paranasal cavities, producing measurable hyponasality; other URIs produce mucosal inflammation distributed across the pharynx, larynx, and nasal cavity without the focal paranasal involvement (Jiang and Huang, 2006).

B07: Group A streptococcal pharyngitis versus other pharyngitis. The positive cohort is visits coded for streptococcal pharyngitis (J02.0). The negative cohort is visits coded for other specified or unspecified pharyngitis (J02.8, J02.9). GAS pharyngitis produces intense tonsillar and pharyngeal inflammation with exudate and severe odynophagia, plausibly altering oropharyngeal resonance, articulation of posterior consonants, and prosody through compensatory adjustments to minimize pain (Finkelstein et al., 1993).

B08: Influenza versus other acute respiratory infection. The positive cohort is visits coded for influenza (J09, J10, J11). The negative cohort is visits coded for other acute respiratory infection (URI, LRI, or COVID-19). Influenza is distinguished from other acute respiratory infections less by its respiratory manifestations than by its systemic prodrome (fever, myalgia, prostration, fatigue), which may plausibly produce voice changes through fatigue and reduced respiratory drive (Behrman, 2023).

B09: COVID-19 versus other acute respiratory infection. The positive cohort is visits coded for confirmed COVID-19 (U07.1, J12.82). The negative cohort is visits coded for other acute respiratory infection (URI, LRI excluding COVID, and influenza). COVID-19 produces a respiratory illness whose acoustic signature has been characterized in prior work on scripted speech (Brown et al., 2020; Shimon et al., 2021; Pahar et al., 2021b); the present binary tests whether the contrast holds on conversational speech and during the Omicron-dominant era of the source data.

B10: Bacterial versus viral respiratory infection. The positive cohort is visits coded for bacterial respiratory infection (identified-organism bacterial pneumonia, streptococcal pharyngitis and tonsillitis, whooping cough, pulmonary tuberculosis). The negative cohort is visits coded for viral respiratory infection (identified-organism viral pneumonia excluding COVID, common cold, influenza). The two etiologies may plausibly produce systematically different patterns of mucosal involvement, secretion character, breath support, and systemic prodrome, though the contrast is acoustically subtle. While cough character (wet versus dry) has been used as a proxy for bacterial versus viral infection (Renjini et al., 2021), direct acoustic discrimination of bacterial from viral respiratory infection, validated against a clinical reference standard in adults, remains to our knowledge unaddressed.

#### 2.2.2 Per-task cohort sizes

The source-pool cohort sizes for each of the 11 binary classification tasks are shown in Figure 3. Cohort sizes range from approximately 10,000 to 280,000 visits per class, and within each binary the positive and negative cohorts differ in size by factors that range from near unity (B04, 1.24 to 1) to over 20 to 1 in the most imbalanced binaries (B01 at 24.3 to 1, B08 at 1 to 20.3, B09 at 1 to 16.1). These ratios reflect the natural prevalence of each clinical contrast in the source primary care population rather than any experimental choice; for example, influenza and COVID-19 confirmed primary diagnoses appear in roughly 1 of every 20 (influenza) and 1 of every 16 (COVID-19) acute respiratory visits in the source extract, which is the structural reason B08 and B09 are highly imbalanced. Class balancing is applied prior to model training and is described in Section 2.3.2. Summary statistics for age, sex, geography, and audio duration stratified by cohort and class are reported in the supplementary material.

**Figure 3.**
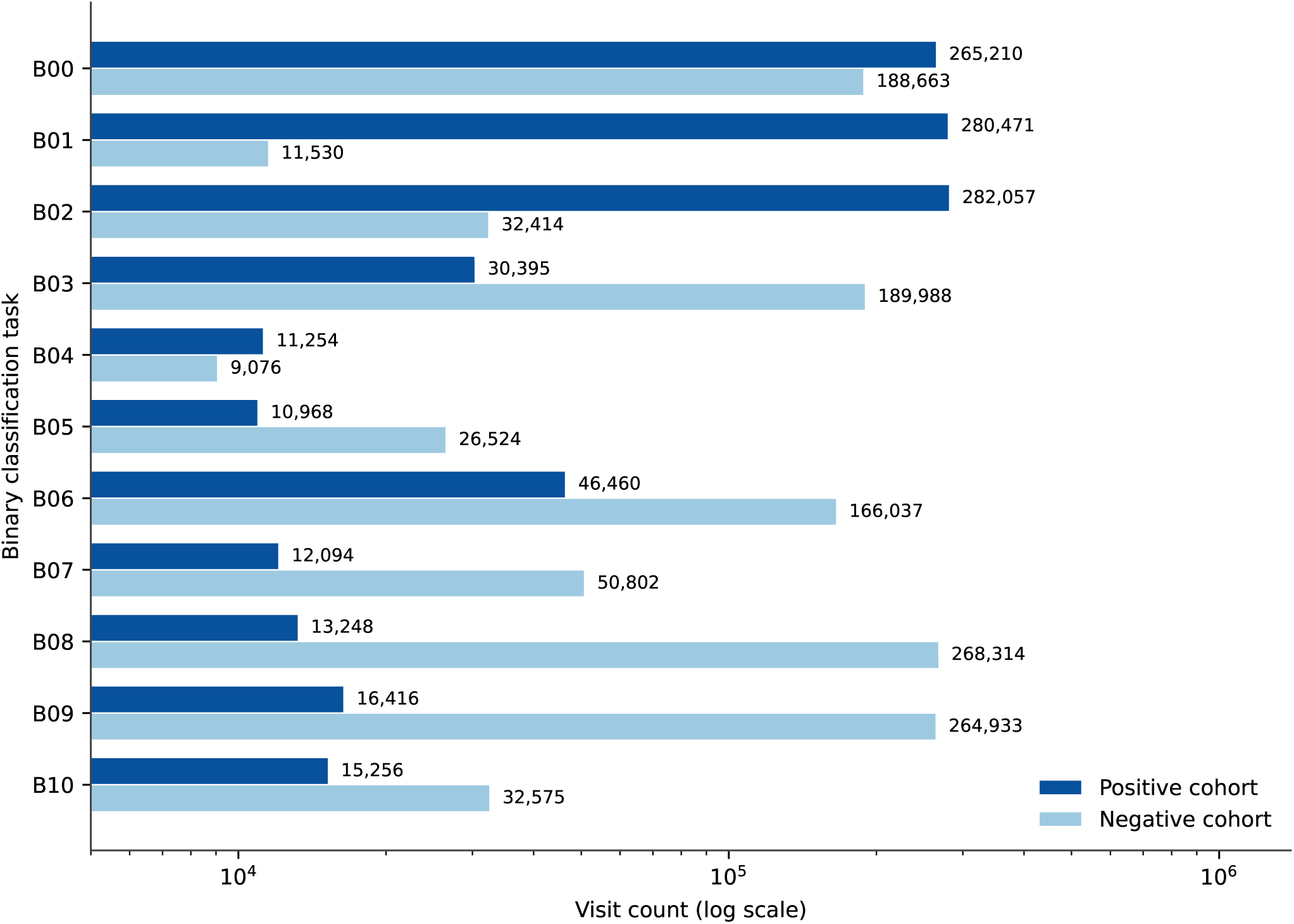
Per-task cohort sizes for the 11 binary classification tasks. Each task has a positive (label=1) and negative (label=0) cohort; sizes range from approximately 10,000 to 280,000 visits per class. Imbalance ratios reflect the natural prevalence of each clinical contrast in the source population, from near unity (B04, 1.24:1) to over 20:1 in the most imbalanced binaries (B01 at 24.3:1, B08 at 1:20.3, B09 at 1:16.1).

### 2.3 Modeling and Evaluation

Each visit contributes one labeled example per binary: the label is determined by the visit’s primary ICD-10 code under the cohort rules of Section 2.2, and the input is the 412-dimensional acoustic feature vector computed from that visit’s patient speech (Section 2.3.1). The models are trained to map the feature vector to the binary label. The processing pipeline for each visit proceeds in five stages: audio preprocessing, feature extraction, model training, evaluation, and panel-level confounder analysis. We deliberately use established open-source feature sets (openSMILE eGeMAPSv02, librosa) and a lightweight feed-forward classifier (approximately 60,000 to 380,000 parameters per binary depending on the selected topology) as the modeling baseline for this study. Representation learning with larger acoustic models is a separate, ongoing line of work in our group, but is out of scope here to keep the present results reproducible from publicly available extractors. The 11 trained classifiers are then composed into a hierarchical screening cascade. The end-to-end pipeline is shown in Figure 1. The subsections below summarize each stage at the level of the main methodological claims; per-stream feature definitions, exact filter thresholds, hyperparameter ranges, and per-binary configurations are reported in the supplementary material.

#### 2.3.1 Audio Processing and Feature Extraction

The audio entering the modeling pipeline is the patient-only channel extracted from each visit recording by an upstream diarization pipeline. The full-recording audio is submitted to Google Cloud Speech-to-Text v2 using the Chirp 3 acoustic model, and the diarized transcript is passed to the gemini-flash-3.1-lite large language model to identify which speaker is the patient; patient-only audio is then reconstructed by concatenating the time-aligned slices of the recording corresponding to the identified patient channel. The speaker diarization is configured for one to two speakers with English recognition and word-level timestamps, and consecutive same-speaker words are merged into segments. The language model is given the diarized transcript and asked to return the speaker label of the patient, defined as the primary subject of the visit rather than a caregiver, translator, or other participant; when it returns a sentinel indicating that no patient is present, the visit is excluded. The two audio-quality gates described in Section 2.1.1 are applied during this pipeline. Full diarization parameters are reported in the supplementary material.

The resulting patient WAV (16 kHz mono) is passed to a fixed feature-extraction pipeline that produces a single 412-dimensional summary vector per visit. The vector is organized into nine streams drawn from four extractor families: openSMILE eGeMAPSv02 (six streams partitioned by speech-production component: pitch, loudness, spectral, voice quality, formants, and temporal) (Eyben et al., 2016, 2010), librosa (one stream of low-level spectral, chromagram, MFCC, energy, and rhythm features) (McFee et al., 2015), a valence-arousal-dominance predictor (one stream from WavLM-VAD-Odyssey2024) (Goncalves et al., 2024), and a temporal-analysis extractor (one stream from praat-parselmouth syllable-and-pause detection and librosa tempo statistics). No loudness normalization, denoising, or additional voice-activity detection is applied. Features in each stream are standardized within each cross-validation fold using a z-score transformation fitted on the training partition only, and the standardized streams are concatenated into a single 412-dimensional input vector. Per-stream feature counts, extractor configurations, the specific eGeMAPSv02 functionals, the librosa frame parameters, and the consolidation drop-rate are reported in the supplementary material.

#### 2.3.2 Model, Training and Evaluation

For each of the 11 classification tasks an independent binary classifier is trained, yielding 11 separate models. Each model is a fully connected feed-forward network whose input is the 412-dimensional concatenated feature vector and whose output is a single logit, trained under binary cross-entropy with logits (Figure 4). Each hidden block is a sequence of a linear layer, a one-dimensional batch normalization, a GELU activation, and a dropout layer with rate 0.5. The depth (two to four hidden blocks) and per-block widths are selected per binary by hyperparameter sweep. For each binary, the cohort is balanced at the patient level to twice the size of the minority class; 30% of unique patients are then held out as the test set, and the remaining 70% are partitioned by stratified five-fold cross-validation, also at the patient level. Each model is trained with AdamW (Loshchilov and Hutter, 2019) under a cosine-annealing learning-rate schedule (Loshchilov and Hutter, 2017) and early stopping on development loss. For each binary classifier, a grid search over four learning rates and four architectures is conducted on a held-out development set, and the best hyperparameter combination is then used to retrain the model under five-fold cross-validation; the fold-specific model with the lowest development loss is selected as the final classifier for that binary. A decision threshold is selected per binary by maximizing Youden’s J statistic (Youden, 1950) on the development partition of the selected fold.

**Figure 4.**
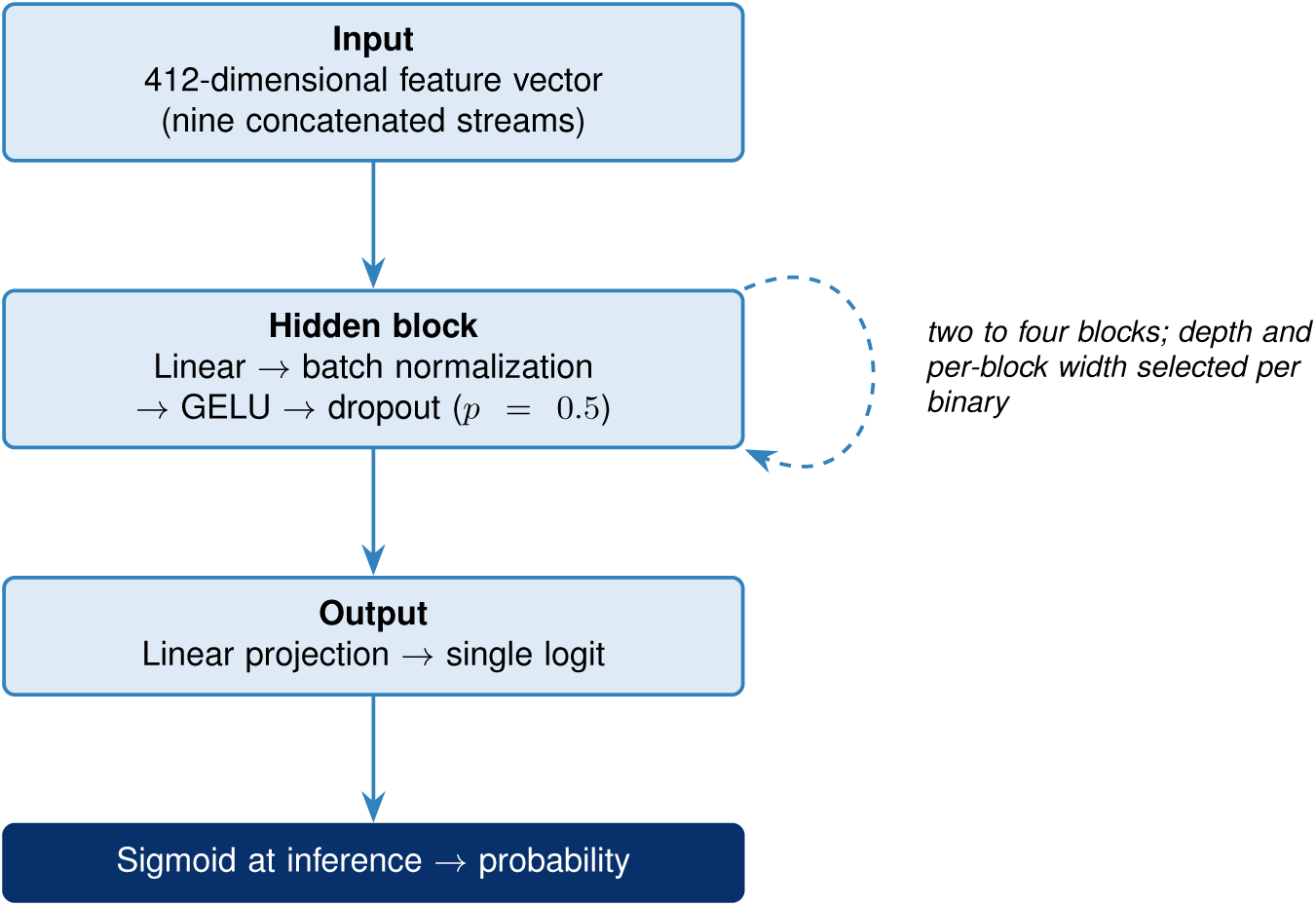
Per-binary feed-forward classifier. The 412-dimensional concatenated feature vector is passed through two to four hidden blocks, each a linear layer followed by one-dimensional batch normalization, a GELU activation, and dropout with rate 0.5, and then a linear projection to a single logit. A sigmoid is applied at inference to produce a probability. The depth and per-block widths are selected per binary by hyperparameter sweep.

Each final classifier is evaluated on its held-out test set using a panel of threshold-independent and threshold-dependent metrics. The primary metric is the area under the receiver operating characteristic curve (AUC), reported with a 95% confidence interval (CI) obtained from a 1000-iteration percentile bootstrap of the test predictions. The area under the precision-recall curve (AUPRC) is reported as a secondary discrimination metric. Calibration is assessed using the expected calibration error (ECE), reported under both equal-width and equal-frequency binning at 10 bins each, alongside a reliability diagram per binary. Threshold-dependent metrics (sensitivity/recall, specificity, precision, and F1 score) are reported at the per-binary decision threshold. Per-binary topology and learning rate, the full hyperparameter grid, and exact training-time hyperparameters (weight decay, batch size, schedule details, early-stopping patience, maximum epochs, gradient clipping) are reported in the supplementary material.

The full per-binary procedure is given in Algorithm 1.

The value reported for each binary is the AUC of the single selected fold (the fold with the lowest development loss) evaluated once on the frozen test set, not an average over the five cross-validation folds; cross-validation is used for model selection, and the test set is scored only once.

Hyperparameters (learning rate and architecture) are selected per binary in an outer sweep on a single training and development split that reuses the same held-out test set, so the test set is never seen during hyperparameter search. This procedure is leakage-safe but is not a fully nested cross-validation. Each binary is a separate model: a patient held out in the test set of one binary may appear in the training set of a different binary’s model, which does not introduce leakage, because each model excludes its own test patients from its own training set and no reported per-task metric combines models.

#### 2.3.3 Confounder analysis

The 412-dimensional acoustic feature vector encodes properties of patient speech, but it is also computed from a recording that is situated in time, geography, and demographic context. A model that achieves above-chance test discrimination could in principle be exploiting a demographic prior (older patients are more likely to be in the lower-respiratory-infection arm), a geographic prior (some clinic catchment areas have systematically different diagnostic distributions), or a temporal prior (influenza is concentrated in winter months) rather than the acoustic signal we intend it to learn. To test whether each binary’s test-set discrimination is genuinely attributable to voice rather than to one of these alternative explanations, we constructed a panel of six confounder baselines per binary, compared each baseline against the corresponding final model on the same held-out test set, and applied panel-level multiple-testing correction to the resulting per-binary comparisons.

The six baseline axes are: patient age (binned into 10-year intervals), patient sex, the joint of age bin and sex, patient city of residence, the joint of patient state and city, and the calendar month of the visit. Each baseline is a non-parametric per-bucket prevalence lookup. For each axis, the training partition (the same partition used to train the final model for that binary) is grouped by the axis key, the mean of the label within each bucket is computed, and that mean is used as the predicted probability for any test-set row whose key falls in the corresponding bucket. The age-sex and state-city baselines use chained fallback

##### Algorithm 1 Per-binary preparation, training, and evaluation (binary *B*).

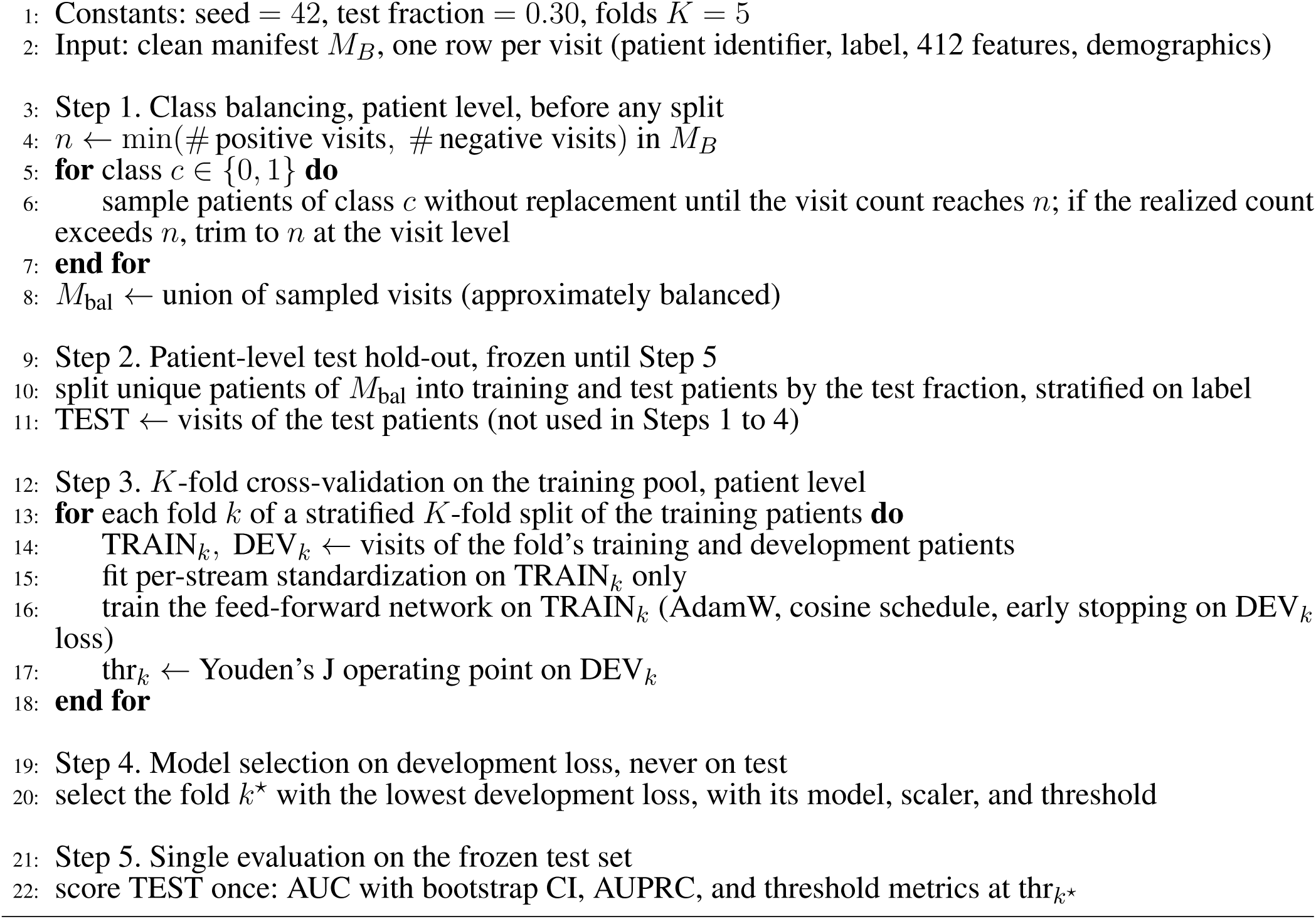

lookups when a test-set key is not present at full granularity (for example, the age-sex baseline falls back to the age-only mean, then to the sex-only mean, then to the global training-set prevalence). Test rows with missing values in the axis key are excluded from both the baseline and the paired comparison for that axis. For each (binary, axis) pair, the model and the baseline are scored on the same axis-filtered test rows, and the difference in AUC is tested by a paired bootstrap of 1,000 resamples. A one-sided test asks whether the model outperforms the baseline. Across the 11-binary panel, the resulting *p*-values for each axis are pooled and adjusted by the Benjamini-Hochberg (BH) procedure (Benjamini and Hochberg, 1995) at a false discovery rate (FDR) of 0.05. A binary passes the confounder check on an axis if its BH-adjusted *q*-value is at most 0.05; each binary therefore receives six pass-or-fail outcomes.

#### 2.3.4 Hierarchical screening cascade

The 11 binary classifiers were trained and evaluated independently. They are composed into a hierarchical screening cascade that mirrors the clinical decision sequence a primary care clinician would follow when evaluating a respiratory complaint (Figure 2).

A visit enters at B00 (acute respiratory versus acute non-respiratory illness). A respiratory classification routes the visit to B01 (acute infection versus chronic respiratory disease management), then to B02 (infection versus non-infectious mimic), then to B03 (lower versus upper respiratory infection). The lower-respiratory branch applies B04 (pneumonia versus acute bronchitis) and B05 (pneumonia versus non-pneumonia lower respiratory infection) for the pneumonia-versus-airway distinction. The pneumonia call at this node averages the B04 and B05 predicted probabilities and compares the average to the mean of their two decision thresholds (0.489 and 0.498, giving 0.494); B04 and B05 share the same pneumonia positive cohort but differ in their negative cohort, so averaging combines the two views of the pneumonia contrast into a single decision. The upper-respiratory branch applies B06 (acute sinusitis versus other URI) and B07 (Group A streptococcal versus other pharyngitis) to refine the anatomical call. Three additional classifiers, B08 (influenza), B09 (COVID-19), and B10 (bacterial versus viral), are applied in parallel as an etiology overlay to any visit routed past the infection gate, since these contrasts cut across anatomical localization. Etiology-correctness was scored per visit as the conjunction of three overlay decisions: the influenza (B08) and COVID-19 (B09) calls each had to match the ICD-derived label exactly (present or absent), and the bacterial-versus-viral (B10) call was required to match only when the visit’s ICD specified a bacterial or viral organism; when the ICD specified neither (the majority of visits), the B10 call was unconstrained. A visit was counted etiology-correct only if all applicable conditions held. The per-binary decision threshold from Section 3.1 is used at each node; thresholds are not jointly optimized within the scope of this pilot.

The leak-safe construction of the cascade evaluation cohort is given in Algorithm 2. Because the cascade applies all 11 models to each visit, every evaluated visit must be unseen by every gate; the cohort therefore excludes, at the patient level, any patient who contributed a visit to the training or development partition of any of the 11 binaries.

##### Algorithm 2 Leak-safe cascade evaluation cohort.

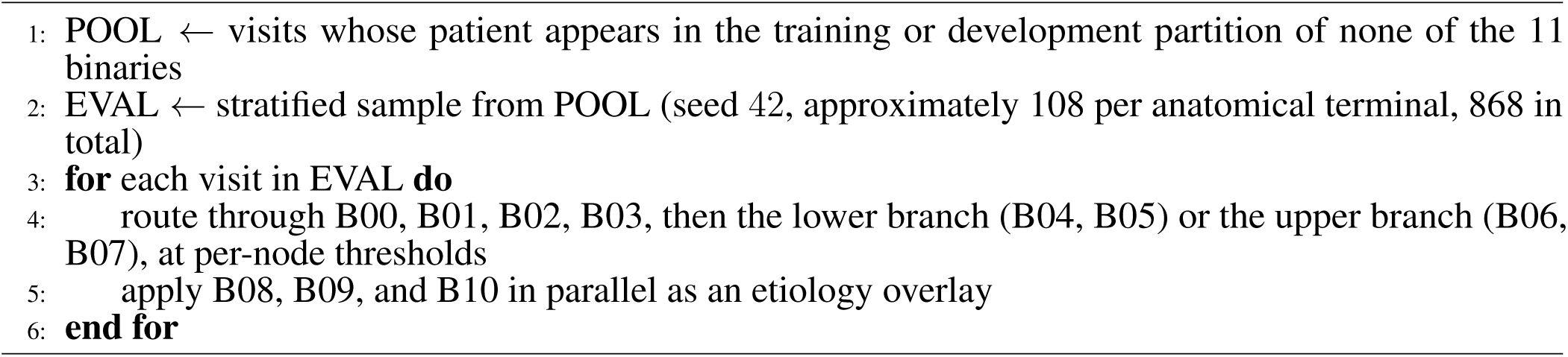

### 2.4 Reproducibility

The modeling pipeline (feature extraction and classification) uses only publicly available components, so the modeling methods can be reproduced from comparable data without any proprietary tool. The one exception is the speaker-diarization front-end (Section 2.3.1), which relies on a commercial cloud speech service; an open diarization pipeline could be substituted at the cost of some front-end fidelity. The acoustic features are produced by openSMILE eGeMAPSv02, librosa, a publicly available valence-arousal-dominance predictor, and praat-parselmouth, and the classifier is a standard feed-forward network. The specification needed to reproduce the work is provided across the paper and the supplementary material. This includes the ICD-10 inclusion and exclusion rules and final cohort counts for all 11 binaries (Section 2.2 and supplementary material), the diarization and audio-preprocessing parameters (Section 2.3.1 and supplementary material), the feature-extraction configuration and stream definitions (Section 2.3.1 and supplementary material), the network architecture (Section 2.3.2 and supplementary material), and the full hyperparameter grid and training settings (supplementary material). Per-class aggregate distributions of all 412 features for each binary are provided as supplementary data. The items that cannot be shared are the raw audio and the visit-level derived feature tables, both of which are protected health information. For the same reason, representative audio waveforms or spectrograms are not shown.

## 3 RESULTS

The 11 binary classifiers trained according to the protocol in Section 2.3 are evaluated on their respective held-out test sets. Section 3.1 reports per-binary discrimination (AUC, AUPRC), calibration (expected calibration error, reliability), and threshold-dependent operating points (sensitivity, specificity, F1) across the panel. Section 3.3 reports the panel of six confounder baselines per binary, with FDR-corrected comparisons against the acoustic model. Detailed per-binary metrics are provided in Table 2 and the headline visualization is shown in Figure 5. Substantive interpretation of failing confounder axes, including stratified within-axis analyses, is deferred to the Discussion in Section 4.

**Figure 5.**
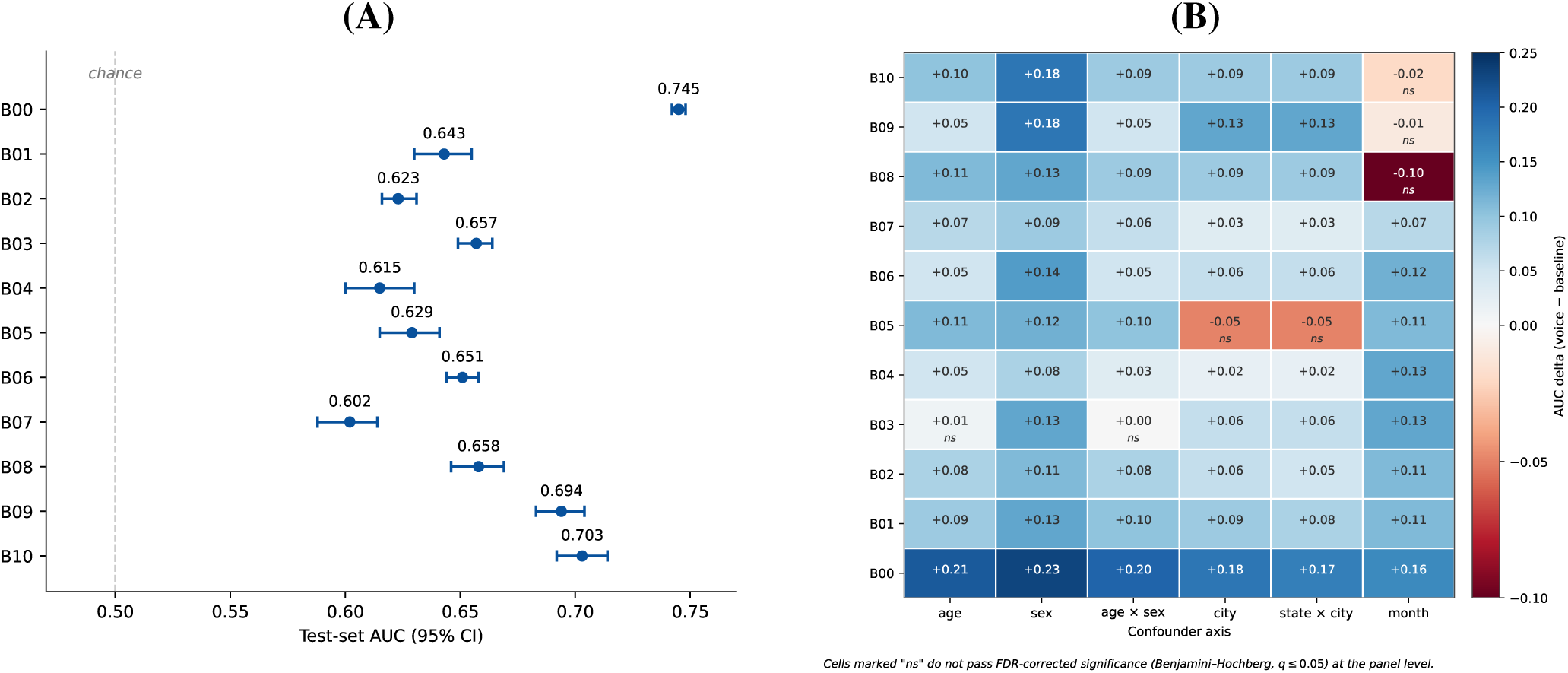
Per-binary discrimination and panel-level confounder analysis on the held-out test set. **(A)** Per-binary AUC across the 11 binary classification tasks, with 95% percentile bootstrap confidence intervals (1,000 resamples). **(B)** Heatmap of the panel of six non-acoustic confounder baselines per binary. Detailed per-binary metrics including AUPRC, F1, sensitivity, specificity, calibration error, and operating-point thresholds are reported in Table 2.

**Table 2.** Per-binary discrimination, calibration, and operating-point metrics on the held-out test set. AUC = area under the ROC curve with 95% percentile bootstrap CI (1,000 resamples). AUPRC = area under the precision-recall curve. F1, sensitivity (Sens.), and specificity (Spec.) reported at the per-binary decision threshold selected by Youden’s J on the development partition. ECE (ef) and ECE (ew) = expected calibration error under equal-frequency and equal-width binning (10 bins each). Brier = Brier score on the test set.

| Binary | AUC [95% CI] | AUPRC | F1 | Sens. | Spec. | ECE (ef) | ECE (ew) | Brier | Threshold |
| --- | --- | --- | --- | --- | --- | --- | --- | --- | --- |
| B00 | 0.745 [0.742, 0.748] | 0.745 | 0.698 | 0.722 | 0.635 | 0.008 | 0.007 | 0.205 | 0.491 |
| B01 | 0.643 [0.630, 0.655] | 0.633 | 0.635 | 0.680 | 0.522 | 0.026 | 0.026 | 0.235 | 0.503 |
| B02 | 0.623 [0.616, 0.631] | 0.618 | 0.583 | 0.567 | 0.606 | 0.014 | 0.009 | 0.238 | 0.516 |
| B03 | 0.657 [0.649, 0.664] | 0.638 | 0.591 | 0.564 | 0.658 | 0.017 | 0.014 | 0.232 | 0.517 |
| B04 | 0.615 [0.600, 0.630] | 0.595 | 0.602 | 0.642 | 0.512 | 0.020 | 0.011 | 0.240 | 0.489 |
| B05 | 0.629 [0.615, 0.641] | 0.604 | 0.583 | 0.585 | 0.585 | 0.022 | 0.019 | 0.236 | 0.498 |
| B06 | 0.651 [0.644, 0.658] | 0.627 | 0.546 | 0.484 | 0.722 | 0.020 | 0.021 | 0.233 | 0.558 |
| B07 | 0.602 [0.588, 0.614] | 0.589 | 0.617 | 0.694 | 0.440 | 0.019 | 0.015 | 0.243 | 0.457 |
| B08 | 0.658 [0.646, 0.669] | 0.625 | 0.634 | 0.679 | 0.557 | 0.011 | 0.006 | 0.231 | 0.478 |
| B09 | 0.694 [0.683, 0.704] | 0.680 | 0.651 | 0.664 | 0.622 | 0.016 | 0.014 | 0.222 | 0.501 |
| B10 | 0.703 [0.692, 0.714] | 0.679 | 0.643 | 0.647 | 0.652 | 0.021 | 0.021 | 0.219 | 0.510 |

### 3.1 Classification and Calibration Performance

Discrimination performance on the held-out test set is summarized in Figure 5A and reported in detail in Table 2. Across the 11 binaries, test-set AUC ranges from 0.602 to 0.745. The gate task (B00, acute respiratory versus acute non-respiratory illness) is the best-performing binary at AUC 0.745 (95% CI 0.742 to 0.748), with the tightest confidence interval reflecting its substantially larger test set. Bacterial versus viral (B10) and COVID-19 versus other acute respiratory infection (B09) form the next tier at AUC 0.703 and 0.694 respectively. A mid-tier of four binaries (B01 acute infection versus chronic, B03 lower versus upper respiratory infection, B06 sinusitis versus other URI, and B08 influenza versus other acute) cluster between 0.643 and 0.658. Three binaries fall in the 0.60 to 0.63 range: B02 (acute infection versus non-infectious mimic), B04 (pneumonia versus acute bronchitis), and B05 (pneumonia versus non-pneumonia LRI). The worst-performing binary is B07 (GAS versus other pharyngitis) at AUC 0.602. AUPRC tracks AUC closely across the panel (0.589 to 0.745, Table 2), and no binary shows a discrepancy between AUC and AUPRC large enough to indicate a precision-recall pathology beyond what discrimination would predict.

Calibration of predicted probabilities is uniformly strong across the panel. Expected calibration error under equal-frequency binning ranges from 0.008 (B00) to 0.026 (B01), and under equal-width binning from 0.006 (B08) to 0.026 (B01), with no binary exceeding 0.026 on either scheme (Table 2). The two schemes track each other within 0.01 across all binaries, indicating that calibration quality is not an artifact of binning choice. No binary shows the systematic over- or under-confidence that would warrant a post-hoc probability re-mapping step before downstream use.

Per-binary decision thresholds selected by Youden’s J on the development partition lie between 0.457 (B07) and 0.558 (B06), close to but not at the prevalence-implied 0.5 (Table 2), reflecting the patient-level balancing applied prior to model training. At these thresholds, sensitivity and specificity show three patterns across the panel. Most binaries operate close to a sensitivity-specificity balance: B05 is exactly balanced (sensitivity 0.585, specificity 0.585), and B09 and B10 are within 0.05 (B09 sensitivity 0.664, specificity 0.622; B10 sensitivity 0.647, specificity 0.652). Two binaries show pronounced asymmetry in opposite directions. B06 (sinusitis versus other URI) trades sensitivity for specificity (sensitivity 0.484, specificity 0.722), reflecting an operating point where the model prefers to confirm sinusitis cautiously. B07 (GAS versus other pharyngitis) trades specificity for sensitivity (sensitivity 0.694, specificity 0.440), reflecting an operating point where the model prefers to flag GAS aggressively. F1 scores at the selected thresholds range from 0.546 (B06) to 0.698 (B00).

### 3.2 Operating-point and prevalence-projected metrics

Discrimination summarized by AUC does not by itself convey triage utility, so we report threshold-dependent predictive metrics at each binary’s operating point and project them across clinically realistic prevalence (Table 3). Because the test set is balanced at the patient level (Section 2.3.2), the directly observed positive and negative predictive values (PPV, NPV) are values at a 50% base rate, while sensitivity, specificity, and the likelihood ratios are computed within each true class and therefore do not depend on prevalence.

**Table 3.** Operating-point predictive metrics on the balanced held-out test set. Sensitivity (Sens.) and specificity (Spec.) are at the per-binary Youden threshold. LR+ and LR− are the positive and negative likelihood ratios. PPV and NPV are the empirical values at the operating point on the balanced held-out test set (test prevalence approximately 50%). The Bayes projection of PPV and NPV across prevalence, computed from sensitivity and specificity, is reported in the supplementary material; its value at a 50% base rate can differ slightly from the empirical values here because the test set is not exactly balanced. Bootstrap 95% confidence intervals (1,000 resamples) for all six metrics are reported in the supplementary material.

| Binary | Sens. | Spec. | LR+ | LR− | PPV | NPV |
| --- | --- | --- | --- | --- | --- | --- |
| B00 | 0.722 | 0.635 | 1.98 | 0.44 | 0.675 | 0.685 |
| B01 | 0.680 | 0.522 | 1.42 | 0.61 | 0.595 | 0.612 |
| B02 | 0.567 | 0.606 | 1.44 | 0.71 | 0.600 | 0.573 |
| B03 | 0.564 | 0.658 | 1.65 | 0.66 | 0.619 | 0.605 |
| B04 | 0.642 | 0.512 | 1.32 | 0.70 | 0.567 | 0.591 |
| B05 | 0.585 | 0.585 | 1.41 | 0.71 | 0.581 | 0.589 |
| B06 | 0.484 | 0.722 | 1.74 | 0.71 | 0.626 | 0.593 |
| B07 | 0.694 | 0.440 | 1.24 | 0.70 | 0.555 | 0.588 |
| B08 | 0.679 | 0.557 | 1.53 | 0.58 | 0.595 | 0.644 |
| B09 | 0.664 | 0.622 | 1.76 | 0.54 | 0.639 | 0.648 |
| B10 | 0.647 | 0.652 | 1.86 | 0.54 | 0.640 | 0.658 |

Across the panel the likelihood ratios are modest. The positive likelihood ratio ranges from 1.24 (B07) to 1.98 (B00) and the negative likelihood ratio from 0.44 (B00) to 0.71. No binary reaches the conventional thresholds for a clinically decisive test (positive likelihood ratio above 5 or negative likelihood ratio below 0.2). At the balanced 50% operating point, PPV ranges from 0.555 (B07) to 0.675 (B00) and NPV from 0.573 (B02) to 0.685 (B00).

The predictive values change substantially when projected to the natural prevalence of each contrast. For the low-prevalence pathogen contrasts, influenza (B08, natural prevalence 4.7%) projects to PPV 0.07 and NPV 0.97, and COVID-19 (B09, natural prevalence 5.8%) projects to PPV 0.10 and NPV 0.97. At these prevalences a positive acoustic call would require confirmatory testing, and the only plausible operating use is rule-out rather than rule-in. The full prevalence projection for all 11 binaries is reported in the supplementary material. Calibration is reported in Table 2 (expected calibration error and Brier score). Decision-curve analysis for all 11 binaries, at the balanced 50% test prevalence, is reported in the supplementary material.

### 3.3 Confounder analysis

The panel of six non-acoustic baselines per binary is summarized in Figure 5B. Baseline AUCs range from 0.489 (B04 month) to 0.753 (B08 month), and acoustic-model deltas relative to each baseline range from −0.095 (B08 month) to +0.232 (B00 sex). Of the 66 (binary, axis) comparisons in the panel, 59 pass the FDR-corrected paired bootstrap criterion at *q* ≤ 0.05 and 7 do not.

Six binaries pass on all six axes: B00, B01, B02, B04, B06, and B07. For these binaries, the acoustic model’s discrimination exceeds every baseline by a margin that survives panel-wide multiple-testing correction.

Five binaries fail on at least one axis. B03 (lower versus upper respiratory infection) fails on age (baseline AUC 0.650, delta +0.007) and on the age-by-sex joint (baseline AUC 0.653, delta +0.004). B05 (pneumonia versus non-pneumonia LRI) fails on patient city (baseline AUC 0.678, delta −0.049) and on the state-by-city joint (baseline AUC 0.679, delta −0.050); these two are the only cells in the panel where the acoustic model is outperformed by a single-axis prevalence lookup. B08 (influenza versus other acute respiratory infection), B09 (COVID-19 versus other acute respiratory infection), and B10 (bacterial versus viral) each fail on calendar month, with baseline AUCs of 0.753, 0.700, and 0.719 respectively.

Substantive interpretation of these five failure cases, including stratified within-axis analyses that quantify the residual acoustic signal after the confound is conditioned on, is taken up in the Discussion (Section 4.2).

### 3.4 Feature-family ablation

To identify which feature families carry the discrimination, each binary’s model was retrained on feature-stream subsets: the full set of nine streams, each single stream alone, and the full set with one family removed (leave-one-family-out). These runs use a single cross-validation fold, so the full-set AUC here (0.604 to 0.747) is the single-fold counterpart of the cross-validated AUC in Table 2, and differences below approximately 0.01 are within run-to-run variation. Full per-family results are in the supplementary material; Table 4 summarizes the two patterns that hold across the panel.

**Table 4.** Feature-family ablation summary (single fold). Full AUC is the nine-stream model. “librosa LOO Δ” is the AUC drop when the librosa stream is removed; “librosa-only AUC” is the librosa stream evaluated alone; “speech-rate/pause LOO Δ” is the AUC drop when the praat-parselmouth speech-rate-and-pause family is removed. Full per-family leave-one-out and single-family results are in the supplementary material.

| Binary | Full AUC | librosa LOO $\Delta$ | librosa-only AUC | speech-rate/pause LOO $\Delta$ |
| --- | --- | --- | --- | --- |
| B00 | 0.747 | 0.025 | 0.719 | 0.001 |
| B01 | 0.638 | 0.014 | 0.631 | −0.004 |
| B02 | 0.626 | 0.009 | 0.612 | 0.005 |
| B03 | 0.656 | 0.017 | 0.641 | 0.001 |
| B04 | 0.604 | 0.023 | 0.597 | 0.003 |
| B05 | 0.633 | 0.029 | 0.632 | −0.003 |
| B06 | 0.649 | 0.011 | 0.638 | −0.001 |
| B07 | 0.604 | 0.031 | 0.592 | 0.001 |
| B08 | 0.657 | 0.017 | 0.650 | −0.004 |
| B09 | 0.687 | 0.017 | 0.679 | 0.000 |
| B10 | 0.703 | 0.046 | 0.689 | 0.001 |

First, the librosa stream (Mel-frequency cepstral, spectral, and rhythm features) carries most of the discrimination on every binary. Removing it produces the largest leave-one-family-out drop for all 11 binaries (0.009 to 0.046), and the librosa stream alone recovers nearly the full AUC (within 0.001 to 0.028). Every other family, including all six openSMILE groups and the affective valence-arousal-dominance stream, has a leave-one-family-out delta below approximately 0.01 on most binaries, several of them slightly negative.

Second, the models do not behave as a recording-length or speech-density proxy. The speech-rate-and-pause family (total syllables, pauses, phonation time, articulation rate) has a leave-one-family-out delta between −0.004 and +0.005 across all 11 binaries and is among the weakest families in isolation (0.526 to 0.600). Speech density therefore contributes negligibly to the trained models, which argues against the concern that performance reflects a recording-length artifact rather than acoustic content. Because these deltas fall within the single-fold variation noted above, the ablation bounds the contribution of these features as small rather than proving it zero; the stronger guarantee that performance is not a recording-length proxy is structural, since recording duration and segment count are not included as features.

### 3.5 Location-stratified generalization

The dataset does not carry clinic or site identifiers, so generalization across settings was assessed using the patient city and state of residence, the only location fields available. Two complementary analyses were run. In the first, entire cities were assigned to five disjoint folds so that no city appeared in both training and test. In a five-fold city-grouped cross-validation, a model was retrained on four folds and evaluated on the held-out cities, giving one held-out-city AUC per fold; Table 5 reports the mean and standard deviation across the five folds. Across the panel the held-out-city AUC was close to the random-split AUC: the gap between the champion AUC (the random-split test AUC reported in Table 2) and the mean held-out-city AUC was at most 0.020 on every binary (0.020 for B09, 0.018 for B08, 0.017 for B03) and was slightly negative for B00. Discrimination on entirely unseen cities therefore matches the random-split estimate, indicating that the models do not depend on city-specific patterns.

**Table 5.** City-grouped held-out generalization. Entire cities were assigned to five disjoint folds (no city in both training and test); the model was retrained on four folds and evaluated on the held-out cities. “Held-out-city AUC” is the mean and standard deviation across folds; “Gap” is the champion AUC (the random-split AUC of Table 2) minus the mean held-out-city AUC (positive means lower performance on unseen cities).

| Binary | Champion AUC | Held-out-city AUC (mean ± SD) | Gap |
| --- | --- | --- | --- |
| B00 | 0.745 | 0.746 ± 0.006 | −0.001 |
| B01 | 0.643 | 0.639 ± 0.006 | 0.004 |
| B02 | 0.623 | 0.616 ± 0.013 | 0.007 |
| B03 | 0.657 | 0.640 ± 0.008 | 0.017 |
| B04 | 0.615 | 0.602 ± 0.009 | 0.013 |
| B05 | 0.629 | 0.620 ± 0.016 | 0.009 |
| B06 | 0.651 | 0.646 ± 0.007 | 0.005 |
| B07 | 0.602 | 0.595 ± 0.004 | 0.007 |
| B08 | 0.658 | 0.640 ± 0.011 | 0.018 |
| B09 | 0.694 | 0.674 ± 0.019 | 0.020 |
| B10 | 0.703 | 0.694 ± 0.006 | 0.009 |

In the second analysis, the model was trained on California visits and tested on the rest of the country, and the reverse. This coarse regional swap produced larger drops, concentrated in the binaries already flagged in the confounder analysis. B00 remained stable (0.723 and 0.722 against a champion AUC of 0.745), while B05 fell to 0.562 and 0.525, close to chance. The seasonal etiology binaries also dropped (B08 to 0.610 and 0.617, B09 to 0.640 and 0.669, B10 to 0.644 and 0.664). Per-city stratified AUC, reported in the supplementary material, showed the same pattern: the visit-weighted within-city AUC was close to the overall AUC for most binaries but lower for B05 (0.569 versus 0.629). The California-versus-rest results and the per-site stratified AUC are reported in full in the supplementary material.

## 4 DISCUSSION

### 4.1 Summary of findings

Across the panel of 11 binary classification tasks, voice models trained on conversational primary care audio achieved above-chance discrimination on every contrast tested. Test-set AUC ranged from 0.602 (B07, Group A streptococcal versus other pharyngitis) to 0.745 (B00, acute respiratory versus acute non-respiratory illness), with calibration uniformly strong and confidence intervals of width 0.006 to 0.030. The AUC range in absolute terms is moderate (Çorbacıoğlu and Aksel, 2023). Two aspects of the experimental setting bear on how this should be read. First, the speech condition is conversational ambient audio from the natural primary care encounter, with no scripted task, no prompted vocalization, and no curated recording environment. Prior voice-based respiratory work has predominantly used scripted speech (sustained vowel phonation, prompted counting, read sentences) or solicited cough produced on demand in controlled or semi-controlled settings, both of which constrain the within-patient acoustic variability that a model must overcome to be useful in practice. Second, the negative cohort for every binary in this panel is itself a sick patient population presenting to the same clinical encounter with a different diagnosis, not a healthy control. The classifier is therefore asked to distinguish between two illness states rather than between illness and health. Both conditions are stricter than the conditions of most prior reported results, and an above-chance signal under these conditions provides an empirically meaningful baseline even where the absolute AUC is moderate.

Within the panel, three patterns are visible. The gate task (B00) achieves the strongest discrimination, consistent with the largest test set and with the broadest acoustic contrast (any respiratory illness against any acute non-respiratory illness). The pathogen-specific and etiology contrasts (B09 COVID-19, B10 bacterial versus viral) reach AUC 0.69 to 0.70. For reference, prior crowdsourced COVID cough studies report AUC 0.70 to 0.85 against healthy controls (Xia et al., 2021; Muguli et al., 2021). Our results sit at the lower end of that range under stricter conditions: conversational speech rather than cough sounds, and sick-versus-sick rather than sick-versus-healthy contrasts. The within-respiratory anatomical and infection-versus-mimic contrasts (B01, B02, B03, B04, B05, B06, B07, B08) cluster in the 0.60 to 0.66 range. The interpretive question for the remainder of this discussion is whether each of these signals reflects genuine acoustic discrimination of the clinical contrast or a demographic, geographic, or temporal artifact, and we take this up at the panel level in Section 4.2.

### 4.2 Interpreting the confounder analysis

The panel-level confounder analysis reported in Section 3.3 found that 5 of the 11 binaries failed at least one axis at FDR *q* ≤ 0.05. The five failures fall into three distinct categories, and their interpretation requires more than the panel-level pass-or-fail verdict. Table 6 summarizes the acoustic-model AUC, the failing confounder-model AUC, and the within-stratum median acoustic-model AUC for each failing binary.

**Table 6.** Per-binary AUC of the acoustic model, the failing confounder-baseline model, and the within-stratum median acoustic-model AUC, for the five binaries that fail at least one confounder axis. “Acoustic-model AUC” is the panel-headline AUC for the binary; “Confounder-model AUC” is the AUC of the non-acoustic prevalence lookup along the failing axis; “Within-stratum median AUC” is the median across strata of the failing axis (5-year age bins for B03, calendar months for B08, B09, B10, and patient city for B05) of the acoustic model’s AUC computed within each stratum. For B05, no within-stratum median is reported (n/a) because the failing axis reflects a city-level documentation artifact rather than an acoustic confound, so within-city conditioning would remove the variance the model relies on and is not an informative rescue (Section 4).

| Binary | Failing axis | Acoustic-model AUC | Confounder-model AUC | Within-stratum median AUC |
| --- | --- | --- | --- | --- |
| B03 | age | 0.657 | 0.650 | 0.615 |
| B05 | city, state × city | 0.629 | 0.678 | n/a |
| B08 | date (month) | 0.658 | 0.753 | 0.627 |
| B09 | date (month) | 0.694 | 0.700 | 0.698 |
| B10 | date (month) | 0.703 | 0.719 | 0.683 |

The first category is structural demographic leakage. B03 (lower versus upper respiratory infection) fails on age and on age-by-sex because the underlying clinical contrast is itself age-stratified in primary care: pediatric exclusion notwithstanding, upper respiratory infections are diagnostically more common among younger adults while lower respiratory infections become relatively more common with age, and an age-only prevalence lookup picks up much of this signal. When the analysis is restricted to within-age-bin discrimination, the voice model still achieves median AUC 0.615 across 5-year age bins from 18 to 82 years, with within-bin AUCs largely concentrated in the 0.59 to 0.63 range. A genuine acoustic signal therefore persists once the demographic prior is removed, and the headline AUC of 0.657 should be read as a combination of that genuine within-stratum signal and the age prior the cohort carries. The age prior is also clinically informative and is not a confound that any deployed system would have reason to ignore.

The second category is seasonal and pandemic-wave temporal structure for the three pathogen-specific contrasts. B08 (influenza) fails on calendar month with a month-baseline AUC of 0.753 against the voice headline of 0.658, the largest baseline-versus-voice gap in the panel. B09 (COVID-19) and B10 (bacterial versus viral) also fail on month, with month-baseline AUCs of 0.700 and 0.719 respectively. The interpretation here is straightforward and clinically expected: influenza is overwhelmingly concentrated in the winter months, COVID-19 follows pandemic waves that are themselves temporal, and the bacterial-viral mix shifts seasonally because strep season and viral cold season do not coincide. Knowing the month is therefore strongly predictive of the diagnosis. Within-month analyses are informative: the median within-month voice-model AUC is 0.627 for B08 (range 0.545 to 0.672 across calendar months), 0.698 for B09 (range 0.669 to 0.723), and 0.683 for B10 (range 0.623 to 0.710). For B09 the within-month voice AUC is essentially unchanged from the headline AUC (0.698 versus 0.694), and for B10 it is modestly lower (0.683 versus 0.703). Because the calendar month carries no information within a single month, this indicates that the B09 and B10 voice discrimination persists when month is held fixed and is therefore largely independent of the seasonal prior rather than a proxy for it. For B08, the within-month AUC is meaningfully lower than the headline (0.627 versus 0.658), suggesting that the headline performance for influenza is more dependent on the seasonal prior than for B09 or B10. In all three cases some discriminatory power persists in the voice signal when month is held fixed, but the temporal prior remains a clinically important and operationally available piece of information that a deployed system would likely condition on rather than exclude.

The third category is a cohort-level confound. B05 (pneumonia versus non-pneumonia LRI) fails on city and on state-by-city, with city-baseline AUC of 0.678 exceeding the voice-model AUC of 0.629. This is the only cell in the panel where a non-acoustic single-axis baseline outperforms the voice model. The most plausible explanation is the J22 unspecified-LRI label noise in the B05 negative cohort, which we flagged at cohort construction in Section 2.2. The J22 code is assigned in practice when the clinician documents a lower respiratory infection without committing to either pneumonia or bronchitis, and the choice of J22 versus a specific code is itself a clinician- and clinic-level habit, not a clinically meaningful disease distinction. A patient city baseline picks this up because clinics in a given city share documentation habits. Within-city analysis is therefore not informative as a rescue (the conditioning would remove the very variance the model would need to learn from), and we treat B05 as the one binary in the panel where the apparent acoustic discrimination is partly explained by a clinic-documentation artifact rather than by a clinical acoustic signal. The pneumonia signal itself is more cleanly assessed in B04 (pneumonia versus acute bronchitis, the same positive cohort with a cleaner negative cohort), where the AUC of 0.615 is lower but is not subject to the same confound.

The location-stratified experiments in Section 3.5 corroborate this reading. Fine-grained generalization holds: when whole cities are held out of training, discrimination stays within 0.020 of the random-split estimate on every binary. A coarse California-versus-rest swap, by contrast, degrades exactly the binaries flagged here, with B05 falling close to chance and the three seasonal etiology binaries (B08, B09, B10) dropping by 0.03 to 0.10. The city and month confounds are therefore visible not only in the prevalence-lookup baselines but also in how the acoustic models transfer across regions. We do not have clinic or site identifiers and so cannot stratify below the city level.

We deliberately did not include demographic, geographic, or temporal variables as conditioning features in the acoustic models, since the goal of this pilot is to isolate the voice-only signal at each contrast; an ablation that fuses voice with these priors and quantifies the marginal contribution of each is left to future work.

### 4.3 The cascade composition: aggregate evaluation and worked examples

The four case studies shown in Figures 6 and 7 were drawn from a broader cascade evaluation that aggregated the 11 trained classifiers into a single hierarchical screening pipeline and applied them to 868 test-set patients, drawn under a strict leak-safe sampling rule that excluded, at the patient level, any patient appearing in the training or development partition of any of the 11 binaries. The cascade is reproducible from the 11 trained classifiers, their splits, and the per-node decision thresholds. Across this evaluation, 20.6% of patients reached the correct anatomical terminal in the cascade, 42.1% had the correct etiology overlay call, and 8.6% were fully correct on both anatomical and etiology assessment. A random-gate Monte Carlo baseline, in which each cascade decision is replaced by a coin flip at the per-step positive rate, yields 13.1% anatomical, 23.5% etiology, and 3.3% fully correct, indicating that the cascade composition performs above chance on every dimension by margins of approximately 1.6-fold (anatomical), 1.8-fold (etiology), and 2.6-fold (fully correct). The etiology rate is not directly comparable to the anatomical rate: the bacterial-versus-viral (B10) call is scored only when the visit’s ICD code specifies a bacterial or viral organism (Section 2.3.4), and is unconstrained on the majority of visits that specify neither. The cascade is reported as an exploratory demonstration that the independently trained classifiers can be composed end to end, not as a triage system ready for clinical use.

**Figure 6.**
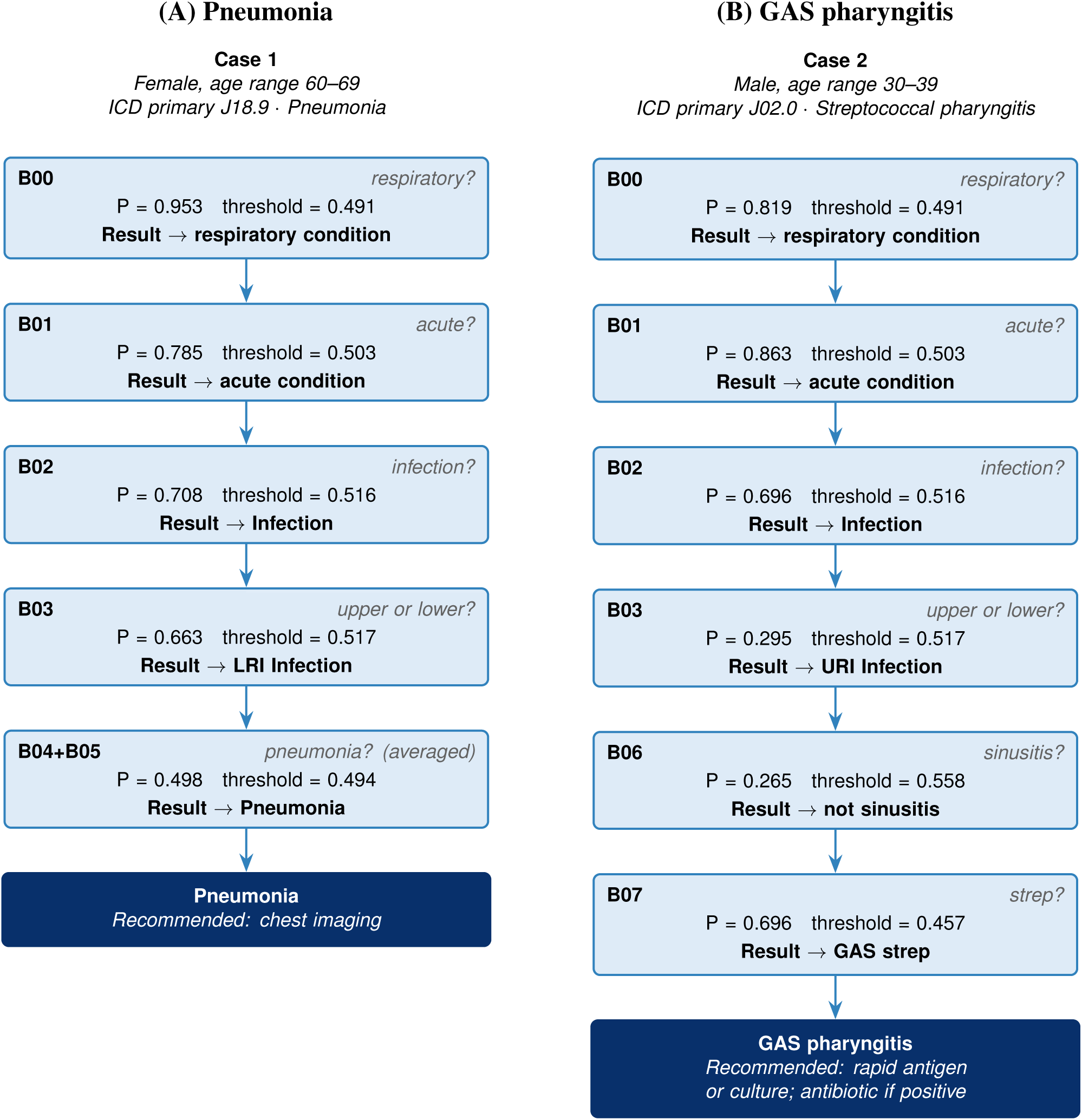
Worked examples of cascade traversal on test-set patients (part 1 of 2). **(A)** A pneumonia visit is routed through the anatomical gates to a pneumonia terminal. **(B)** A streptococcal pharyngitis visit is routed through the upper respiratory branch to a Group A streptococcal (GAS) terminal.

**Figure 7.**
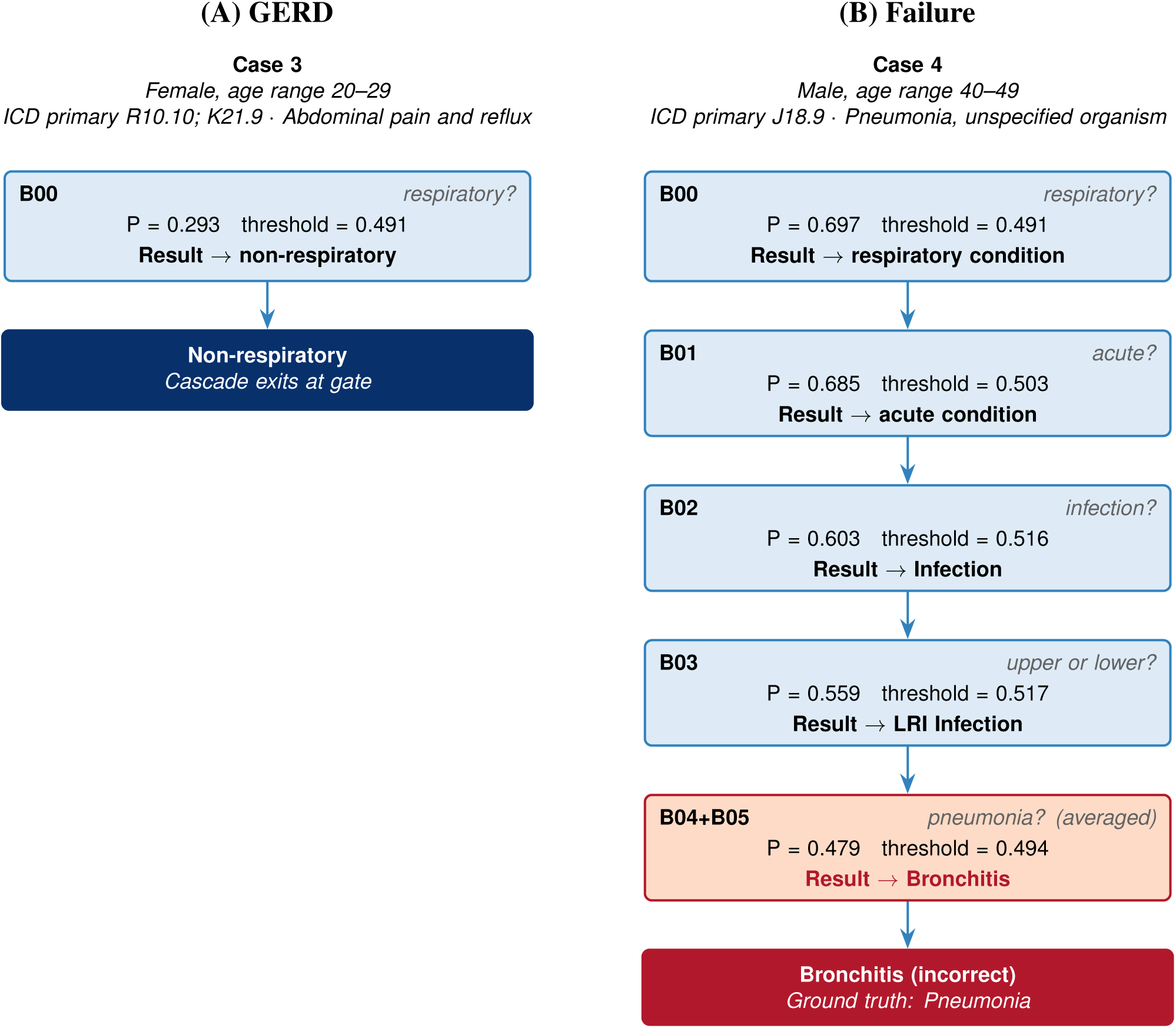
Worked examples of cascade traversal on test-set patients (part 2 of 2). **(A)** A presentation of abdominal pain and reflux (R10.10, K21.9) is correctly rejected at the gate task (B00) as non-respiratory, exiting the cascade. **(B)** A confirmed pneumonia case (J18.9) is correctly routed through the first four gates but misclassified at the final B04+B05 step, where the pneumonia probability falls just below the decision threshold, and the cascade exits at acute bronchitis; this illustrates the canonical compositional failure mode of a stacked cascade.

Two patterns in these numbers warrant explicit comment. First, the anatomical-correct rate is meaningfully lower than the etiology-correct rate. The reason is structural rather than acoustic: the anatomical cascade (B00, B01, B02, B03, B04, B05) requires four to five sequential gate decisions to be jointly correct, while the etiology overlay (B08, B09, B10) is a single-shot parallel call that does not depend on upstream routing. Per-step gate accuracies on the cascade evaluation cohort range from approximately 56% to 64%, and the multiplicative composition of four to five such gates is consistent with the observed 20.6% anatomical rate. This is the standard error-propagation behavior expected when independently trained classifiers are stacked into a deep decision sequence without joint training, and it is the principal methodological reason a hierarchical cascade composed from independently optimized binaries underperforms the per-binary AUCs reported in Section 3.1. The 8.6% fully-correct rate, which requires both the anatomical traversal and the etiology overlay to be correct simultaneously, is therefore best read as a conservative estimate that reflects compositional error propagation across independently trained gates, which joint training may improve, and the gap between independent-task performance and composed cascade performance is itself the most informative methodological finding of this analysis.

Second, the four illustrative case studies span three successful traversals and one informative failure. Figure 6A routes a pneumonia visit (J18.9) through the full lower respiratory branch (B00 respiratory → B01 acute → B02 infection → B03 lower → B04+B05 pneumonia) to a pneumonia terminal. Figure 6B routes a Group A streptococcal pharyngitis visit (J02.0) through the upper respiratory branch (B00 → B01 → B02 → B03 upper → B06 not sinusitis → B07 GAS) to a GAS terminal. Figure 7A shows the cascade correctly rejecting a presentation of abdominal pain and reflux (R10.10, K21.9) at the gate task without proceeding further, illustrating the importance of B00 as a screening gate against non-respiratory complaints that can present with cough or throat symptoms. Figure 7B illustrates a compositional failure-by-narrow-margin: a 40–49-year-old patient with confirmed pneumonia (J18.9) is routed correctly through the first four gates (respiratory, acute, infection, lower respiratory) but the final B04+B05 step returns a pneumonia probability just below the decision threshold, and the cascade exits at bronchitis. The auto-generated recommendation for bronchitis (supportive care, no antibiotic) is the wrong clinical recommendation for a patient with confirmed pneumonia, and this case illustrates the canonical compositional failure mode of a stacked cascade: the upstream gates all behave correctly but a borderline probability at the deepest gate flips the terminal call.

The cascade results, both the aggregate and the worked examples, motivate a clear direction for future work that we return to in Section 4.5: joint training of the panel as a single hierarchical model rather than as 11 independently trained binaries. The per-binary models reported here were optimized in isolation; the cascade applies them as a sequence; no part of the training procedure was aware that compositional error propagation would compound per-step accuracies when the gates are composed. Joint training, in which the loss function reflects the cascade traversal rather than 11 independent binary losses, would allow the upstream gates to learn representations that are useful for the downstream gates and would, in principle, narrow the gap between the per-binary AUCs of Section 3 and the composed cascade performance reported here.

### 4.4 Limitations

Several limitations of the present work constrain the strength of the claims that can be drawn from the reported results.

Clinical labels in this study are ICD-10 codes assigned at the visit by the treating clinician, not laboratory-confirmed or radiographically confirmed diagnoses. ICD coding fidelity varies by code and by setting. For some binaries the coding is closely aligned with the underlying clinical entity (J18 pneumonia is typically coded after imaging confirmation; J02.0 streptococcal pharyngitis after rapid antigen or culture confirmation); for others the coding reflects clinical impression alone (J01 acute sinusitis is in practice often assigned to viral upper respiratory presentations with sinus-symptom predominance, as discussed in Section 2.2). The acoustic labels therefore inherit the noise structure of the coding system, and a portion of the observed AUC ceiling on every binary is attributable to label noise rather than to a limit on what voice can in principle distinguish. We do not have access to the underlying laboratory, antigen, or imaging results for these visits, so we cannot report the proportion of cases in any binary that is supported by confirmatory testing. The level of confirmation behind a code is expected to differ across conditions, for example imaging for pneumonia, rapid antigen for influenza and streptococcal pharyngitis, and clinical impression for acute sinusitis.

External validation is absent. The dataset is drawn entirely from a single US primary care network, and the train, development, and test partitions are sampled from the same source population. Geographic and demographic distributions reported in Section 2.1 are broad relative to prior conversational voice datasets in this space, but they do not constitute external validation in the sense that the deployment-relevant clinical informatics community would require before any operational claim. The present work establishes that an acoustic signal exists at the contrasts and conditions tested; demonstrating that the signal generalizes to other primary care networks, other recording pipelines, other patient populations, and other coding practices is the necessary next step before any deployment-adjacent claim. We frame this work explicitly as a baseline and proof-of-concept that motivates such future validation, not as a substitute for it. This concern is not hypothetical: cough-acoustic models for COVID-19 and tuberculosis have shown marked performance drops when validated on a population different from their training population (Zimmer et al., 2026b,a), and the California-versus-rest degradation reported in Section 3.5 shows the same pattern within this dataset.

The B01 (acute infection versus chronic respiratory disease) negative cohort is substantially enriched for patients on inhaled corticosteroid therapy, which is known to produce dysphonia and voice quality changes through laryngeal deposition. Inhaled corticosteroid exposure cannot be reliably identified from available metadata, and we therefore cannot disentangle the contribution of medication exposure from the contribution of underlying disease state to the observed B01 discrimination. The B01 result should be read with this confound noted, and we have not attempted to correct for it in the present analysis.

The B05 (pneumonia versus non-pneumonia LRI) negative cohort includes a substantial proportion of J22 unspecified-acute-LRI codes, which as discussed in Section 4.2 reflects clinic-level documentation habits as much as a clinical disease entity. The patient-city confound surfaced in the confounder analysis is the empirical signature of this label-noise structure, and we do not claim a clean acoustic discrimination of pneumonia from bronchitis at B05; the cleaner pneumonia signal in this work is at B04, against a more homogeneous bronchitis negative cohort.

The diarization and patient-channel extraction pipeline that produces the audio input to feature extraction is operated upstream of the modeling work reported here. The Chirp 3 speech recognition model, the gemini-flash-3.1-lite large language model used to identify the patient speaker tag, and the audio recomposition step are described in Section 2.3.1 at the level of their operational specification, but we did not independently validate the speaker identification accuracy on a manually labeled reference set. Errors in patient-channel extraction would propagate into every downstream feature and every reported result; the reliability of the pipeline in practice is suggested by the panel-level AUCs but is not formally established by the present study. A manually annotated validation subset was outside the scope of this secondary analysis: the data-use agreement restricts use of the recordings to the aims of the approved protocol and does not permit sharing the audio with annotators or granting listening access for manual labeling. The pipeline therefore runs without manual speaker labels, as any deployed system would. We also did not locate a published diarization error rate for the speech recognition model used. Three features of the design limit the influence of extraction errors on the reported contrasts. The same diarization pipeline is applied to every visit identically, so it does not introduce systematic differences between the two arms of a binary. The models use only acoustic and affective features computed from the audio and do not use the transcript content, word identity, or text embeddings, so they cannot exploit what was said. Every binary contrasts one symptomatic group against another, so structural differences between healthy and ill speech do not apply, although we cannot rule out that two respiratory conditions differ in conversational timing or turn structure as well as in voice. The temporal and speech-rate and pause features, which are the descriptors most susceptible to a diarization artifact, contribute negligibly in the feature-family ablation (Section 3.4), which further limits the plausible effect of extraction errors.

Several further sources of variation are not modeled because the necessary metadata are not available. The recordings come from the routine ambient-documentation system, and we do not have the recording device, microphone type, or microphone distance for each visit. We also do not have language or accent, race or ethnicity, or symptom severity, and comorbidity and medication exposure are observable only to the extent that they are coded, which undercounts them. We cannot model these factors and note them as residual confounders; the city-level and seasonal confounds quantified in Section 3.3 are the subset we were able to test with the available metadata.

### 4.5 Positioning and future work

The present work sits adjacent to a body of prior literature, surveyed in detail in Section 2.1.2, that has demonstrated acoustic signal for specific respiratory diseases using scripted speech or solicited cough collected primarily in crowdsourced or controlled clinical settings. Against this prior literature the contribution here is differentiated along three axes that we set out in the Introduction. First, the speech condition is conversational rather than scripted, and the audio is acquired passively from the natural primary care encounter rather than from a structured elicitation task. Second, the contrasts tested are differential decisions among sick patients, not single-condition detections against healthy controls. Third, the evaluation operates at a scale of unique adult patients that is approximately an order of magnitude above the largest prior conversational respiratory dataset, and that approaches the operational scale at which a primary care population would actually need to be served. We do not claim novelty in the modeling architecture or in the feature extraction pipeline, which use established acoustic feature sets and standard supervised learning. The novelty is in the empirical demonstration that voice carries discriminating signal across a panel of clinically meaningful contrasts under these stricter conditions.

Several directions for future work follow from the present results. Joint training of the panel as a single hierarchical model, motivated by the compositional error propagation observed in the cascade analysis in Section 4.3, is the most direct methodological extension. A model whose loss function reflects the cascade traversal would in principle allow upstream gates to learn representations that are useful for downstream gates, narrowing the gap between per-binary AUCs and composed cascade performance. Complementary to joint training, node-level error analysis, that is, quantifying where in the cascade traversal errors are introduced, and probabilistic error propagation, which would carry per-gate uncertainty forward rather than committing a hard decision at each node, are natural extensions for understanding and mitigating the compositional error observed here. External validation, on data from primary care networks not represented in this study, is the most consequential empirical extension and is a prerequisite to any operational or deployment-adjacent claim. Prospective evaluation, in which the modeling pipeline is run against incoming visits in real time rather than against a retrospective cohort, would address sources of temporal drift and population shift that retrospective evaluation cannot. Strengthening label quality, either by sampling a subset of visits for chart review or for laboratory and imaging confirmation of the ICD code, would address the coding-fidelity limitations described in Section 4.4 and would establish performance ceilings that are not currently distinguishable from coding noise. Feature-level explainability and analysis, identifying which acoustic features drive each binary’s discrimination and how they map to specific speech-production mechanisms, are out of scope of the present study and represent another direction for future work. Finally, the present panel covers 11 binary contrasts chosen as a proof-of-concept set; extending the panel to additional clinically relevant contrasts (chronic respiratory disease subtypes, severity stratification within pneumonia, comorbidity interactions) is a natural direction once joint training and external validation are established.

## 5 CONCLUSION

This paper has presented a pilot study of voice-based respiratory triage from conversational primary care audio. Eleven binary classification tasks, organized around a clinical respiratory decision cascade and selected by criteria of clinical meaningfulness, data sufficiency, and biological plausibility, were trained and evaluated independently on a dataset of 514,377 visits from 379,225 unique adult patients drawn from a US primary care clinic network. Each binary achieved above-chance discrimination on its held-out test set, with AUC ranging from 0.602 to 0.745 and uniformly strong calibration. A panel-level confounder analysis along six demographic, geographic, and temporal axes found that six of the 11 binaries pass on all axes after multiple-testing correction; the remaining five failures were either explained by within-stratum analyses that show residual acoustic discrimination once the confounder is held fixed (weakest for the influenza binary, where the seasonal prior dominates) or, in one case, reflect a documentation artifact in the cohort construction rather than an acoustic confound. A hierarchical screening cascade composed from the 11 trained classifiers operated above chance on a strict leak-safe test cohort but at a substantial gap below per-binary AUCs, consistent with compositional error propagation in independently trained classifiers stacked into a decision sequence and motivating joint training of the panel as a single hierarchical model.

We frame this work as a baseline and proof-of-concept. The conditions of the evaluation, including conversational ambient audio at population scale, differential-diagnosis contrasts against sick rather than healthy negatives, and ICD-coded clinical labels under real-world coding noise, are substantively stricter than those of prior voice and cough work in respiratory disease. The moderate AUC range observed under these conditions establishes that voice carries discriminating signal at clinically meaningful contrasts in primary care, and the cascade composition is an exploratory demonstration that the trained classifiers can be assembled into a decision sequence even without joint optimization. Substantial future work, including external validation, prospective evaluation, label-quality strengthening, and joint training of the cascade, is required before any operational claim can be supported. We hope the present results, the cohort design, and the modeling and evaluation protocol provide a useful foundation for that work.

## Supporting information

Supplementary data CSV file

Supplementary data description

## ETHICS STATEMENT

This study was reviewed by Pearl IRB (Indianapolis, IN, USA) and determined to be exempt human-subjects research under 45 CFR 46.104(d)(4)(iii) (secondary research use of identifiable private information) on 15 July 2025 (IRB ID 2025-0405). As exempt research, it was not subject to the Common Rule’s informed-consent requirements, and study-specific consent was not obtained for this secondary analysis. Because voice recordings are a biometric identifier and therefore protected health information (PHI), the IRB separately granted a waiver of HIPAA authorization under 45 CFR 164.512(i), finding that the use of PHI involved no more than minimal risk to privacy and that the research could not practicably be conducted without access to and use of the unaltered recordings. Secondary research use was disclosed to patients through the originating clinic network’s Notice of Privacy Practices, under which patients could opt out of research participation; patients who opted out were excluded. The recordings and associated clinical data were transferred to the authors as protected health information under a data-use agreement that prohibited re-identification and restricted use to the aims of this protocol. All audio and derived data were stored and processed only within a controlled cloud environment, with no local downloads and no storage on personal devices.

## AUTHOR CONTRIBUTIONS

VR led the design, implementation, and execution of the study, performed all data analysis, led the data preparation for the experiments reported here, and wrote the original draft of the manuscript. CN contributed to the study design, carried out preliminary data preparation, supervised the research and provided project administration, and contributed to and edited the manuscript. All authors contributed to the article and approved the submitted version.

## ACKNOWLEDGMENTS

The authors thank Amit Mehta, MD, who served as Principal Investigator for the IRB-approved protocol, for medical insights, and Oleksii Abramenko, Abinay Reddy Naini, PhD, and Peh Teh for manuscript feedback and review.

## DATA AVAILABILITY STATEMENT

The raw recordings underlying this study cannot be shared owing to patient-privacy obligations. The derived and transformed datasets generated for this study are proprietary to Amplifier Health, Inc. and are not publicly available. Aggregate and distributional summaries supporting the findings are provided in the Supplementary Material; further enquiries may be directed to the corresponding author.

## REFERENCES

Behrman, A. (2023). Speech and Voice Science (San Diego, CA: Plural Publishing), 4th edn.

Benjamini, Y. and Hochberg, Y. (1995). Controlling the false discovery rate: A practical and powerful approach to multiple testing. Journal of the Royal Statistical Society: Series B (Methodological) 57, 289–300. doi:10.1111/j.2517-6161.1995.tb02031.x

Bhattacharya, D., Sharma, N. K., Dutta, D., Chetupalli, S. R., Mote, P., Ganapathy, S., et al. (2023). Coswara: A respiratory sounds and symptoms dataset for remote screening of SARS-CoV-2 infection. Scientific Data 10, 397. doi:10.1038/s41597-023-02266-0. 2,635 subjects (2023 Scientific Data paper; cited per author decision Q3).

Botha, G. H. R., Theron, G., Warren, R. M., Klopper, M., Dheda, K., van Helden, P., et al. (2018). Detection of tuberculosis by automatic cough sound analysis. Physiological Measurement 39, 045005. doi:10.1088/1361-6579/aab6d0. 38 patients.

Brown, C., Chauhan, J., Grammenos, A., Han, J., Hasthanasombat, A., Spathis, D., et al. (2020). Exploring automatic diagnosis of COVID-19 from crowdsourced respiratory sound data. In Proceedings of the 26th ACM SIGKDD International Conference on Knowledge Discovery & Data Mining (KDD ‘20) (Association for Computing Machinery), 3474–3484. doi:10.1145/3394486.3412865

Centers for Disease Control and Prevention (2019). Antibiotic Resistance Threats in the United States, 2019. Tech. rep., U.S. Department of Health and Human Services, CDC, Atlanta, GA. doi:10.15620/cdc:82532

[Dataset] Chaudhari, G., Jiang, X., Fakhry, A., Han, A., Xiao, J., Shen, S., et al. (2020). Virufy: Global applicability of crowdsourced and clinical datasets for AI detection of COVID-19 from cough. doi:10.48550/arXiv.2011.13320. Open cough subset = 121 recordings. https://github.com/virufy/virufy-data

Cohen-McFarlane, M., Goubran, R., and Knoefel, F. (2020). Novel coronavirus cough database: NoCoCoDa. IEEE Access 8, 154087–154094. doi:10.1109/ACCESS.2020.3018028. 73 cough events (workflow-confirmed); coughs segmented from public media interviews.

Çorbacıoğlu, Ş. K. and Aksel, G. (2023). Receiver operating characteristic curve analysis in diagnostic accuracy studies: A guide to interpreting the area under the curve value. Turkish Journal of Emergency Medicine 23, 195–198. doi:10.4103/tjem.tjem_182_23

de Boer, G. and Bressmann, T. (2016). Application of linear discriminant analysis to the long-term averaged spectra of simulated disorders of oral-nasal balance. The Cleft Palate-Craniofacial Journal 53, e163–e171. doi:10.1597/14-236. Hyponasal/oral-nasal-balance disorders produce distinctive long-term averaged spectra (spectral-balance change). PMID 26068387.

Eyben, F., Scherer, K. R., Schuller, B. W., Sundberg, J., André, E., Busso, C., et al. (2016). The Geneva minimalistic acoustic parameter set (GeMAPS) for voice research and affective computing. IEEE Transactions on Affective Computing 7, 190–202. doi:10.1109/TAFFC.2015.2457417

Eyben, F., Wöllmer, M., and Schuller, B. (2010). opensmile: The munich versatile and fast open-source audio feature extractor. In Proceedings of the 18th ACM International Conference on Multimedia (MM ‘10) (Association for Computing Machinery), 1459–1462. doi:10.1145/1873951.1874246

Finkelstein, Y., Bar-Ziv, J., Nachmani, A., Berger, G., and Ophir, D. (1993). Peritonsillar abscess as a cause of transient velopharyngeal insufficiency. The Cleft Palate-Craniofacial Journal 30, 421–428. doi:10.1597/1545-1569_1993_030_0421_paaaco_2.3.co_2. Tonsillar/pharyngeal mass effect on oropharyngeal resonance. CAVEAT: describes the severe (peritonsillar abscess) end.

Fleming-Dutra, K. E., Hersh, A. L., Shapiro, D. J., Bartoces, M., Enns, E. A., File, T. M., et al. (2016). Prevalence of inappropriate antibiotic prescriptions among US ambulatory care visits, 2010-2011. JAMA 315, 1864–1873. doi:10.1001/jama.2016.4151. Acute respiratory conditions = 44% of outpatient antibiotic prescriptions; 50% of antibiotics for those conditions unnecessary; 30% of all outpatient antibiotics inappropriate.

Goncalves, L., Salman, A. N., Naini, A. R., Moro-Velázquez, L., Thebaud, T., Garcia, P., et al. (2024). Odyssey 2024 - speech emotion recognition challenge: Dataset, baseline framework, and results. In The Speaker and Language Recognition Workshop (Odyssey 2024). 247–254. doi:10.21437/odyssey.2024-35

Hammond, J., Leister-Tebbe, H., Gardner, A., Abreu, P., Bao, W., Wisemandle, W., et al. (2022). Oral nirmatrelvir for high-risk, nonhospitalized adults with Covid-19. New England Journal of Medicine 386, 1397–1408. doi:10.1056/NEJMoa2118542. EPIC-HR trial: nirmatrelvir-ritonavir (Paxlovid) started within 5 days of symptom onset.

Huddart, S., Geocaniga-Gaviola, D. M., Crowder, R., Ardrey, A., Theron, G., et al. (2024). A dataset of solicited cough sound for tuberculosis triage testing. Scientific Data 11, 1149. doi:10.1038/s41597-024-03972-z. 733,756 cough sounds / 2,143 participants / 565 longitudinal. Verify full author list at DOI.

Jiang, R.-S. and Huang, H.-T. (2006). Changes in nasal resonance after functional endoscopic sinus surgery. American Journal of Rhinology 20, 432–437. doi:10.2500/ajr.2006.20.2882. Nasalance (hyponasality) measured in chronic rhinosinusitis (n=81). CAVEAT: chronic + prompted speech, not acute J01 spontaneous speech. PMID 16955774.

Kaur, S., Larsen, E., Harper, J., Purandare, B., Uluer, A., Hasdianda, M. A., et al. (2023). Development and validation of a respiratory-responsive vocal biomarker–based tool for generalizable detection of respiratory impairment: Independent case-control studies in multiple respiratory conditions including asthma, chronic obstructive pulmonary disease, and COVID-19. Journal of Medical Internet Research 25, e44410. doi:10.2196/44410. Validation cohort 4,936 (asthma 1,694 + COPD 625 + persistent cough 814 + ILD 98 + healthy 1,705); COVID-19 is a separate 283-analyzed/497-enrolled case-control study. 6-sec sustained vowel.

Koç, E. A. Ö., Koç, B., and Erbek, S. (2014). Comparison of acoustic and stroboscopic findings and voice handicap index between allergic rhinitis patients and controls. Balkan Medical Journal 31, 340–344. doi:10.5152/balkanmedj.2014.14511. Non-infectious (allergic rhinitis) upper-airway disease produces measurable voice change.

Liao, L., Song, W., Wang, S., and Wang, X. (2022). A classification framework for identifying bronchitis and pneumonia in children based on a small-scale cough sounds dataset. PLOS ONE 17, e0275479. doi:10.1371/journal.pone.0275479. Cough-acoustic bronchitis-vs-pneumonia in 173 children, 86% accuracy. PMC9612535.

Loshchilov, I. and Hutter, F. (2017). SGDR: Stochastic gradient descent with warm restarts. In International Conference on Learning Representations (ICLR)

Loshchilov, I. and Hutter, F. (2019). Decoupled weight decay regularization. In International Conference on Learning Representations (ICLR)

Lyu, Y., Jiang, Q.-C., Yuan, S., Hong, J., Chen, C.-F., Wu, H.-M., et al. (2025). Non-invasive acoustic classification of adult asthma using an XGBoost model with vocal biomarkers. Scientific Reports 15, 28682. doi:10.1038/s41598-025-14645-1. Asthma vocal-biomarker classification; pair with lri-breath-support-prosody (COPD).

McFee, B., Raffel, C., Liang, D., Ellis, D. P. W., McVicar, M., Battenberg, E., et al. (2015). librosa: Audio and music signal analysis in python. In Proceedings of the 14th Python in Science Conference (SciPy 2015). 18–24. doi:10.25080/Majora-7b98e3ed-003

Muguli, A., Pinto, L., Nirmala, R., Sharma, N., Krishnan, P., Ghosh, P. K., et al. (2021). DiCOVA challenge: Dataset, task, and baseline system for COVID-19 diagnosis using acoustics. In Proc. Interspeech 2021. 901–905. doi:10.21437/Interspeech.2021-74

[Dataset] National Center for Health Statistics (2025). Preliminary estimates of visits to health centers in the united states, january 2022–december 2025. NAMCS Health Center Component, data.cdc.gov dataset 367e-pucc. Lung- and breathing-related (respiratory) diagnosis chapter accounted for 10.6% to 13.5% of health center visits across biannual periods, 2022–2025 (author calculation from visit counts).

Olson, K. D., Meeker, D., Troup, M., Barker, T. D., Nguyen, V. H., Manders, J. B., et al. (2025). Use of ambient ai scribes to reduce administrative burden and professional burnout. JAMA Network Open 8, e2534976. doi:10.1001/jamanetworkopen.2025.34976

Orlandic, L., Teijeiro, T., and Atienza, D. (2021). The COUGHVID crowdsourcing dataset, a corpus for the study of large-scale cough analysis algorithms. Scientific Data 8, 156. doi:10.1038/s41597-021-00937-4. Over 25,000 crowdsourced cough recordings.

Pahar, M., Klopper, M., Reeve, B., Warren, R., Theron, G., and Niesler, T. (2021a). Automatic cough classification for tuberculosis screening in a real-world environment. Physiological Measurement 42, 105014. doi:10.1088/1361-6579/ac2fb8. 51 subjects / 1,358 forced cough recordings.

Pahar, M., Klopper, M., Warren, R., and Niesler, T. (2021b). COVID-19 detection in cough, breath and speech using deep transfer learning and bottleneck features. Computers in Biology and Medicine 141, 105153. doi:10.1016/j.compbiomed.2021.105153. COVID-19 detection from cough/breath/SCRIPTED speech (counting, fixed utterance); speech AUC 0.92, less informative than cough. Crowdsourced + small clinical validation; pre-Omicron. (Distinct from pahar-tb-cough-2021; renders as Pahar et al. 2021a/b.)

Pimenta, J., Macedo, J., de Rezende Neto, A. L., and de Moraes Marchiori, L. L. (2022). Sensation and repercussion of the use of humid heat in the treatment of dysphonia due to laryngitis in singers. Journal of Voice 38, 496–502. doi:10.1016/j.jvoice.2021.09.038

Porter, P., Abeyratne, U., Swarnkar, V., Tan, J., Ng, T.-W., Brisbane, J. M., et al. (2019a). A prospective multicentre study testing the diagnostic accuracy of an automated cough sound centred analytic system for the identification of common respiratory disorders in children. Respiratory Research 20, 81. doi:10.1186/s12931-019-1046-6

Porter, P., Abeyratne, U., Swarnkar, V., Tan, J., Ng, T.-w., Brisbane, J. M., et al. (2019b). A prospective multicentre study testing the diagnostic accuracy of an automated cough sound centred analytic system for the identification of common respiratory disorders in children. Respiratory Research 20, 81. doi: 10.1186/s12931-019-1046-6. Automated cough analysis identifying common respiratory disorders incl. pneumonia (pediatric).

Renjini, A., Swapna, M. S., Raj, V., and Sankararaman, S. (2021). Graph-based feature extraction and classification of wet and dry cough signals: a machine learning approach. Journal of Complex Networks 9, cnab039. doi:10.1093/comnet/cnab039. Wet-vs-dry cough classification, framed as bacterial-vs-viral proxy; classifies cough morphology, NOT validated etiology. Cite to acknowledge adjacent work; the validated bacterial-vs-viral acoustic gap (adults) remains.

Sara, J. D. S., Orbelo, D., Maor, E., Lerman, L. O., and Lerman, A. (2023). Guess what we can hear—novel voice biomarkers for the remote detection of disease. Mayo Clinic Proceedings 98, 1353–1375. doi:10.1016/j.mayocp.2023.03.007. Review of voice-biomarker disease mechanisms incl. infectious/respiratory disease (B00 umbrella).

Shah, S. J., Devon-Sand, A., Ma, S. P., Jeong, Y., Crowell, T., Smith, M., et al. (2025). Ambient artificial intelligence scribes: physician burnout and perspectives on usability and documentation burden. Journal of the American Medical Informatics Association 32, 375–380. doi:10.1093/jamia/ocae295

Sharan, R. V., Qian, K., and Yamamoto, Y. (2024). Automated cough sound analysis for detecting childhood pneumonia. IEEE Journal of Biomedical and Health Informatics 28, 193–203. doi:10.1109/JBHI.2023.3327292. Distinguishes pneumonia from other acute respiratory diseases via cough (pediatric).

Sharma, M., Nduba, N., Bishai, W., Tahamont, S., et al. (2024). TBscreen: A passive cough classifier for tuberculosis screening with a controlled dataset. Science Advances 10, eadi0282. doi:10.1126/sciadv.adi0282. 195 subjects; 34,600 cough events (33k passive + 1.6k forced). Verify full author list at DOI.

Sharma, N. K., Chetupalli, S. R., Bhattacharya, D., Dutta, D., Mote, P., and Ganapathy, S. (2022). The second DiCOVA challenge: Dataset and performance analysis for diagnosis of COVID-19 using acoustics. In Proc. IEEE International Conference on Acoustics, Speech and Signal Processing (ICASSP). 556–560. doi:10.1109/ICASSP43922.2022.9747188. 1,436 subjects (965 development + 471 evaluation).

Shimon, C., Shafat, G., Dangoor, I., and Ben-Shitrit, A. (2021). Artificial intelligence enabled preliminary diagnosis for COVID-19 from voice cues and questionnaires. The Journal of the Acoustical Society of America 149, 1120–1124. doi:10.1121/10.0003434. Scripted-speech (vowel /a/) + cough COVID-19 classification, pre-Omicron.

Singh, H., Giardina, T. D., Meyer, A. N. D., Forjuoh, S. N., Reis, M. D., and Thomas, E. J. (2013). Types and origins of diagnostic errors in primary care settings. JAMA Internal Medicine 173, 418–425. doi:10.1001/jamainternmed.2013.2777. Pneumonia = most common single missed diagnosis (6.7%) in primary-care diagnostic errors.

Uyeki, T. M., Bernstein, H. H., Bradley, J. S., Englund, J. A., File, T. M., Fry, A. M., et al. (2019). Clinical practice guidelines by the infectious diseases society of america: 2018 update on diagnosis, treatment, chemoprophylaxis, and institutional outbreak management of seasonal influenza. Clinical Infectious Diseases 68, e1–e47. doi:10.1093/cid/ciy866. Influenza antiviral treatment most beneficial within 48 h of symptom onset.

Weglarz, K., Szczygiel, E., Maslon, A., and Blaut, J. (2025). Assessment of breathing patterns and voice of patients with COPD and dysphonia. Respiratory Medicine 240, 108012. doi:10.1016/j.rmed.2025.108012

Xia, T., Spathis, D., Brown, C., Chauhan, J., Grammenos, A., Han, J., et al. (2021). COVID-19 sounds: A large-scale audio dataset for digital respiratory screening. In Proceedings of the 35th Conference on Neural Information Processing Systems (NeurIPS) Datasets and Benchmarks Trac*k*. 36,116 participants.

Yan, Y., van Bemmel, L., Franssen, F. M. E., Simons, S. O., and Urovi, V. (2025). Developing a multi-feature fusion model for exacerbation classification in asthma and COPD. Computer Methods and Programs in Biomedicine 268, 108796. doi:10.1016/j.cmpb.2025.108796. Speech-based exacerbation classification in asthma and COPD (TACTICAS dataset), MFCC + multi-domain feature fusion, accuracy 0.892 / AUC 0.955. PMID 40347619.

Youden, W. J. (1950). Index for rating diagnostic tests. Cancer 3, 32–35. doi:10.1002/1097-0142(1950)3:1⟨32::AID-CNCR2820030106⟩3.0.CO;2-3

Zimmer, A. J., Espinoza-Lopez, P., Ravi, V., et al. (2026a). External validation of cough-based algorithms for pulmonary tuberculosis screening from the CODA TB DREAM challenge using cough data from Peru. Scientific Reports 16, 50492. doi:10.1038/s41598-026-50492-4

Zimmer, A. J., Ravi, V., Espinoza-Lopez, P., Kafentzis, G. P., Ravanelli, M., Abbasgholizadeh Rahimi, S., et al. (2026b). Cough acoustic analysis using artificial intelligence for COVID-19 detection: a comparative study of patient cohorts from Lima, Peru and Montreal, Canada. Annals of Epidemiology 118, 110076. doi:10.1016/j.annepidem.2026.110076

