## Supplementary data description for "Conversational Speech for Respiratory Triage in Primary Care: A Pilot Study"

### Supplementary Material

#### 1 DATA DISTRIBUTION OF THE ANALYTIC SAMPLE

The analytic sample comprises 514,377 visits from 379,225 unique adult patients. The following subsections describe its demographic, geographic, clinical, and audio-duration distributions.

##### 1.1 Demographic distribution

Median patient age at the time of visit is 38 years, with an interquartile range of 28 to 55 years and a mean of 42.7 years. The age range spans 18 to 106 years. The distribution is right-skewed (Figure S1A), peaking in the 25 to 29 bin and decaying monotonically thereafter. Sex distribution across the 514,377 analytic-sample visits is 326,412 (63.5%) female, 187,884 (36.5%) male, 72 (<0.1%) Unknown, and 9 (<0.1%) Other. The female skew is consistent with primary care utilization patterns reported in US ambulatory care surveys (National Center for Health Statistics, 2017) and is not a recruitment artifact of this work. Patient-level deduplication (n=379,225) yields the same median age (38), an interquartile range of 28 to 55, and a sex distribution within 0.1% of the visit-level numbers, indicating that the visit-level view is not distorted by a small number of high-frequency patients. The longitudinal structure of the sample is sparse (Figure S1B): 77.1% of patients (n=292,422) contributed a single visit, 15.4% (n=58,322) contributed two, 6.9% (n=26,328) contributed three to five, and 0.5% (n=2,064) contributed six to ten. A small tail of 89 patients (0.02%) contributed between 11 and 25 visits, and no patient in the sample exceeded 25 visits.

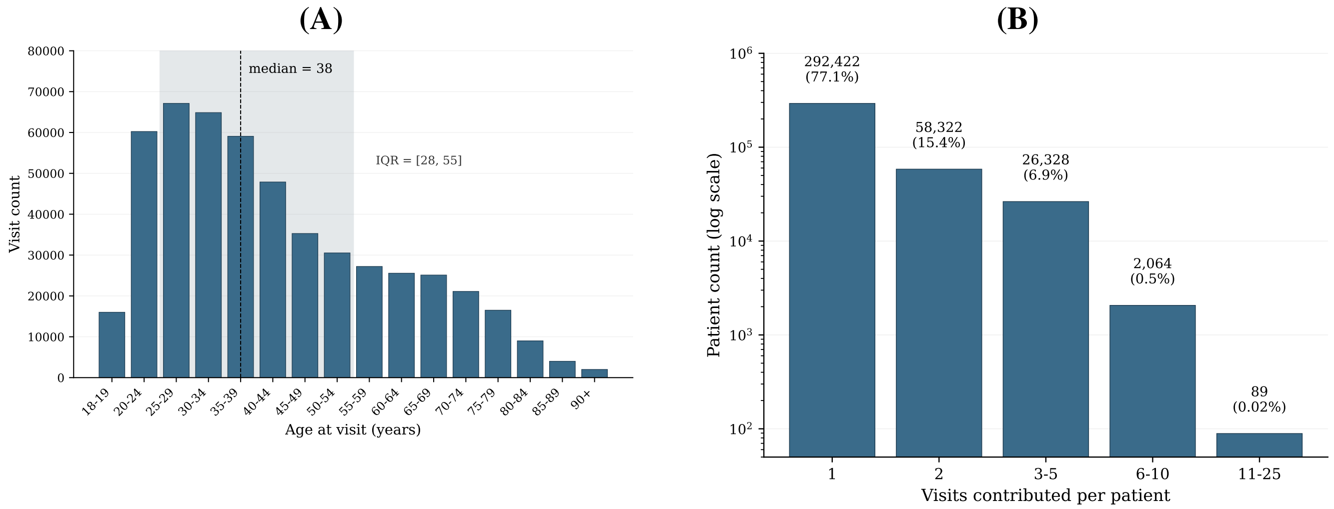


**Figure S1.** Demographic distribution of the analytic sample (n=514,377 visits from 379,225 unique patients). **(A)** Patient age distribution at the time of visit, binned in 5-year intervals; the distribution is right-skewed and peaks in the 25 to 29 bin. **(B)** Visits-per-patient distribution; 77.1% of patients contributed a single visit and no patient contributed more than 25.

##### 1.2 Geographic distribution

Geographic information is recorded at the patient level as the state and city of the patient's address. The analytic sample spans all 50 US states, the District of Columbia, and four US territories (Puerto Rico, the US Virgin Islands, Guam, and the Northern Mariana Islands), with a small number of visits also recorded from US military addresses and from international jurisdictions where the network's patients were temporarily resident. State and city fields were normalized prior to the analysis below: full-name spellings of states were merged into their two-letter codes, and visits with unparseable state values (single characters, punctuation, numeric sentinels, and similar artifacts, totaling approximately 0.17% of the sample) were excluded from the geographic counts but retained in all other analyses. The resulting distribution is heavily concentrated in California and the Northeast (Figure S2A): California alone accounts for 58.4% of visits (n=300,229), followed by New Jersey at 15.6% (n=80,350), Arizona at 5.3% (n=27,383), Washington at 4.2% (n=21,348), and Massachusetts at 3.4% (n=17,571). These five states together account for 86.9% of the sample, and the top ten states account for 95.3%. At the city level, the sample spans 9,251 unique cities, with the most heavily represented being Los Angeles, San Francisco, Tucson, San Jose, and Simi Valley (Figure S2B). The long tail is substantial: 3,800 cities (41% of unique cities) contributed a single visit each.

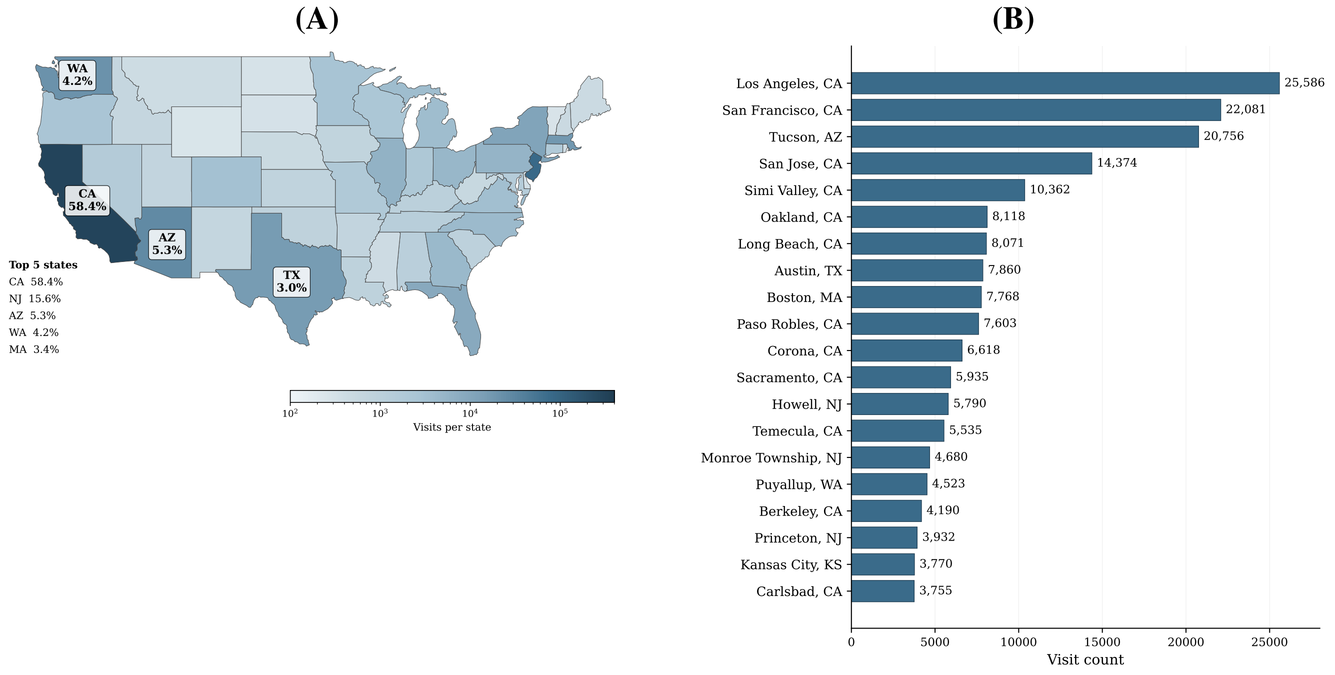


**Figure S2.** Geographic distribution of the analytic sample (n=514,377 visits from 379,225 unique patients). **(A)** State-level visit counts; California accounts for 58.4% of visits and the top five states (California, New Jersey, Arizona, Washington, Massachusetts) account for 86.9%. **(B)** City-level visit distribution across 9,251 unique cities; the long tail comprises 3,800 cities (41%) that contribute a single visit each.

#### 1.3 Clinical distribution

The clinical composition of the analytic sample is summarized by the distribution of primary ICD-10 diagnosis codes assigned to each visit. At the chapter level, respiratory diagnoses (Chapter J) account for 52.3% of all primary codes (Figure S3A), which is the direct consequence of the cohort construction described in the main paper: visits are included in the analytic sample if and only if they satisfy the inclusion or exclusion criteria of at least one of the 11 binary classification tasks, and most of those tasks anchor on respiratory ICD-10 codes. The non-respiratory share is dominated by symptoms and signs (Chapter R, 12.74%), musculoskeletal complaints (Chapter M, 10.64%), genitourinary visits (Chapter N, 8.49%), and skin conditions (Chapter L, 6.20%). The U chapter (special purposes), which contains the U07.1 code for confirmed COVID-19, contributes 2.92%. All remaining chapters together account for less than 7% of visits. This distribution reflects the analytic sample, not the originating primary care population at large; the upstream selection into the 11 binaries deliberately enriches for respiratory presentations and for non-respiratory acute-illness negatives.

At the individual-code level (Figure S3B), the analytic sample is dominated by acute upper respiratory presentations. J06.9 (acute upper respiratory infection, unspecified) alone accounts for 19.74% of all visits, followed by J02.9 (acute pharyngitis, unspecified) at 9.33%. Sinusitis codes (J01.90 and J01.00) together contribute 8.43%. The top 20 primary codes (each representing at least 1% of visits) collectively account for 71.07% of the analytic sample. Notable non-respiratory codes in the top 20 are N39.0 (urinary tract infection, 7.34%), N30.01 (acute cystitis with hematuria, 1.31%), R30.0 (dysuria, 1.62%), M54.50 (low back pain, 1.67%), R10.9 (unspecified abdominal pain, 1.07%), and R07.0 (pain in throat, 1.17%); these visits are present in the analytic sample because they serve as acute non-respiratory negative controls for one or more of the binary classification tasks. The prominence of "unspecified" codes (J06.9, J02.9, J01.90, J22, J18.9, J20.9, R10.9, R07.0, J45.901) reflects the reality of primary care coding, where clinicians frequently assign the least-specific code consistent with the presentation.

Patient-level documentation of chronic conditions and comorbidities is available for the analytic sample through the full ICD-10 code arrays attached to each patient's visit history. Documented prevalences across the analytic sample of 379,225 patients are 4.69% for chronic respiratory disease (defined as any occurrence of J44, J45, J47, J84, or E84), 1.35% for anxiety (F41), 0.93% for gastroesophageal reflux disease (K21), 0.59% for depression (F32 or F33), 0.33% for obesity (E66), 0.12% for tobacco use or dependence (F17, Z72.0, or Z87.891), 0.09% for obstructive sleep apnea (G47.3x), and 0.04% for heart failure (I50). These prevalences are computed against the full denominator of 379,225 patients, treating absence of the code as absence of the condition. They are therefore documentation prevalences rather than true population prevalences, and they undercount by construction: a condition that exists but has not been coded during any visit in the observation window does not contribute. The tobacco use figure of 0.12% is the clearest example, since US adult tobacco use prevalence is approximately 14% (Arrazola et al., 2025), and the present figure is best read as a coding-rate floor rather than a true-prevalence estimate.

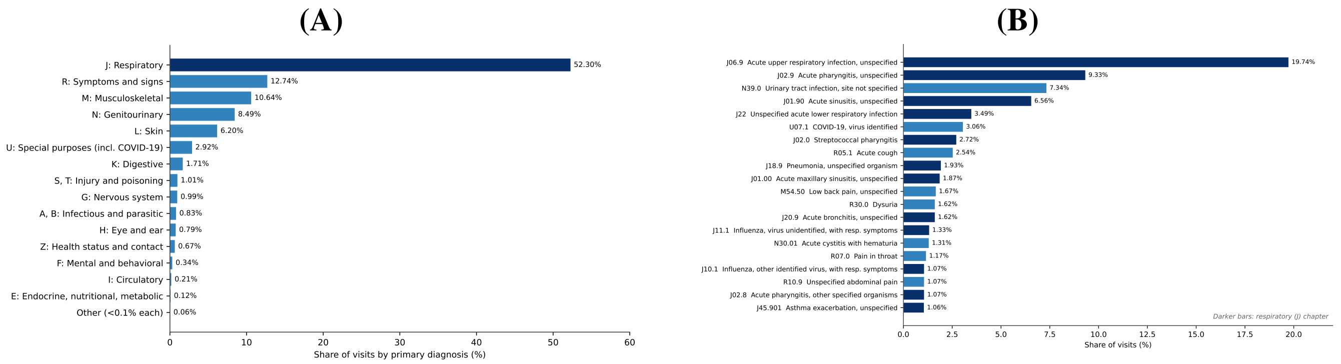


**Figure S3.** Clinical distribution of primary ICD-10 codes across the analytic sample (n=514,377 visits from 379,225 unique patients). **(A)** ICD-10 chapter distribution; the respiratory chapter (J) accounts for 52.3% of primary diagnoses, followed by symptoms and signs (R, 12.74%), musculoskeletal (M, 10.64%), genitourinary (N, 8.49%), and skin (L, 6.20%). **(B)** Top primary ICD-10 codes; the top 20 codes account for 71.07% of visits, led by J06.9 (acute upper respiratory infection, unspecified) at 19.74% and J02.9 (acute pharyngitis, unspecified) at 9.33%.

#### 1.4 Audio duration distribution

Patient audio duration is the length of the patient-channel signal extracted from each recording by the diarization pipeline described in the main paper; it is the input length to the feature extractor for that visit. Across the 514,377 visits in the analytic sample, the median duration is 27.8 seconds with an interquartile range of 15.4 to 47.2 seconds, the mean is 36.8 ± 34.1 seconds, and the range is 0.7 to 776 seconds (Figure S4). The distribution is unimodal and right-skewed on a linear scale; on the log scale of Figure S4 it appears roughly symmetric. A small accumulation near the one-second boundary corresponds to a minor leak in the audio-quality filter, where 3,542 visits (0.7%) recorded a duration below the nominal one-second floor; this leak is too small to bias any downstream analysis but we note it for transparency. The total measured patient audio across the analytic sample is 5,258 hours.

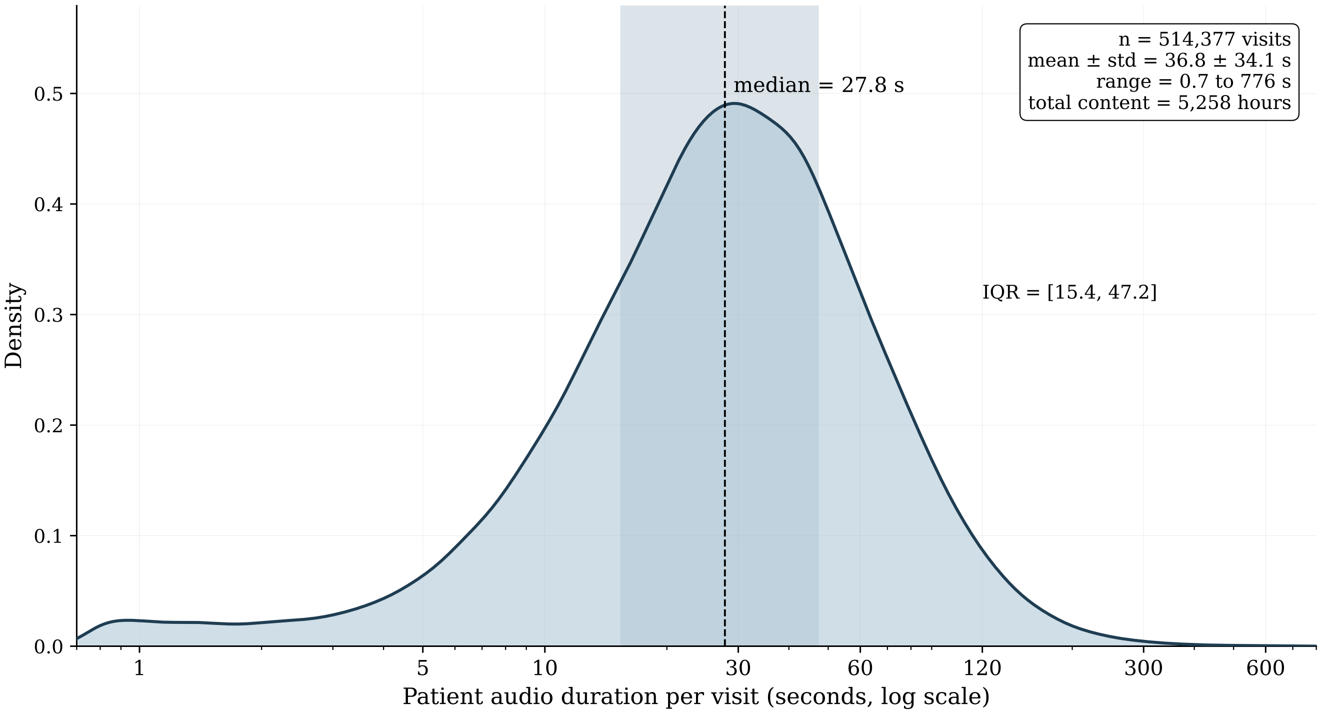


**Figure S4.** Patient-channel audio duration across the analytic sample (n=514,377 visits). The distribution is unimodal and right-skewed on a linear scale (median 27.8 s, interquartile range 15.4 to 47.2 s, mean 36.8 ± 34.1 s, range 0.7 to 776 s); on the log scale shown here it appears roughly symmetric. The total measured patient audio is 5,258 hours.

### 2 MODELING AND EVALUATION DETAILS

#### 2.1 Diarization configuration

The full-recording audio for each visit is submitted to Google Cloud Speech-to-Text v2 using the Chirp 3 acoustic model in its BatchRecognize mode, with one to two expected speakers, English as the recognition language, and word-level timestamps as the output granularity. Words from consecutive same-speaker positions are merged into per-speaker segments, with speaker labels normalized to SPEAKER_01 and SPEAKER_02. To identify which of the diarized speakers is the patient, the diarized transcript is passed to the gemini-flash-3.1-lite large language model with a deterministic system prompt that asks for the speaker label of the primary subject of the visit, defined as the patient or person under evaluation rather than a caregiver, translator, automated system, or any other participant. The classifier returns either a speaker label or a sentinel value indicating that no patient is present in the recording, in which case the visit is excluded from the pipeline. Patient-only audio is then reconstructed by concatenating, in chronological order, the time-aligned slices of the original recording corresponding to the chosen speaker label, producing a single contiguous WAV file containing only the patient's voice.

The signal entering feature extraction is the patient-only WAV at the rate produced by the diarization pipeline (16 kHz). When the source audio is stereo or multi-channel, channels are averaged to mono before processing; the source data for this work is single-channel by construction. No loudness or amplitude normalization, no denoising, no voice activity detection beyond the diarization-based speaker labels, and no silence trimming beyond the segment-level cuts produced by diarization are applied.

#### 2.2 Feature extraction details

One set of six streams is composed of 88 features that are produced by the openSMILE eGeMAPSv02 Functionals feature set (computed at 16 kHz sampling rate using the published eGeMAPSv02 configuration of frame-level low-level descriptors aggregated into clip-level functionals such as mean, normalized standard deviation, percentiles, percentile ranges, and rising and falling slope statistics). The 88 outputs are partitioned into six streams aligned with the components of the speech production apparatus: pitch (10 features, F0-based), loudness (10 features, perceptual loudness statistics), spectral (28 features including MFCC1 to MFCC4 [Mel-frequency cepstral coefficients], spectral flux, the Hammarberg index, and voiced and unvoiced spectral slopes), voice quality (10 features covering jitter, shimmer, and harmonics-to-noise ratio), formants (18 features covering F1, F2, and F3 frequencies, bandwidths, and amplitudes relative to F0), and temporal (12 features including voiced and unvoiced segment statistics, loudness peaks per second, and equivalent sound level).

One stream contains 281 features derived from librosa (version 0.11.0), summarizing seven families of low-level descriptors over the whole patient clip: Mel-frequency cepstral coefficients (13 bands with nine summary statistics each), chromagram (12 pitch classes with nine statistics each), spectral descriptors (centroid, bandwidth, contrast, flatness, rolloff, zero-crossing rate), energy features (root-mean-square), spectral contrast, tonnetz on the harmonic component, and rhythm features (tempo and predominant local pulse). Librosa frame-level extraction uses the library defaults of a 2048-sample FFT window, a 512-sample hop, and a 2048-sample analysis window, which at the 16 kHz sampling rate corresponds to a 128 ms window and 32 ms hop. Each per-frame low-level descriptor is summarized into nine statistics (mean, standard deviation, minimum, maximum, median, 25th and 75th percentiles, interquartile range, and range), and single-scalar features such as tempo are stored directly.

One stream contains 24 features derived from a valence, arousal, and dominance predictor (WavLM-VAD-Odyssey2024), computed as per-clip distribution statistics of the three predicted axes. One stream contains 19 features from a temporal-analysis extractor that combines praat-parselmouth syllable-and-pause detection (yielding total syllables, total pauses, total file duration, phonation time, speech rate, articulation rate, average syllable duration, total pause duration, and maximum, minimum, and mean pause durations) with librosa tempo statistics (mean, median, standard deviation, minimum, maximum, interquartile range, coefficient of variation, and range).

Non-numeric columns and extraction-metadata columns are dropped at the consolidation step, and visits whose source feature parquet is missing one or more required columns are dropped at compact-build time; this affects approximately 3 to 5% of the visits per binary and is recorded in a sidecar audit file.

#### 2.3 Model architecture details

The nine feature streams are concatenated in a fixed order (valence-arousal-dominance, pitch, loudness, spectral, voice quality, formants, temporal, temporal analysis, librosa) before being passed to the network.

**Table S1.** Training and hyperparameter settings for the single-stream feed-forward classifier, applied per binary.

| Setting | Value |
|---|---|
| Class balancing | minority class $\times 2$, patient-level downsampling, seeded |
| Test hold-out | 30% of unique patients (patient-level, stratified) |
| Cross-validation | 5-fold, patient-level, stratified |
| Optimizer | AdamW, weight decay 0.001 |
| Learning-rate schedule | cosine annealing, no warmup |
| Gradient clipping | maximum norm 1.0 |
| Minibatch size | 256 |
| Maximum epochs | 50 |
| Early stopping | patience 10 on development loss |
| Learning-rate grid | 1e-4, 3e-4, 1e-3, 3e-3 |
| Architecture grid | [128, 64]; [256, 128, 64]; [256, 128, 64, 32]; [512, 256, 128] |
| Hidden block | linear, batch normalization, GELU, dropout 0.5 |
| Decision threshold | Youden's J on the development partition |

The network consists of two to four hidden blocks followed by a final linear projection to a single output logit. Each hidden block is a sequence of a linear layer, a one-dimensional batch normalization, a GELU activation, and a dropout layer with rate 0.5. The final linear layer projects the last hidden block's activations to a single logit, with no activation; sigmoid is applied at inference time when probabilities are required.

##### 2.4 Training hyperparameters

For each binary, balancing reduces the analytic sample to twice the size of the minority class. The minority class is taken in full at the visit level. The majority class is downsampled by sampling unique patients without replacement until the expected visit count matches the minority size; if the realized visit count exceeds the target, the class is trimmed to exact size at the visit level. Sampling is seeded for reproducibility.

Each model is trained for up to 50 epochs with AdamW (weight decay 0.001), a cosine-annealing learning rate schedule with no warmup, and gradient norm clipping at 1.0. The minibatch size is 256. Training halts early when development loss has not improved for 10 consecutive epochs, and the checkpoint with the lowest development loss is retained as that fold's final model.

The learning rate and the hidden-layer topology are chosen per binary by a grid search over four learning rates (1e-4, 3e-4, 1e-3, 3e-3) and four candidate topologies (two-block at 128 and 64; three-block at 256, 128, 64; four-block at 256, 128, 64, 32; three-block at 512, 256, 128), yielding 16 combinations per binary. Each combination is trained once on a single development split of the balanced cohort, and the combination achieving the lowest development loss is selected. The selected combination is then retrained under five-fold cross-validation, producing five fold-specific models. Table S1 summarizes these settings.

#### 3 DETAILED COHORT DESIGN FOR THE 11-BINARY PANEL

Each binary in the panel was specified by a clinical brief, audited against the published ICD-10 / acoustic biomarker literature, and reduced to a SQL predicate. Predicates were finalised under a single *real-world cohort principle*: a visit's class membership is determined by the ICD-10 codes recorded on that visit, and is not extrapolated from clinical history. Patient-history exclusions were applied only where a pre-existing condition is known to shift the acoustic baseline at every subsequent visit (acute respiratory failure, neurological speech disease, post-intubation laryngeal injury). Codes were neither added to inflate a class size nor removed to maximise discriminability.

The same seven patient-level covariate flags are carried through every binary's manifest output (heart failure, obesity, obstructive sleep apnoea, depression, anxiety, gastro-oesophageal reflux disease, and smoking history) for downstream matching and subgroup AUC. These are carried as metadata only and are not used as exclusion criteria.

Per-binary specifications and the final review decisions that shaped the SQL predicate are summarised below.

#### 3.1 B00: Acute respiratory illness vs acute non-respiratory illness

B00 is the panel's clinical-triage anchor. The contrast was reformulated during review from its original form ("acute respiratory infection vs healthy non-illness visit") to its final form ("acute respiratory illness vs acute non-respiratory illness") so that three structural confounds (sick-affect envelope, conversation length, and audio-quality or recording-context skew) cancel by construction. Both arms are acute primary-care visits; any above-chance separation is therefore, by design, respiratory-mechanism specific rather than generic-sick-voice. The price of this reformulation is a lower attainable AUC ceiling (estimated 0.72–0.80 versus the original framing's 0.85–0.92), accepted for the stronger respiratory-specificity claim.

###### *3.1.0.1 Class definitions (final).*

The positive class is defined by a primary ICD-10 code in J00, J01, J02, J03, J06 (with J04 and J05 deliberately excluded for consistency with B03), J09–J11, J12–J22 with J12.82/J18.2 removed, pneumonia sequelae (J85, J86, J90, J91.8, J98.11), and acute COVID-19 (U07.1, J12.82). The negative class is defined by a primary ICD-10 code in acute gastrointestinal (A09, K52, K30, K59.0, K57, K35–K38, R10, R11), acute genitourinary (N30, N39.0, N20, N23), musculoskeletal (M54, M25.5, M79 excluding M79.7 fibromyalgia), headache (R51, G43, G44), acute skin (L02, L03), acute mental-health (F32, F41, F43.0), and acute symptom codes (R52, R50.9, R53.1), with the additional rule that the visit must carry no respiratory code at all. A patient-level exclusion drops anyone with a chronic respiratory diagnosis in history (J44, J45, J47, J84, E84). Final cohort: 265,210 positive and 188,663 negative visits (class ratio 1.41:1).

###### *3.1.0.2 Key review decisions.*

The visit-level respiratory-anywhere exclusion was widened to include R07.1 (painful respiration), which encodes a directly respiratory mechanic, dropping 27 visits from the negative class. F32 and F41 episode-codes were retained in the negative class despite their non-strictly-acute character: measured prevalence was 1.3% of the negative class, judged insufficient to warrant special handling, and consistent with a real-world clinical distribution. Seasonality matching was deferred to the fairness and robustness audit (which stratifies performance by month) rather than encoded in SQL, on the basis that the concern is theoretical for within-clinic audio inputs. A 90-day per-patient washout from prior respiratory visits was declined under the real-world cohort principle. A set of acute-by-ICD-definition negative sub-codes was added during review, growing the negative class by 12% (+20,210 visits); paired chronic-by-definition codes (F43.1 PTSD, F43.2 adjustment disorders, M79.7 fibromyalgia, chronic-fatigue R53.81–.83) were excluded to preserve the acute-vs-acute symmetry.

###### 3.1.0.3 Acknowledged limitations.

Depression somatisation (undiagnosed depression presenting with somatic complaints coded under back pain or fatigue) imports a depression-prosody signal into the negative arm that cannot be ICD-isolated; this is reported as a methods limitation.

##### 3.2 B01: Acute respiratory infection vs chronic respiratory disease management

B01 contrasts an acute infectious presentation against a chronic respiratory management visit. The negative arm pools obstructive (J44 COPD, J45 asthma), bronchiectatic (J47), interstitial (J84), and cystic-fibrosis (E84) chronic disease.

###### 3.2.0.1 Class definitions (final).

The positive class is defined by a primary ICD-10 code in URI (J00–J06), LRI (J12–J22), influenza (J09–J11), or COVID-19 (U07.1, U09, J12.82). The negative class is defined by a primary ICD-10 code in J44, J45, J47, J84, E84, J67 (hypersensitivity pneumonitis, added during review), or J70 (drug- or external-agent-induced lung disease, added during review), covering all subtypes including compound acute-on-chronic codes. A patient-level exclusion drops both arms when J96.x (acute respiratory failure) appears anywhere in history. A visit-level exclusion drops visits with both acute and chronic primary codes. Final cohort: 280,471 positive and 11,530 negative visits (class ratio 24.3:1).

###### 3.2.0.2 Key review decisions.

The dominant residual limitation is inhaled-corticosteroid (ICS) dysphonia in the negative arm: long-term ICS produces chronic vocal-fold edema (elevated jitter and shimmer, reduced harmonics-to-noise ratio) at high prevalence in the chronic-asthma and chronic-COPD population (Williamson et al., 1995), and an exposure proxy (Z79.51) is coded in only four patients in the entire dataset. The ICS confound is accepted as a primary limitation of B01 rather than mitigated, and is explicitly documented in the methods. J04.0 (acute laryngitis) and J06.0 (acute laryngopharyngitis) were retained in the positive arm despite producing the same acoustic signature as the chronic ICS dysphonia in the negative arm, under the real-world principle that they are genuine acute respiratory infections.

The negative-class compound acute-on-chronic problem was addressed by pre-registering two analysis paths. In Path A the pooled negative class (compound acute-on-chronic plus pure stable chronic) is reported as the primary endpoint; in Path B the negative class is restricted to `pure_stable_chronic` visits only ($n = 3{,}457$), and the AUC is reported alongside Path A. The interpretation is pre-specified: similar AUC across paths indicates a true respiratory-mechanism contrast independent of acute symptoms, whereas a substantially higher Path A AUC indicates that the contrast is being carried by the acute-event signature (cough, dyspnoea) shared between pure acute infections and acute exacerbations.

Patient-level chronic-respiratory history was kept in the positive arm (with stratified subgroup AUC by `pat_has_chronic_resp`) rather than excluded, both to preserve real-world prevalence and because cross-arm ICS exposure weakens the ICS-shortcut.

###### 3.2.0.3 Acknowledged limitations.

Smoking-status under-coding, and ICS-exposure unobservability, are documented as primary limitations.

#### 3.3 B02: Acute respiratory infection vs symptomatic non-infectious respiratory presentation

B02 contrasts an acute respiratory infection against a respiratory or non-respiratory complaint that is coded as non-infectious. The negative arm pools allergic rhinitis, chronic structural sinus disease, and respiratory symptom codes without an infection diagnosis.

###### *3.3.0.1 Class definitions (final).*

The positive class is defined by a primary ICD-10 code in J00–J06, J12–J22, J09–J11, or acute COVID-19 (U07.1, J12.82). The negative class is defined by a primary ICD-10 code in J30 (allergic rhinitis), J32 (chronic sinusitis), J33 (nasal polyp), R05 (cough family), R06 excluding R06.03 (dyspnoea, wheeze, stridor; acute respiratory distress R06.03 excluded as a severe acute state), or R07.0 (throat pain), with the additional rule that no acute respiratory infection code appears anywhere on the visit. Final cohort: 282,057 positive and 32,414 negative visits (class ratio 8.7:1).

###### *3.3.0.2 Key review decisions.*

Two implementation bugs flagged during review were corrected: U09.x (post-COVID condition, a chronic or post-acute sequela rather than acute infection) was removed from the positive predicate, dropping 587 net visits from the positive arm and ensuring U09.x visits are excluded from B02 entirely; and R06.03 (acute respiratory distress) was excluded from the negative predicate, dropping 28 visits, since it encodes a severe acute respiratory state rather than a non-infection symptom complaint. The negative class is explicitly heterogeneous (R05 cough $\sim$50%, J30 allergic rhinitis $\sim$18%, R06+R07.0 $\sim$24%, J32+J33 $\sim$11%). R05.3 (chronic cough) was retained in the negative class under the real-world principle, despite the possibility that some R05.3 visits represent post-infectious cough.

###### *3.3.0.3 Acknowledged limitations.*

Symptom-only R-code visits empirically include some under-coded infection (with documented under-coding precedent of $\sim$26% laboratory-confirmed-influenza ICD-coding sensitivity (Mira-Iglesias et al., 2025a)); the negative class is therefore stated as “coded as non-infectious” rather than “confirmed non-infectious.” Post-COVID dysphonia carryover from U09.x history is acknowledged but not mitigated beyond the same-visit U09.x rule.

#### 3.4 B03: Lower vs upper respiratory infection

B03 separates upper respiratory infection from lower respiratory infection within the acute-infection cohort. The contrast targets breath-support and cough-event differences associated with parenchymal or large-airway involvement.

###### *3.4.0.1 Class definitions (final).*

The positive (LRI) class is defined by a primary ICD-10 code in pneumonia (J12 excluding J12.82, J13–J17, J18 excluding J18.2), acute bronchitis (J20), bronchiolitis (J21), unspecified acute LRI (J22), influenza with pneumonia (J09.X1, J10.00/.01/.08, J11.00/.08), or pneumonia sequelae (J85, J86, J90, J91.8, J98.11). The negative (URI) class is defined by a primary ICD-10 code in J00, J01, J02, J03, or J06. Final cohort: 30,395 LRI and 189,988 URI visits (class ratio 1:6.25).

###### 3.4.0.2 Key review decisions.

J04.0 (acute laryngitis) was removed from the URI inclusion list, since it is a vocal-fold disease whose acoustic signature confounds an LRI-vs-URI separation built on breath-support and cough features. J22 (unspecified acute LRI) was retained in the LRI arm despite its known heterogeneity, with the caveat reported in the discussion; excluding it would have lost a large fraction of LRI volume that is real to primary-care coding practice. J06.9 (unspecified URI) primary visits were dropped from the URI arm when either R06.0x (dyspnoea) or R09.0x (hypoxaemia / asphyxia) was co-coded, since the combination signals an LRI workup in progress; this removed 571 visits (∼0.26%).

An asymmetric per-patient washout was applied: the LRI arm rejects any visit when the patient had a primary URI code in the prior 90 days, and the URI arm rejects any visit when the patient had a primary LRI code in the prior 180 days. The asymmetry reflects the longer duration of post-LRI respiratory residuals (cough and altered breath support persist 12+ weeks after pneumonia) compared to post-URI residuals (median 7–10 days, residual under 30 days). The washouts removed 3,215 LRI visits (∼9.0%) and 2,957 URI visits (∼1.4%).

A patient-level exclusion was applied symmetrically to both arms for J96.x (acute respiratory failure) in history, removing nine visits across 18 patients with altered respiratory-mechanic baselines. A second patient-level symmetric exclusion was applied for chronic upper-airway disease (J30–J35, J37–J39) which produces a stable resonance baseline shift, removing 1,888 LRI (∼5.26%) and 18,585 URI (∼8.6%) visits and resolving a prior asymmetry where only the URI arm carried this exclusion. A third patient-level symmetric exclusion was applied for neurological speech disease (G20 Parkinson's, G30 Alzheimer's, F03 dementia, I63 / I69 stroke and sequelae, G12.21 ALS, G35 MS, G70 myasthenia gravis) and post-intubation laryngeal injury (J95.0, J95.5); 49 visits across 236 patients were removed.

Five pneumonia-sequela codes (J85, J86, J90, J91.8, J98.11) were added to the LRI primary inclusion list, contributing 91 visits (∼0.3%), with equivalent codes propagated into the washout-window CTE. J40 and J42 (unspecified and chronic bronchitis) were added to the chronic-respiratory covariate flag rather than treated as exclusions, re-tagging 6,308 patients (2,020 LRI / 3,133 URI visits).

#### 3.5 B04: Pneumonia vs acute bronchitis

B04 separates pneumonia from acute bronchitis, the two most common LRI presentations in primary care. This is the most balanced binary in the panel.

###### 3.5.0.1 Class definitions (final).

The positive class is defined by a primary ICD-10 code in J12–J18 (any pneumonia). The negative class is defined by a primary ICD-10 code in J20 (acute bronchitis). A visit-level exclusion drops both arms when COVID-19 (U07.1, U09, J12.82) or influenza (J09–J11) is primary, since those visits are handled by B09 and B08 respectively; visits with both pneumonia and acute bronchitis in primary are also dropped as ambiguous. Final cohort: 11,254 pneumonia and 9,076 acute bronchitis visits (class ratio 1.24:1).

###### 3.5.0.2 Key review decisions.

J69 (aspiration pneumonia) and J68.0 (chemical pneumonitis) were already outside the positive predicate; patient-history exclusion for these (and for J95.851 ventilator-associated pneumonia history) was declined under the real-world cohort principle. J18.9 (unspecified pneumonia) was measured at 87% of the positive class, raising the concern that imaging-unconfirmed clinical-suspicion pneumonia contaminates the positive arm. J18.9 was retained under the real-world cohort principle. Mycoplasma cross-coding between J20.0 and J15.7 across a single episode was declined as a patient-history mitigation under the same principle.

The chronic opioid confound (cough-reflex suppression and shallow breathing) was acknowledged as a methods limitation rather than addressed via a B04-only covariate, since F11.x captures only opioid use disorder rather than chronic exposure, the same coding-fidelity limitation that applies to smoking. J85.1 (lung abscess with pneumonia) and J18.2 (hypostatic pneumonia) were measured at zero visits in the feature-available cohort and required no action.

#### 3.6 B05: Pneumonia vs non-pneumonia lower respiratory infection

B05 extends B04's contrast to the full non-pneumonia LRI population, adding bronchiolitis and unspecified acute LRI to the negative arm.

###### 3.6.0.1 Class definitions (final).

The positive class is defined by a primary ICD-10 code in J12–J18 (any pneumonia). The negative class is defined by a primary ICD-10 code in J20 (acute bronchitis), J21 (bronchiolitis, adult only after the feature-available filter), or J22 (unspecified acute LRI). The same COVID-19 and influenza visit-level exclusions as B04 apply, along with the dual-class exclusion. Final cohort: 10,968 pneumonia and 26,524 non-pneumonia LRI visits (class ratio 1:2.4), with the negative arm composed of 34% J20, $<$ 1% J21, and 66% J22.

###### 3.6.0.2 Key review decisions.

The dominant issue is J22 label noise. J22 (unspecified acute LRI) is less specific than J18.9, and literature on ancestor ICD-9 code 486 reports as low as 14.2% sensitivity for laboratory-confirmed pneumococcal pneumonia (Guevara et al., 1999). With J22 at 66% of the B05 negative class, an estimated 2,600–5,275 visits in the negative arm may be unrecognised pneumonia.

J17 (pneumonia in diseases classified elsewhere), J18.2 (hypostatic pneumonia), J95.851 (ventilator-associated pneumonia history), and J69 (aspiration pneumonia history) were either measured at zero visits in the feature-available cohort or declined under the real-world cohort principle.

#### 3.7 B06: Acute sinusitis vs other upper respiratory infection

B06 separates J01-coded acute sinusitis from other coded URI. It is explicitly reported as a secondary binary with a primary labelling-bias caveat.

###### 3.7.0.1 Class definitions (final).

The positive class is defined by a primary ICD-10 code starting with J01. The negative class is defined by a primary ICD-10 code starting with J00, J02, J03, J04, J05, or J06. A visit-level exclusion drops any LRI, COVID-19, or influenza primary code, holding the binary scope to URI only. Final cohort: 46,460 J01 and 166,037 other-URI visits (class ratio 1:3.6).

###### 3.7.0.2 Critical framing.

Approximately 10–15% of primary-care J01-coded visits represent true acute bacterial sinusitis (Chow et al., 2012); the remaining $\sim$85% are viral URI with sinus-symptom predominance that are clinically indistinguishable from a regular cold at the bedside. The contrast measured by B06 is therefore "ICD-coded acute sinusitis (predominantly viral URI with sinus-symptom presentation) vs other ICD-coded URI," not "bacterial sinusitis vs cold." An AUC near chance (0.55–0.65) is reported as a meaningful empirical negative result demonstrating that ICD-coded acute sinusitis is not acoustically separable from other ICD-coded URI in spontaneous primary-care conversation.

###### 3.7.0.3 Key review decisions.

The cohort was kept as the real-world J01-coded distribution and the manuscript framing was re-aligned to the labelling-bias caveat above. Hyponasality acoustic evidence used in the brief was downgraded from `grounded` to `plausible` where the cited paper studied chronic rhinosinusitis (J32) or nasal polyps (J33) after surgical intervention rather than acute J01; the manuscript states this evidence-translation gap explicitly. The 12-month look-back exclusion for chronic J32 history was declined under the real-world cohort principle; chronic-respiratory baseline shift is acknowledged as a methods limitation rather than a cohort exclusion. The odontogenic-maxillary-sinusitis sub-population ($\sim$10–30% of unilateral J01.00/.01) has no separate ICD code and is reported as residual variance.

#### 3.8 B07: GAS pharyngitis vs other pharyngitis

B07 separates Group A streptococcal pharyngitis (*Streptococcus pyogenes*) from other coded pharyngitis. The contrast is framed as "GAS vs non-GAS pharyngitis," not "bacterial vs viral pharyngitis."

###### 3.8.0.1 Class definitions (final).

The positive class is defined by a primary ICD-10 code of J02.0 (acute streptococcal pharyngitis). The negative class is defined by a primary ICD-10 code starting with J02 and not equal to J02.0 (i.e. J02.8 other specified and J02.9 unspecified). Visits with both J02.0 and other J02.x in primary are dropped as ambiguous. Final cohort: 12,094 J02.0 and 50,802 other-J02 visits (class ratio 1:4.2). A pre-specified go/no-go gate of $\geq 50$ J02.0 visits was cleared by a large margin.

###### 3.8.0.2 Critical framing.

The negative arm contains most viral pharyngitis (rhinovirus, adenovirus, parainfluenza), under-coded infectious mononucleosis (EBV) miscoded as J02.9, *Fusobacterium necrophorum* and other non-GAS bacterial causes (rarely tested in primary care), *Mycoplasma pneumoniae*, and *Chlamydophila pneumoniae*. The clinically more useful binary (bacterial vs viral pharyngitis) is not implementable with available ICD coding. The expected performance ceiling against a hypothetical RADT- or culture-confirmed gold standard is AUC 0.65–0.70 (Fine et al., 2012) per the brief's self-audit; an AUC closer to chance is reported as a meaningful empirical result rather than a model failure.

###### 3.8.0.3 Key review decisions.

The manuscript explicitly states that B07 targets the *milder* pharyngeal-cavity mass-effect signature (small F2 / F3 reductions on back vowels), not "hot potato voice," which is severe-end only (peritonsillar abscess, severe tonsillitis) and is not in the Centor or McIsaac clinical scoring criteria (Centor et al., 1981; McIsaac et al., 1998). B27.x (mononucleosis) patient-history exclusion was reaffirmed, and under-coded mononucleosis within J02.9 is acknowledged as irreducible contamination of the negative arm.

###### 3.8.0.4 Acknowledged limitations.

J02.0 label fidelity in adult primary care without RADT or culture confirmation (20–40% may be clinical-only); under-coded mononucleosis in the negative arm; and non-GAS bacterial pharyngitis (*F. necrophorum*, *Mycoplasma*, *Chlamydophila*) in the negative arm complicating any downstream antibiotic-decision framing.

#### 3.9 B08: Influenza vs other acute respiratory infection

B08 separates ICD-coded influenza from other acute respiratory infection. It is the most imbalanced binary in the panel.

###### 3.9.0.1 Class definitions (final).

The positive class is defined by a primary ICD-10 code starting with J09, J10, or J11. The negative class is defined by a primary ICD-10 code in URI (J00–J06), LRI (J12–J22), or COVID-19 (U07.1, U09, J12.82), with the additional rule that no flu code appears anywhere on the visit. Final cohort: 13,248 influenza and 268,314 other-acute-respiratory visits (class ratio 1:20.3).

###### 3.9.0.2 Expected performance ceiling.

AUC approximately 0.70–0.75 against a hypothetical laboratory-confirmed gold standard. Documented primary-care flu coding shows 61% under-coding of laboratory-confirmed cases and a clinical-criteria positive predictive value of 60% (peak-season 68.5%) against RT-PCR confirmation (Mira-Iglesias et al., 2025b). Adult respiratory syncytial virus (RSV) produces a flu-like syndrome that is acoustically near-indistinguishable from influenza.

###### 3.9.0.3 Key review decisions.

J11.1 (unidentified influenza with other respiratory manifestations) is expected to dominate the positive arm (∼60–70% of primary-care flu) and is the lowest-fidelity flu label. Adult RSV in the negative arm is identifiable from J12.1, J20.5, J21.0, and the etiologic B97.4 code.

A within-season pathogen-substitution effect (RSV peaking November–December, flu peaking January–February, COVID having its own winter wave) remains uncontrolled at the weekly level even after monthly matching and is reported as a methods limitation. Vaccination metadata is not available and the residual mild-systemic-symptoms-after-vaccination effect is reported as a methods limitation; Z23 (vaccination encounter) exclusion does not catch the illness-visit-after-Z23-encounter case.

#### 3.10 B09: COVID-19 vs other acute respiratory infection

B09 separates ICD-coded acute COVID-19 from other acute respiratory infection. Performance is expected to be era-bimodal.

###### 3.10.0.1 Class definitions (final).

The positive class is defined by a primary ICD-10 code in U07.1 (COVID-19, virus identified) or J12.82 (pneumonia due to COVID-19). The negative class is defined by a primary ICD-10 code in URI (J00–J06), LRI (J12 excluding J12.82, J13–J22), or influenza (J09–J11), with the additional rule that no COVID code (U07.1, U09, J12.82) appears anywhere on the visit. Final cohort: ∼15,750 COVID and ∼264,933 non-COVID-acute-respiratory visits (class ratio ∼1:16.8).

###### 3.10.0.2 Key review decisions.

U09.x (“post-COVID-19 condition”) was removed from the positive predicate as a chronic or post-acute sequela rather than acute infection, dropping 666 visits (∼4%) from the positive arm; U09.x visits are still excluded from the negative arm via the visit-level COVID-anywhere rule and therefore drop from B09 entirely. R43.0 (anosmia) era-conditional exclusion was declined under the real-world cohort principle.

Long-COVID baseline contamination in the negative arm (∼60–80% of US adults have prior COVID exposure by 2026, ∼5–17% with persistent voice alterations) (Lin et al., 2023) is reported as an irreducible methods limitation without serology data. Provider-level coding heterogeneity (U07.1 positive predictive value 77.7–93.8% across clinical settings (Lynch et al., 2021)) is deferred to the fairness and robustness audit, where provider-stratified cross-validation is reported.

#### 3.11 B10: Bacterial vs viral respiratory infection

B10 is the panel's hardest binary by design. A literature check during review found no published pooled bacterial-vs-viral voice or cough signature in adult primary-care spontaneous speech (Renjini et al., 2021). The binary collapses seven mechanistically distinct phenotypes under a single etiologic label and is reported with explicit anatomy-mix caveats.

##### *3.11.0.1 Class definitions (final).*

The positive (definite bacterial) class is defined by a primary ICD-10 code in J13 (*Streptococcus pneumoniae* pneumonia), J14 (*Haemophilus influenzae* pneumonia), J15 (other bacterial pneumonia, including J15.7 Mycoplasma), J02.0 (GAS pharyngitis), J03.00 / J03.01 (GAS tonsillitis, acute and recurrent), A37 (pertussis), or A15 (respiratory tuberculosis). The negative (definite viral) class is defined by a primary ICD-10 code in J12 excluding J12.82 (viral pneumonia, excluding COVID variants), J00 (common cold), or J09–J11 (influenza). Ambiguous codes (J06 unspecified URI, J20 acute bronchitis, J01 acute sinusitis, J03.8 / J03.9 other or unspecified tonsillitis, J02.8 / J02.9 other or unspecified pharyngitis) are in neither arm. U07.1 (COVID-19) is in neither arm since COVID has its dedicated B09 binary. Visits with co-coded bacterial and viral codes are dropped. Final cohort: 15,256 bacterial and ∼16,899 viral visits (class ratio ∼1:1.1).

##### *3.11.0.2 Key review decisions.*

U07.1 was removed from the viral predicate during review, restoring internal consistency with the existing J12.82 COVID exclusion and respecting B09's jurisdiction over COVID-19. The change removed 15,723 visits from the viral arm (48%), improving class balance from ∼1:2.1 to ∼1:1.1 and reframing B10 as a GAS-pharyngitis-versus-non-COVID-viral contrast rather than the prior GAS-pharyngitis-versus-COVID-plus-flu contrast. With 92% of the bacterial arm being J02.0 (URI) and the viral arm spanning J00 (URI) plus J09–J11 (mixed) plus J12 viral pneumonia (LRI), B10 largely measures URI-versus-LRI anatomy mix and entity-specific signals (pertussis cough, J00 nasal-coupling, tonsillar mass effect) rather than generic bacterial-versus-viral host-response biology.

A37 ($n = 28$) and A15 ($n = 14$) were retained under the real-world principle as bacterial-by-taxonomy entries. J15.7 (Mycoplasma, $n = 152$, ∼1% of the bacterial arm) was retained despite its viral-like clinical phenotype (gradual onset, dry cough, low-grade fever, outpatient). The expected ∼5–15% GAS-carrier-state false-positive rate on J02.0 is reported as an upper bound on the bacterial-URI sub-binary AUC. Adult pertussis is acknowledged to typically present as prolonged cough without the classic pediatric whoop, and pertussis sub-class AUC is reported separately rather than pooled into the bacterial arm.

##### *3.11.0.3 Acknowledged limitations.*

No published pooled bacterial-vs-viral acoustic signature in adult primary-care spontaneous speech; anatomy-mix dominance over host-response biology; hidden bacterial superinfection in the viral arm (irreducible without antibiotic-prescription metadata); GAS pharyngeal carrier-state false-positives on J02.0;

**Table S2.** Summary statistics by binary and class. Age and audio duration are reported as mean (±SD) and median (interquartile range). Female / male percentages exclude a small fraction of visits with non-binary or missing gender. Geography is summarised by share of visits from California (the modal state in all binaries) and by the number of unique states and cities represented.

| Binary | Class | Visits | Patients | Age mean (SD) | Age median (IQR) | %F | %CA | States | Duration s, median (IQR) |
|---|---|---|---|---|---|---|---|---|---|
| B00 | pos | 255,516 | 206,145 | 41.9 (17.1) | 38 (28–53) | 62.2 | 54.6 | 109 | 24.9 (14.0–41.3) |
| B00 | neg | 178,806 | 146,838 | 43.6 (18.3) | 39 (28–57) | 65.1 | 63.6 | 100 | 31.9 (17.3–55.0) |
| B01 | pos | 270,296 | 215,776 | 42.0 (17.1) | 38 (28–54) | 62.6 | 54.8 | 110 | 25.1 (14.1–41.7) |
| B01 | neg | 11,109 | 9,296 | 43.8 (18.1) | 39 (29–58) | 63.1 | 56.8 | 58 | 31.6 (17.7–52.6) |
| B02 | pos | 271,849 | 216,801 | 42.0 (17.1) | 38 (28–54) | 62.6 | 54.8 | 110 | 25.1 (14.2–41.8) |
| B02 | neg | 31,048 | 29,238 | 44.8 (18.5) | 41 (30–59) | 59.0 | 61.4 | 59 | 31.7 (18.4–52.4) |
| B03 | pos | 29,716 | 25,991 | 49.8 (18.6) | 48 (35–65) | 58.4 | 55.8 | 68 | 28.3 (16.5–46.1) |
| B03 | neg | 182,868 | 155,425 | 40.0 (16.1) | 36 (27–50) | 63.3 | 54.9 | 101 | 24.5 (13.8–40.7) |
| B04 | pos | 10,971 | 9,279 | 50.2 (19.1) | 48 (35–66) | 57.0 | 60.0 | 60 | 29.5 (16.9–47.8) |
| B04 | neg | 8,870 | 8,491 | 45.9 (17.2) | 43 (32–60) | 62.9 | 54.1 | 53 | 30.5 (17.6–48.9) |
| B05 | pos | 10,688 | 9,044 | 50.2 (19.1) | 48 (35–66) | 57.1 | 60.2 | 60 | 29.5 (16.8–47.8) |
| B05 | neg | 25,960 | 23,589 | 49.1 (18.3) | 47 (34–64) | 60.2 | 54.2 | 64 | 28.3 (16.5–46.0) |
| B06 | pos | 45,093 | 40,334 | 44.0 (16.2) | 41 (31–56) | 66.6 | 53.8 | 71 | 29.5 (17.6–47.5) |
| B06 | neg | 159,403 | 137,412 | 39.1 (16.1) | 35 (26–48) | 62.8 | 55.6 | 95 | 23.5 (13.2–39.3) |
| B07 | pos | 11,599 | 10,913 | 33.3 (11.4) | 32 (24–39) | 64.6 | 54.4 | 54 | 18.7 (10.4–32.1) |
| B07 | neg | 48,489 | 45,017 | 35.5 (13.9) | 32 (25–42) | 63.2 | 59.4 | 76 | 21.8 (12.1–36.8) |
| B08 | pos | 12,593 | 12,276 | 39.4 (15.3) | 37 (27–48) | 58.4 | 48.6 | 58 | 20.8 (11.4–35.0) |
| B08 | neg | 258,782 | 207,311 | 42.2 (17.2) | 38 (28–54) | 62.8 | 55.2 | 108 | 25.4 (14.3–42.1) |
| B09 | pos | 15,108 | 14,512 | 50.7 (18.7) | 50 (35–67) | 59.5 | 52.1 | 58 | 24.7 (13.9–41.7) |
| B09 | neg | 255,421 | 204,817 | 41.5 (16.9) | 38 (28–53) | 62.8 | 55.0 | 108 | 25.2 (14.2–41.8) |
| B10 | pos | 14,638 | 13,681 | 33.9 (12.1) | 32 (24–40) | 64.0 | 56.0 | 58 | 19.3 (10.6–33.0) |
| B10 | neg | 16,068 | 15,680 | 39.9 (15.9) | 37 (27–49) | 58.7 | 50.2 | 60 | 21.3 (11.7–36.4) |

uncertain pediatric-to-adult pertussis transfer; Mycoplasma bacterial-by-taxonomy / viral-by-phenotype. An AUC near chance is reported as a meaningful empirical result consistent with the literature null.

#### 4 COHORT DEMOGRAPHICS, GEOGRAPHY, AND AUDIO DURATION

This section reports per-binary summary statistics for age, sex, geography, and audio duration stratified by class. All counts are taken from the feature-available cohort (the joined intersection of the cohort SQL output and the acoustic feature manifest used for training and evaluation), not from the raw cohort SQL row counts referenced in §3.

Geography is summarised by the share of visits from California, the single largest state in the dataset across all binaries, alongside the count of unique US states and cities represented. Audio duration is the per-visit total clip duration in seconds, taken from the extracted feature stream. The recording window spans roughly two calendar years (January 2024 through January–February 2026) for every binary.

##### 4.1 B00: acute respiratory vs acute non-respiratory illness

The two arms are well matched on age and sex: positive class mean age 41.9 years (SD 17.1) versus negative class 43.6 (SD 18.3), and female share 62.2% versus 65.1%. California contributes 54.6% of positive visits and a higher 63.6% of negative visits; New Jersey is correspondingly under-represented on the negative side (11.3% versus 19.1%). Both arms span 100+ states and 6,500+ cities, indicating broad geographic coverage with a California-dominant mode. The positive class is acoustically shorter on average, with median clip duration 24.9 s (IQR 14.0–41.3) versus 31.9 s (17.3–55.0) on the negative side, consistent with the brevity of acute upper respiratory presentations relative to the heterogeneous acute non-respiratory complaint pool.

##### 4.2 B01: acute respiratory infection vs chronic respiratory disease management

Age and sex are tightly matched (positive 42.0 y, 62.6% female; negative 43.8 y, 63.1% female) despite the strong cohort imbalance (24.3:1 by visit). The negative class spans 58 states and 1,471 cities, noticeably narrower than the positive's 110 states and 8,033 cities, reflecting both the smaller sample and the clinic-set bias toward locations with established chronic-respiratory patient panels (California share rises from 54.8% to 56.8% with larger gains for Arizona, 5.3% to 7.1%). Median clip duration is again longer on the negative side (31.6 s versus 25.1 s), consistent with longer-format chronic-disease visits.

##### 4.3 B02: acute respiratory infection vs symptomatic non-infectious respiratory presentation

The negative arm is slightly older on average (44.8 vs 42.0 y) and has a noticeably lower female share (59.0% versus 62.6%), reflecting the inclusion of the male-skewed musculoskeletal and respiratory-symptom codes in the negative class. California is over-represented in the negative class (61.4% versus 54.8%) and the negative arm covers a narrower geography (59 states versus 110). The clip-duration gap (negative median 31.7 s versus positive 25.1 s) matches the B00 and B01 pattern of longer recordings for non-acute or chronic-symptom presentations.

##### 4.4 B03: lower vs upper respiratory infection

The strongest demographic gap of the panel: LRI cases (positive) are roughly a decade older than URI cases (mean 49.8 y vs 40.0 y; median 48 vs 36; IQR 35–65 vs 27–50), with a lower female share (58.4% vs 63.3%). The age confound is well-documented in primary care. Both arms have similar California share ($\sim$55%), but New Jersey is over-represented on the LRI side (21.8% versus 18.2%). Median clip duration is 28.3 s for LRI and 24.5 s for URI; the LRI arm records slightly longer, consistent with more extensive cough-event content.

##### 4.5 B04: pneumonia vs acute bronchitis

Pneumonia patients are 4–5 years older on average (50.2 vs 45.9 y) and are less female-skewed (57.0% versus 62.9%). California is over-represented on the pneumonia side (60.0% versus 54.1%); Washington also rises (7.9% versus 6.2%), and New Jersey halves (11.3% versus 19.4%). Audio duration is essentially identical across arms (median 29.5 s versus 30.5 s), reflecting the shared LRI character of the two presentations. This is the panel's most balanced binary (1.24:1 by visit) and one of the better-matched on demographics.

##### 4.6 B05: pneumonia vs non-pneumonia LRI

Age is closely matched (50.2 vs 49.1 y, IQRs overlap heavily), unlike the B03 URI-vs-LRI contrast, both arms are LRI populations and inherit a similar older-adult demographic. Sex is also more balanced (57.1% vs 60.2% female). California share is elevated on the pneumonia side (60.2% versus 54.2%) and New Jersey is elevated on the non-pneumonia LRI side (25.9% versus 11.0%), the largest single-state geographic skew in the panel; this is plausibly a coding-practice signature of one or more high-volume clinic networks. Median clip duration is 29.5 s versus 28.3 s.

##### 4.7 B06: acute sinusitis vs other URI

J01-coded sinusitis patients are about 5 years older than the other URI pool (mean 44.0 vs 39.1 y) and have a higher female share (66.6% versus 62.8%) consistent with adult female predominance in chronic-and-acute rhinosinusitis presentations. California share is slightly lower in the positive class (53.8% versus 55.6%) while Arizona rises (7.7% versus 4.5%). Median clip duration is substantially longer in the sinusitis arm (29.5 s versus 23.5 s), giving more material for nasal-coupling and resonance features, which, given the dominant viral-URI fraction inside the J01 class, does not translate to expected high AUC (see §3.7).

##### 4.8 B07: GAS pharyngitis vs other pharyngitis

The pharyngitis cohort is the panel's youngest (positive mean age 33.3 y, SD 11.4; negative 35.5 y, SD 13.9), consistent with the young-adult skew of GAS-positive sore-throat presentations after pediatric exclusion. Sex is similar across arms (64.6% vs 63.2% female). California share is slightly lower on the positive side (54.4% versus 59.4%) with a small Colorado bump (4.5% versus 3.7%). Median clip duration is the shortest in the panel (18.7 s GAS versus 21.8 s other pharyngitis), consistent with brief sore-throat visits in younger adults.

##### 4.9 B08: influenza vs other acute respiratory infection

The positive (flu) arm is slightly younger (mean 39.4 vs 42.2 y) and has a lower female share (58.4% versus 62.8%), consistent with the working-age and male-skewed coding pattern of clinical-flu diagnoses in primary care. The flu arm is under-represented in California (48.6% versus 55.2%) and over-represented in New Jersey (24.0% versus 18.6%), pointing to a regional / seasonal coding pattern that is plausibly entangled with peak-flu-season timing in the dataset. Median clip duration is also shorter (20.8 s versus 25.4 s).

##### 4.10 B09: COVID-19 vs other acute respiratory infection

The COVID-positive arm is roughly a decade older (mean 50.7 vs 41.5 y; median 50 vs 38, IQR 35–67 vs 28–53), reflecting the heavier coding of U07.1 / J12.82 in symptomatic adults with risk factors. Female share is slightly lower (59.5% versus 62.8%). Arizona is over-represented on the COVID side (11.0% versus 4.9%), which combined with the era-bimodal performance expectation (§3.10) is reported through the era-stratified AUC. Median clip duration is essentially identical (24.7 s versus 25.2 s).

##### 4.11 B10: bacterial vs viral respiratory infection

The bacterial arm (dominated 92% by J02.0 GAS pharyngitis) is much younger than the viral arm (mean 33.9 y, IQR 24–40 vs 39.9 y, IQR 27–49), and has a higher female share (64.0% versus 58.7%). This demographic gap is a direct consequence of the bacterial arm collapsing to the GAS-pharyngitis demographic, while the viral arm spans common cold, viral pneumonia, and influenza, and is older and more balanced by sex. Geography is similar (California 56.0% versus 50.2%; New Jersey 11.9% versus 22.2%, the latter again reflecting the influenza-clinic regional pattern from B08). Median clip duration is short (19.3 s versus 21.3 s) given the pharyngitis dominance of the bacterial arm. The anatomy-mix dominance of this binary (§3.11) is partly visible in these demographics: any above-chance separation could be driven by the age and clip-length differences alone.

#### 5 PREVALENCE-PROJECTED PREDICTIVE METRICS

Sensitivity, specificity, and the likelihood ratios reported in the main text are computed within each true class and do not depend on prevalence. The positive and negative predictive values, in contrast, depend on the base rate. Because the test set is balanced at the patient level, the directly observed predictive values are

**Table S3.** Positive and negative predictive values (reported as PPV/NPV) at each binary's natural cohort prevalence and across a prevalence grid, projected from the class-conditional sensitivity and specificity by Bayes' rule. The 50% column is the projection at a 50% base rate and can differ slightly from the empirical operating-point values in the main text because the test set is not exactly balanced.

| Binary | Natural prev. (%) | PPV/NPV at natural | 1% | 5% | 10% | 20% | 50% |
|---|---|---|---|---|---|---|---|
| B00 | 58.4 | 0.736/0.619 | 0.020/0.996 | 0.094/0.977 | 0.180/0.954 | 0.331/0.901 | 0.664/0.695 |
| B01 | 96.1 | 0.972/0.063 | 0.014/0.994 | 0.070/0.969 | 0.136/0.936 | 0.262/0.867 | 0.587/0.620 |
| B02 | 89.7 | 0.926/0.139 | 0.014/0.993 | 0.070/0.964 | 0.138/0.926 | 0.265/0.848 | 0.590/0.583 |
| B03 | 13.8 | 0.209/0.904 | 0.016/0.993 | 0.080/0.966 | 0.155/0.931 | 0.292/0.858 | 0.622/0.602 |
| B04 | 55.4 | 0.620/0.536 | 0.013/0.993 | 0.065/0.965 | 0.128/0.928 | 0.248/0.851 | 0.568/0.589 |
| B05 | 29.3 | 0.368/0.773 | 0.014/0.993 | 0.069/0.964 | 0.135/0.927 | 0.260/0.849 | 0.585/0.585 |
| B06 | 21.9 | 0.328/0.833 | 0.017/0.993 | 0.084/0.964 | 0.162/0.926 | 0.303/0.848 | 0.635/0.583 |
| B07 | 19.2 | 0.228/0.858 | 0.012/0.993 | 0.061/0.965 | 0.121/0.928 | 0.236/0.852 | 0.553/0.589 |
| B08 | 4.7 | 0.070/0.972 | 0.015/0.994 | 0.075/0.971 | 0.145/0.940 | 0.277/0.874 | 0.605/0.634 |
| B09 | 5.8 | 0.098/0.968 | 0.017/0.995 | 0.085/0.972 | 0.163/0.943 | 0.305/0.881 | 0.637/0.649 |
| B10 | 31.9 | 0.465/0.797 | 0.018/0.995 | 0.089/0.972 | 0.171/0.943 | 0.317/0.881 | 0.650/0.648 |

**Table S4.** Operating-point metrics with 95% bootstrap confidence intervals (1,000 resamples) on the held-out test set, at each binary's decision threshold. Each entry is the point estimate followed by the interval in brackets. LR+ and LR− are the positive and negative likelihood ratios.

| Binary | Sensitivity | Specificity | LR+ | LR− | PPV | NPV |
|---|---|---|---|---|---|---|
| B00 | 0.722 [0.718, 0.725] | 0.635 [0.632, 0.640] | 1.98 [1.96, 2.00] | 0.44 [0.43, 0.44] | 0.675 [0.672, 0.679] | 0.685 [0.680, 0.689] |
| B01 | 0.680 [0.665, 0.695] | 0.522 [0.504, 0.539] | 1.42 [1.37, 1.48] | 0.61 [0.58, 0.65] | 0.595 [0.581, 0.610] | 0.612 [0.594, 0.628] |
| B02 | 0.567 [0.557, 0.577] | 0.606 [0.596, 0.615] | 1.44 [1.40, 1.48] | 0.71 [0.70, 0.73] | 0.600 [0.590, 0.610] | 0.573 [0.564, 0.582] |
| B03 | 0.564 [0.554, 0.574] | 0.658 [0.648, 0.667] | 1.65 [1.59, 1.70] | 0.66 [0.64, 0.68] | 0.619 [0.609, 0.629] | 0.605 [0.595, 0.614] |
| B04 | 0.642 [0.622, 0.661] | 0.512 [0.493, 0.531] | 1.32 [1.25, 1.38] | 0.70 [0.66, 0.74] | 0.567 [0.549, 0.586] | 0.591 [0.570, 0.610] |
| B05 | 0.585 [0.568, 0.601] | 0.585 [0.568, 0.602] | 1.41 [1.34, 1.48] | 0.71 [0.68, 0.75] | 0.581 [0.564, 0.597] | 0.589 [0.573, 0.607] |
| B06 | 0.484 [0.476, 0.492] | 0.722 [0.714, 0.729] | 1.74 [1.69, 1.80] | 0.71 [0.70, 0.73] | 0.626 [0.616, 0.635] | 0.593 [0.585, 0.600] |
| B07 | 0.694 [0.679, 0.709] | 0.440 [0.423, 0.456] | 1.24 [1.19, 1.28] | 0.70 [0.65, 0.74] | 0.555 [0.540, 0.569] | 0.588 [0.568, 0.607] |
| B08 | 0.679 [0.663, 0.693] | 0.557 [0.542, 0.572] | 1.53 [1.47, 1.59] | 0.58 [0.55, 0.61] | 0.595 [0.581, 0.609] | 0.644 [0.627, 0.661] |
| B09 | 0.664 [0.651, 0.677] | 0.622 [0.609, 0.637] | 1.76 [1.68, 1.83] | 0.54 [0.52, 0.56] | 0.639 [0.625, 0.652] | 0.648 [0.636, 0.662] |
| B10 | 0.647 [0.632, 0.661] | 0.652 [0.638, 0.665] | 1.86 [1.77, 1.94] | 0.54 [0.52, 0.57] | 0.640 [0.626, 0.653] | 0.658 [0.644, 0.672] |

at a 50% base rate. Table S3 re-projects the predictive values to each binary's natural cohort prevalence (the positive rate in the pre-balance clean manifest) and to a grid of prevalences, by applying Bayes' rule to the class-conditional sensitivity and specificity. For the low-prevalence pathogen contrasts B08 (influenza) and B09 (COVID-19), the natural-prevalence positive predictive value is 0.07 and 0.10 respectively, while the negative predictive value is 0.97 in both, indicating that the only operating use these models support at their natural prevalence is rule-out.

Table S4 reports 95% bootstrap confidence intervals for the operating-point metrics of Table 3 in the main text. Each interval was obtained by resampling the held-out test predictions of the corresponding binary with replacement (1,000 resamples) and recomputing the metric at the binary's decision threshold; the point estimates coincide with those in Table 3.

#### 6 DECISION-CURVE ANALYSIS

Decision-curve analysis evaluates whether acting on the model's predictions yields more net benefit than the default strategies of treating all patients or treating none, across the range of threshold probabilities a

**Table S5.** Leave-one-family-out AUC change per binary (full model AUC minus the AUC with that family removed; single fold). A positive value means removing the family lowers AUC, so the family contributes.

| Binary | Full AUC | Pitch | Loud. | Spec. | Voice qual. | Form. | Temp. | Speech rate/pause | VAD | librosa |
|---|---|---|---|---|---|---|---|---|---|---|
| B00 | 0.747 | 0.002 | 0.002 | 0.006 | 0.002 | 0.006 | 0.002 | 0.001 | 0.007 | 0.025 |
| B01 | 0.638 | 0.005 | 0.001 | −0.006 | −0.004 | −0.003 | −0.004 | −0.004 | −0.002 | 0.014 |
| B02 | 0.626 | 0.004 | 0.004 | 0.003 | 0.004 | 0.004 | 0.003 | 0.005 | 0.007 | 0.009 |
| B03 | 0.656 | 0.002 | 0.002 | 0.001 | 0.003 | 0.002 | 0.002 | 0.001 | 0.007 | 0.017 |
| B04 | 0.604 | −0.001 | 0.010 | 0.001 | −0.007 | −0.005 | −0.002 | 0.003 | 0.005 | 0.023 |
| B05 | 0.633 | 0.000 | −0.001 | 0.002 | 0.008 | 0.003 | 0.000 | −0.003 | 0.001 | 0.029 |
| B06 | 0.649 | −0.002 | −0.002 | −0.001 | 0.001 | 0.003 | 0.001 | −0.001 | 0.000 | 0.011 |
| B07 | 0.604 | −0.002 | 0.001 | 0.002 | 0.000 | 0.000 | 0.001 | 0.001 | 0.003 | 0.031 |
| B08 | 0.657 | −0.005 | −0.003 | −0.003 | −0.004 | −0.006 | −0.005 | −0.004 | 0.000 | 0.017 |
| B09 | 0.687 | 0.000 | 0.001 | 0.001 | 0.003 | 0.003 | −0.001 | 0.000 | 0.000 | 0.017 |
| B10 | 0.703 | 0.002 | 0.002 | 0.002 | 0.001 | 0.010 | 0.008 | 0.001 | 0.007 | 0.046 |

clinician might adopt. At a threshold probability $p_t$, a visit is called positive if its predicted probability is at least $p_t$, and the net benefit is $\text{TP}/N - (\text{FP}/N)\,[\,p_t/(1-p_t)\,]$, where $N$ is the number of test visits. The threshold probability encodes the weight placed on a false positive relative to a true positive. Figure S5 shows the net-benefit curves for all 11 binaries, computed on the balanced (approximately 50%) held-out test set, so the curves and the treat-all reference are at a 50% base rate; predictive values at each contrast's natural prevalence are given in Table S3. Across the panel, the voice model's net-benefit curve lies at or above both the treat-all and treat-none references over the lower and middle range of threshold probabilities.

#### 7 FEATURE-FAMILY ABLATION

This section reports the full per-family feature ablation summarized in the main text. For each binary, the model was retrained on the full set of nine feature streams, on each single stream alone, and on the full set with one family removed (leave-one-family-out). All ablation runs use a single cross-validation fold, so the full-set AUC is the single-fold counterpart of the cross-validated AUC reported in the main text, and differences below approximately 0.01 are within run-to-run variation. The families are pitch (openSMILE F0 descriptors), loudness, spectral, voice quality (jitter, shimmer, harmonics-to-noise ratio), formants, temporal (openSMILE timing descriptors), speech rate and pause (praat-parselmouth syllable and pause statistics), VAD (the valence-arousal-dominance affective stream), and librosa (Mel-frequency cepstral, spectral, chromagram, and rhythm features).

Table S5 reports the leave-one-family-out AUC change (full minus leave-one-out; positive means the family contributes). Table S6 reports the AUC of each family in isolation.

#### 8 LOCATION-STRATIFIED GENERALIZATION

This section reports the location analyses summarized in the main text. In the tables below, champion AUC is the random-split test AUC reported in Table 2 of the main text. The dataset carries no clinic or site identifiers, so location is represented by the patient city and state of residence. The city-grouped held-out experiment is reported in the main text; Table S7 reports the California-versus-rest experiment and Table S8 the per-site stratified AUC.

**Table S6.** Single-family AUC per binary (each feature family evaluated in isolation; single fold).

| Binary | Pitch | Loud. | Spec. | Voice qual. | Form. | Temp. | Speech rate/pause | VAD | librosa |
|---|---|---|---|---|---|---|---|---|---|
| B00 | 0.575 | 0.578 | 0.659 | 0.599 | 0.632 | 0.604 | 0.600 | 0.620 | 0.719 |
| B01 | 0.563 | 0.556 | 0.574 | 0.546 | 0.566 | 0.555 | 0.578 | 0.582 | 0.631 |
| B02 | 0.551 | 0.554 | 0.566 | 0.544 | 0.548 | 0.545 | 0.592 | 0.588 | 0.612 |
| B03 | 0.571 | 0.575 | 0.609 | 0.568 | 0.585 | 0.568 | 0.565 | 0.594 | 0.641 |
| B04 | 0.548 | 0.544 | 0.571 | 0.545 | 0.555 | 0.572 | 0.526 | 0.553 | 0.597 |
| B05 | 0.562 | 0.566 | 0.587 | 0.568 | 0.546 | 0.596 | 0.537 | 0.581 | 0.632 |
| B06 | 0.564 | 0.567 | 0.589 | 0.554 | 0.593 | 0.561 | 0.587 | 0.595 | 0.638 |
| B07 | 0.528 | 0.542 | 0.551 | 0.529 | 0.537 | 0.552 | 0.544 | 0.551 | 0.592 |
| B08 | 0.574 | 0.558 | 0.593 | 0.577 | 0.574 | 0.570 | 0.581 | 0.585 | 0.650 |
| B09 | 0.529 | 0.575 | 0.629 | 0.571 | 0.591 | 0.594 | 0.539 | 0.541 | 0.679 |
| B10 | 0.545 | 0.559 | 0.622 | 0.580 | 0.608 | 0.599 | 0.532 | 0.566 | 0.689 |

**Table S7.** California-versus-rest cross-region generalization. “Train CA, test rest” trains on California visits and tests on the rest of the country; “Train rest, test CA” is the reverse. Reported for the subset of binaries run in this experiment.

| Binary | Champion AUC | Train CA, test rest | Train rest, test CA |
|---|---|---|---|
| B00 | 0.745 | 0.723 | 0.722 |
| B05 | 0.629 | 0.562 | 0.525 |
| B08 | 0.658 | 0.610 | 0.617 |
| B09 | 0.694 | 0.640 | 0.669 |
| B10 | 0.703 | 0.644 | 0.664 |

**Table S8.** Per-site stratified AUC from the champion test predictions (no retraining). Within-city and within-state AUC are computed over sites with at least 50 test cases of each class and aggregated as a visit-weighted mean; the city minimum and maximum show the spread across qualifying cities.

| Binary | Overall AUC | City mean | City min | City max | State mean |
|---|---|---|---|---|---|
| B00 | 0.745 | 0.740 | 0.619 | 0.926 | 0.739 |
| B01 | 0.643 | 0.658 | 0.483 | 0.801 | 0.638 |
| B02 | 0.623 | 0.614 | 0.333 | 0.845 | 0.616 |
| B03 | 0.657 | 0.639 | 0.374 | 0.813 | 0.654 |
| B04 | 0.615 | 0.612 | 0.452 | 0.717 | 0.604 |
| B05 | 0.629 | 0.569 | 0.440 | 0.714 | 0.599 |
| B06 | 0.651 | 0.635 | 0.487 | 0.790 | 0.648 |
| B07 | 0.602 | 0.595 | 0.492 | 0.692 | 0.598 |
| B08 | 0.658 | 0.656 | 0.554 | 0.743 | 0.655 |
| B09 | 0.694 | 0.693 | 0.523 | 0.801 | 0.686 |
| B10 | 0.703 | 0.693 | 0.569 | 0.798 | 0.695 |

#### 9 PER-CLASS AGGREGATE FEATURE DISTRIBUTIONS

We provide, as supplementary data, the per-class aggregate distribution of every feature for every binary: for each of the 11 binaries and each of the 412 features, the per-class count, mean, standard deviation, and median, together with the mean difference, pooled standard deviation, and Cohen’s d. The file contains

**Table S9.** Feature with the largest absolute Cohen's d in each binary, from the per-class aggregate feature distributions. Cohen's d is signed (positive means a higher mean in the positive class).

| Binary | Feature (largest $|d|$) | Stream | Cohen's d |
|---|---|---|---|
| B00 | total syllable count | speech rate/pause | $-0.340$ |
| B01 | total syllable count | speech rate/pause | $-0.337$ |
| B02 | total syllable count | speech rate/pause | $-0.322$ |
| B03 | MFCC 9 minimum | librosa | $-0.191$ |
| B04 | openSMILE MFCC3 mean | openSMILE | 0.176 |
| B05 | MFCC 10, 75th percentile | librosa | 0.232 |
| B06 | total syllable count | speech rate/pause | 0.249 |
| B07 | phonation time | speech rate/pause | $-0.169$ |
| B08 | phonation time | speech rate/pause | $-0.215$ |
| B09 | spectral rolloff mean | librosa | $-0.267$ |
| B10 | spectral rolloff mean | librosa | 0.297 |

$11 \times 412 = 4{,}532$ rows and carries no patient identifiers, no per-visit values, no dates, and no geography, so it can be released without disclosing protected health information. No single feature is strongly discriminative in any binary: the maximum absolute Cohen's d across the panel is approximately 0.34 and no feature exceeds 0.5, consistent with the moderate AUCs. The largest single-feature effects include the speech-rate and pause family, which the ablation in Section 7 shows the trained models nonetheless rely on little. Table S9 lists, for each binary, the feature with the largest absolute standardized effect.

#### REFERENCES


Arrazola, R. A., Husten, C. G., Cornelius, M. E., and Armour, B. S. (2025). Notes from the field: Tobacco product use among adults — united states, 2017–2023. *MMWR. Morbidity and Mortality Weekly Report* 74, 118–121. doi:10.15585/mmwr.mm7407a3. Any tobacco product use 19.5% (2023). Cigarette smoking 9.9% (2024 NHIS, CDC FastStats) cited alongside per author decision Q4.

Centor, R. M., Witherspoon, J. M., Dalton, H. P., Brody, C. E., and Link, K. (1981). The diagnosis of strep throat in adults in the emergency room. *Medical Decision Making* 1, 239–246. doi:10.1177/0272989X8100100304

Chow, A. W., Benninger, M. S., Brook, I., Brozek, J. L., Goldstein, E. J. C., Hicks, L. A., et al. (2012). IDSA clinical practice guideline for acute bacterial rhinosinusitis in children and adults. *Clinical Infectious Diseases* 54, e72–e112. doi:10.1093/cid/cis370. Bacterial superinfection complicates 0.5-2.0% of viral URIs (reword from "10-15%" per Q11).

Fine, A. M., Nizet, V., and Mandl, K. D. (2012). Large-scale validation of the Centor and McIsaac scores to predict group A streptococcal pharyngitis. *Archives of Internal Medicine* 172, 847–852. doi:10.1001/archinternmed.2012.950. Centor score AUC 0.72; McIsaac score AUC 0.71 (use exact values per Q12).

Guevara, R. E., Butler, J. C., Marston, B. J., Plouffe, J. F., File, T. M., and Breiman, R. F. (1999). Accuracy of ICD-9-CM codes in detecting community-acquired pneumococcal pneumonia for incidence and vaccine efficacy studies. *American Journal of Epidemiology* 149, 282–289. doi:10.1093/oxfordjournals.aje.a009804. Sensitivity 14.2% for code 486.0 (pneumonia, organism unspecified) vs lab-confirmed pneumococcal pneumonia.

Lin, C.-W., Wang, Y.-H., Li, Y.-E., Chiang, T.-Y., Chiu, L.-W., Lin, H.-C., et al. (2023). COVID-related dysphonia and persistent long-COVID voice sequelae: A systematic review and meta-analysis. *American*

*Journal of Otolaryngology* 44, 103950. doi:10.1016/j.amjoto.2023.103950. 17.1% post-recovery dysphonia; 20.1% long-COVID dysphonia (drop the unsourced 5% lower bound per Q13).

Lynch, K. E., Viernes, B., Gatsby, E., DuVall, S. L., Jones, B. E., Box, T. L., et al. (2021). Positive predictive value of COVID-19 ICD-10 diagnosis codes across calendar time and clinical setting. *Clinical Epidemiology* 13, 1011–1018. doi:10.2147/CLEP.S335621. U07.1 PPV 77.7% (outpatient) to 93.8% (inpatient); overall 84.2%. VA data.

McIsaac, W. J., White, D., Tannenbaum, D., and Low, D. E. (1998). A clinical score to reduce unnecessary antibiotic use in patients with sore throat. *CMAJ: Canadian Medical Association Journal* 158, 75–83

Mira-Iglesias, A., López-Lacort, M., Bricout, H., Loiacono, M., Carballido-Fernández, M., Mollar-Maseres, J., et al. (2025a). Accuracy of ICD influenza discharge diagnosis codes in hospitalized adults from the valencia region, spain, 2012/2013 to 2017/2018. *Influenza and Other Respiratory Viruses* 19, e70069. doi:10.1111/irv.70069. HOSPITAL discharge data (relabel; not primary care). Adjusted sensitivity 25.3%; 61% of lab-confirmed cases not ICD-coded.

Mira-Iglesias, A., López-Lacort, M., Bricout, H., Loiacono, M., Carballido-Fernández, M., Mollar-Maseres, J., et al. (2025b). Accuracy of ICD influenza discharge diagnosis codes in hospitalized adults from the valencia region, spain, 2012/2013 to 2017/2018. *Influenza and Other Respiratory Viruses* 19, e70069. doi:10.1111/irv.70069. 61% of lab-confirmed influenza not ICD-coded (HOSPITAL discharge). The ”clinical-criteria PPV 60%/68.5%” claim was NOT verifiable and should be dropped.

National Center for Health Statistics (2017). Quickstats: Rate of visits to office-based physicians, by patient age and sex — national ambulatory medical care survey, united states, 2015. *MMWR. Morbidity and Mortality Weekly Report* 66, 1060. doi:10.15585/mmwr.mm6639a6. Females 362 vs males 262 office visits per 100 persons.

Renjini, A., Swapna, M. S., Raj, V., and Sankararaman, S. (2021). Graph-based feature extraction and classification of wet and dry cough signals: a machine learning approach. *Journal of Complex Networks* 9, cnab039. doi:10.1093/comnet/cnab039. Wet-vs-dry cough classification, framed as bacterial-vs-viral proxy; classifies cough morphology, NOT validated etiology. Cite to acknowledge adjacent work; the validated bacterial-vs-viral acoustic gap (adults) remains.

Williamson, I. J., Matusiewicz, S. P., Brown, P. H., Greening, A. P., and Crompton, G. K. (1995). Frequency of voice problems and cough in patients using pressurized aerosol inhaled steroid preparations. *European Respiratory Journal* 8, 590–592. doi:10.1183/09031936.95.08040590. Classic prevalence study of ICS-related voice problems. PMID 7664859.

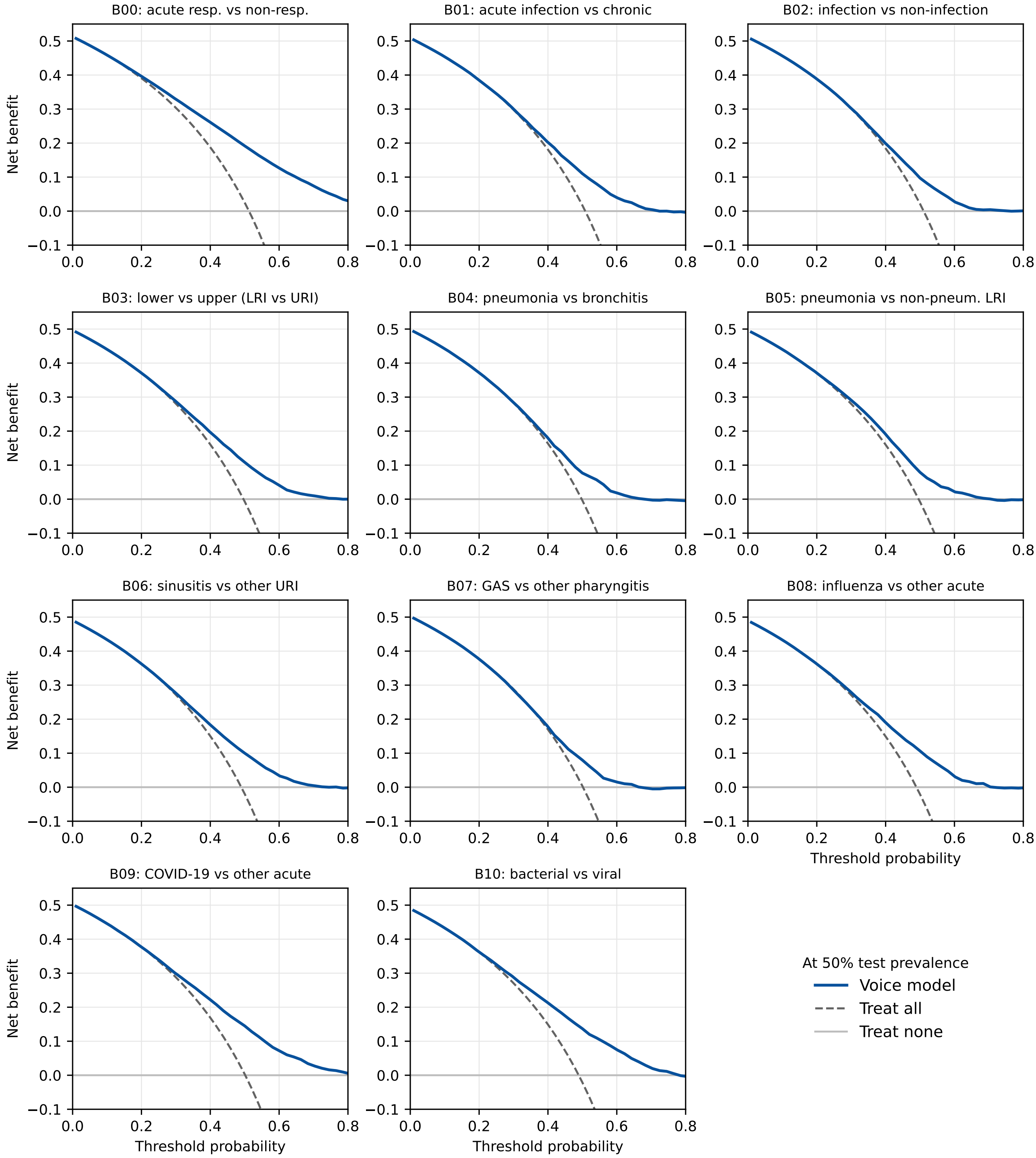


**Figure S5.** Decision-curve analysis for the 11 binary classification tasks on the held-out test set at the balanced 50% prevalence. Each panel plots net benefit against threshold probability for the voice model (solid), a treat-all strategy (dashed), and a treat-none strategy (net benefit zero). A model provides net benefit over the default strategies where its curve lies above both references; these curves are at the balanced 50% test prevalence and do not by themselves establish deployment utility at each contrast's natural prevalence.
